# Proteomic Signatures Capture Environmental Exposures Across Diverse Populations

**DOI:** 10.64898/2026.10.06.26364311

**Authors:** Yon Ho Jee, Kristin Tsuo, Sophie Parsa, M. Austin Argentieri, Shakson Isaac, Amanda Elliott, Raymond Walters, Evin Padhi, Elizabeth Kiernan, Lee Lichtenstein, Michael E. Talkowski, Stephen B. Montgomery, Niall J. Lennon, Chirag J. Patel, Konrad J. Karczewski, Mark J. Daly, Alicia R. Martin

**Affiliations:** Analytic and Translational Genetics Unit, Massachusetts General Hospital, Boston, Massachusetts 02114, USA; Program in Medical and Population Genetics, Broad Institute of MIT and Harvard, Cambridge, Massachusetts 02142, USA; Harvard-MIT Program in Health Sciences and Technology, Harvard Medical School, Boston, MA 02446, USA; Department of Biomedical Informatics, Harvard Medical School, Boston, MA 02215, USA; Stanley Center for Psychiatric Research, Broad Institute of MIT and Harvard, Cambridge, MA 02142, USA; Department of Pathology, Stanford University, Stanford, CA 94305, USA; Scalable Services, Broad Institute of MIT and Harvard, Cambridge, MA, USA; Center for Genomic Medicine, Massachusetts General Hospital, Boston, MA, USA; All of Us Research Program, Office of the Director, National Institutes of Health, Bethesda, MD, USA, Department of Genetics, Stanford University, Stanford, CA 94305, USA; Novo Nordisk Foundation Center for Genomic Mechanisms of Disease, Broad Institute of MIT and Harvard, Cambridge, Massachusetts 02142, USA; Institute for Molecular Medicine Finland (FIMM), Helsinki Institute of Life Science, University of Helsinki, Helsinki, Finland

## Abstract

Environmental exposures shape disease risk, but the generalizability of circulating protein-based readouts of exposures across diverse populations remains unknown. Using plasma proteomic profiles of 5,416 proteins in 10,043 participants from the *All of Us* Research Program, we evaluated associations with and composite scores of 100+ exposures, spanning adiposity, air pollution, ambient temperature, and substance use. We find that proteomic exposure signatures are largely consistent across populations and socioeconomic strata, with <1% of proteins showing significant cross-population heterogeneity, which included BMI as an exception, with leptin being the most heterogeneous association. Population-specific leptin trajectories adiposity, together with residual leptin-type 2 diabetes associations after BMI adjustment, revealed sex-specific metabolic risk not captured by BMI alone. Aggregated protein scores were largely exposure-specific, but substance-use scores showed cross-prediction and SES scores were broadly associated. PM_2.5_ protein scores captured within-region exposure variation and outperformed geographic and socioeconomic correlates, but spatial confounders remain embedded within protein-environment associations. Scores trained in UK Biobank transferred to *All of Us* for biologically proximal exposures, supporting cross-biobank portability. These findings establish plasma proteomics as a scalable framework for exposure measurement across diverse populations, while underscoring that responsible applications require careful consideration of biological context, social patterning, and cohort structure.

## Introduction

Environmental exposures shape disease risk across the life course, but they are often measured inconsistently and/or subjectively across datasets, and the molecular processes they perturb remain incompletely characterized. Plasma proteomics is well positioned to inform the molecular processes that environmental exposures perturb because circulating proteins can accurately capture these and downstream physiologic responses. Recent large-scale studies have shown that modifiable lifestyle and clinical exposures, including smoking, alcohol use, dietary behavior, exercise, adiposity, renal function, and inflammation, are associated with widespread variation in the plasma proteome (*1*).

Protein-based models of incident disease outperform traditional clinical models and polygenic risk scores for a wide range of diseases (*2–4*) (e.g. AUC > 0.8 for 92 diseases (*5*)). However, accumulating evidence suggests that many disease-associated proteins are not themselves causal drivers. Instead, a substantial fraction of their predictive signal appears to reflect environmental risk exposure, early physiologic dysregulation, or subclinical disease processes. Notably, over 90% of proteins associated with diseases like lung cancer and chronic obstructive pulmonary disease are also associated with smoking, and disease-associated proteins tested in Mendelian randomization analysis show smoking associations at a markedly higher rate (640 of 782, 82%) than the general background rate across all measured proteins (1,673 of 2,923, 57%) (*2*, *6*). Thus, improving biological interpretation of disease-predictive proteomic signatures will require two complementary approaches: first, identifying proteins causally implicated in disease and defining the mechanisms through which they act; and second (the focus of this study), developing quantitative proteomic measures of environmental exposures that clarify the exposure histories and longitudinal trajectories giving rise to these signatures. Disentangling the causal from the correlative roles those exposures play in disease risk benefits from integration of genomic and environmental data and is a key step towards more precisely intervening to minimize disease risk or progression.

Understanding the degree to which genetic and environmental exposures contribute to proteomic variation requires consideration at individual proteins versus across the entire proteome. At the level of individual proteins, the variance in abundance is shaped by both genetic and environmental architecture, though genetic contributions are more pervasive. Integrating genetic, exposomic, and proteomic data from the UK Biobank Pharma Proteomics Project (UKB-PPP) has revealed extensive genetic regulation, with 2,021 proteins (75% of 2,686 measured proteins) exhibiting genetic *R^2^* values greater than 1%, compared to 926 proteins (34% of measured) influenced by exposomic factors at the same threshold (*7*). In addition, modifiable factors explain meaningful variation across many proteins; body composition traits, such as body mass index (BMI; median 0.36%, up to 7.99%), total fat mass (median 0.55%, up to 40.30%) or peripheral fat (median 0.26%, up to 7.23%) are among the exposures most strongly reflected in circulating protein profiles (*1*). However, in aggregate, proteomic patterns capture substantial non-genetic information to quantify environmental history. Prior studies have developed proteomic scores for smoking and alcohol consumption and demonstrated strong discrimination between current and never users (AUC > 0.95 and 0.98, respectively), supporting the use of plasma proteomic profiles as quantitative measures of environmental exposures (*6*, *8*, *9*). These objective measures are therefore poised more broadly for studies of environmental exposures and health disparities.

A major barrier to broader research and translational use of these findings is uncertainty around the generalizability of proteomic signatures across computed genetic ancestry, race and/or ethnicity, and socioeconomic groups, as many foundational proteomic studies have been conducted in ancestrally, culturally, and socioeconomically homogeneous populations (*10*, *11*). Leveraging the diversity of the *All of Us* (AoU) cohort across these strata, we sought to evaluate proteomic signatures of environmental exposures to characterize their biological and contextual relevance. Specifically, our objectives are to assess their generalizability, biological specificity, and potential to enhance disease risk prediction and health equity across diverse populations.

## Results

### Proteomic signatures capture environmental exposure patterns to varying degrees

#### Proteins associate with many exposures with varying specificity

After quality control of the AoU Olink HT 5k proteomic data, 9,561 samples and 5,414 proteins were retained for analysis (**Fig. 1, fig. S1**, **table S1**). We tested protein associations with 108 health-related exposure variables spanning behavioral, socioeconomic, environmental, anthropometric, clinical and biomarker domains (**table S2**), identifying 62,042 significant protein-exposure pairs at a Bonferroni-corrected threshold. These health-related exposures were selected to reflect domains commonly evaluated in prior social and environmental determinants of health frameworks, expanded to include additional environmental variables available through AoU’s geocoded neighborhood-level data. These should not be viewed as fully environmental, however, as they each carry a heritable component (mean *h^2^*= 0.16, range 0.10-0.28 across trait classes), indicating that even the least heritable exposure category retains a non-trivial genetic contribution, as observed in other biobanks previously (*12*). The proportion of significant proteins per exposure broadly followed the specificity and biological proximity of the trait, from anthropometric (adiposity) and biomarkers (each associated with roughly a third of proteins) to socioeconomic and air pollution measures (each under 10%) (**Fig. 2A, table S3**). About half of the measured proteins were associated with age (49.4%) and BMI (47.7%), followed by sex at birth (referred to hereafter as “sex”; see methods for details) (34.5%). Notably, 18.9% of proteins were associated with current cigarette smoking, a smaller proportion than reported in UKB (57% (*6*)), consistent with reduced statistical power in AoU and reflected by downsampling (**Supplementary text**).

**Fig. 1.**
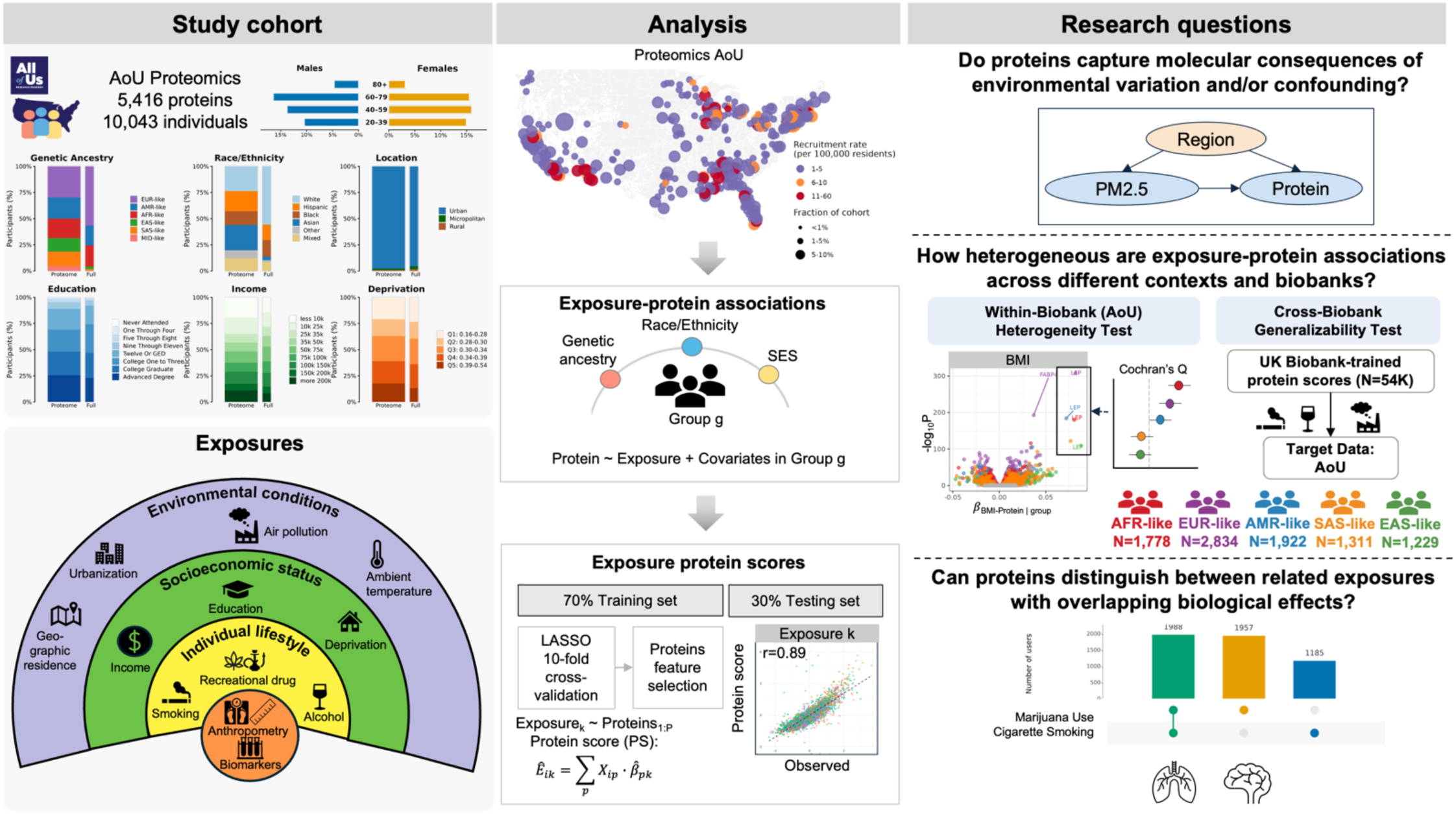
**Linking the plasma proteome to exposures in a diverse, population-scale U.S. biobank**. (**Left**) Study cohort. Plasma proteomics data (5,416 proteins) were measured in 10,043 All of Us (AoU) participants. Panels show sex and age, genetic ancestry (EUR-like, AMR-like, EAS-like, SAS-like, AFR-like), self-reported race and/or ethnicity, as a separate social/ structural construct; geographic setting, education, income, and deprivation index. In stacked bars, the first (thicker) bar is the proteomics AoU subset and the second (thinner) bar is the full AoU cohort. Bottom, exposure hierarchy from environmental conditions to socioeconomic status, lifestyle factors, and anthropometry/biomarkers. (**Middle**) Analysis. Recruitment maps compare full AoU and proteomics AoU samples. Exposure-protein associations are tested across strata of ancestry, race and/or ethnicity, and socioeconomic status. Exposure protein scores are built via LASSO with 10-fold cross-validation (70% training/30% testing) and validated against observed exposures. (**Right**) Research questions: (i) whether proteins capture environmental exposure effects versus regional confounding; (ii) heterogeneity of associations within AoU and across biobanks; (iii) whether protein signatures discriminate related exposures.

**Fig. 2.**
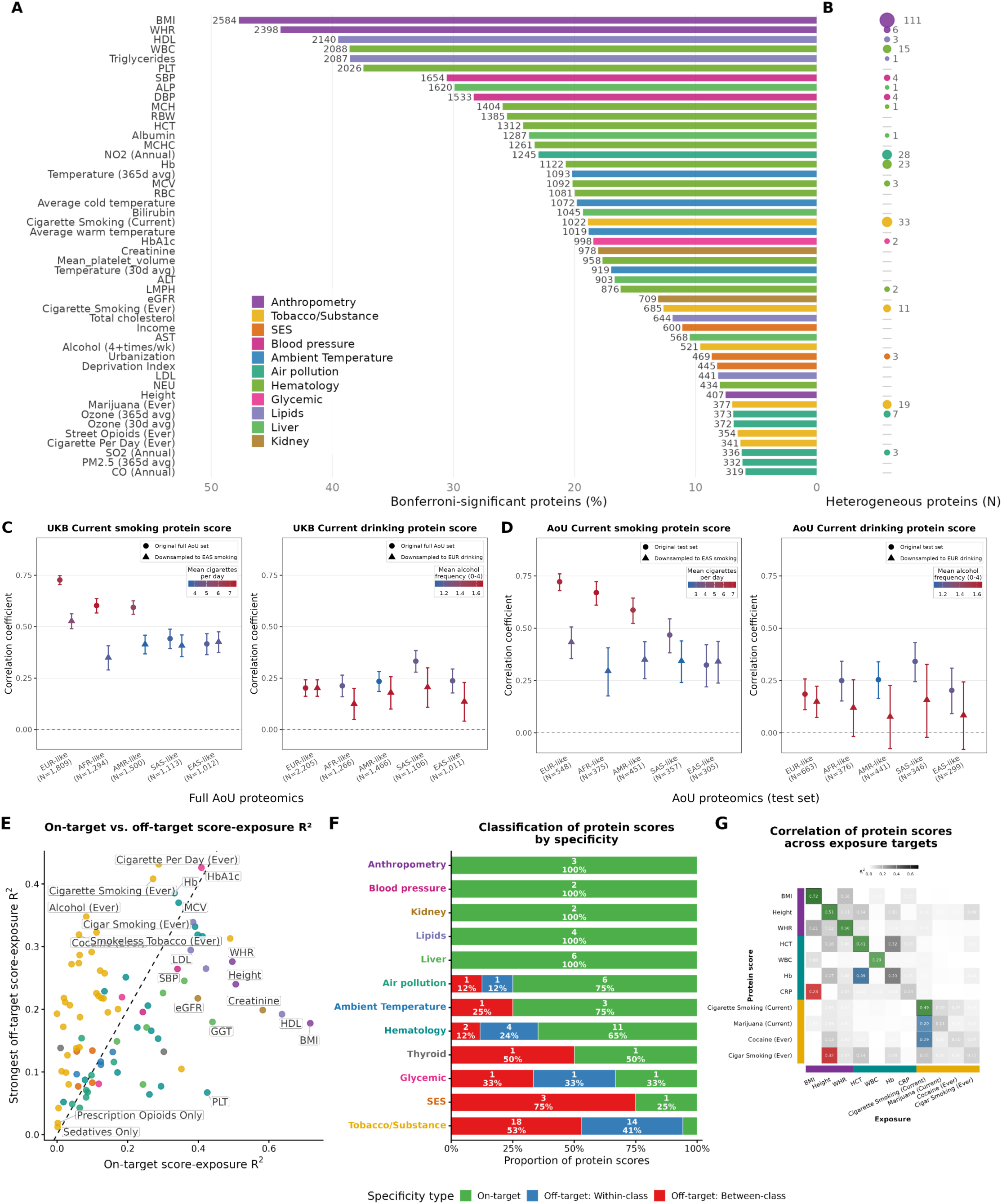
Proteomic signatures of diverse exposures transfer across biobanks, generalize across genetic ancestries, and largely reflect their target exposures. (**A**) Number and percentage of proteins significantly associated with each exposure after Bonferroni correction. Colors denote exposure classes. (**B**) Number of proteins with heterogeneous associations across genetic ancestry groups, defined as proteins with a significant fixed-effect meta-analysis across ancestry groups (Bonferroni-adjusted P < 0.05) and significant heterogeneity between groups (Cochran’s Q, P < 0.05/3,011 independent proteins). (**C**) Cross-cohort validation of UK Biobank (UKB)-trained protein-exposure scores in the All of Us (AoU) full proteomics cohort, before and after downsampling to match the exposure prevalence and intensity distribution of a reference ancestry. Circles: original AoU full proteomics cohort. Triangles: cohort downsampled to jointly match ever-exposure prevalence and intensity to the distribution observed in EAS-like (smoking) or EUR-like (drinking) – the ancestry with the lowest covariate-adjusted point-biserial correlation between protein score and exposure in the original analysis. Point color indicates the matched sample’s mean cigarettes/day (smoking) or mean past-year drinking frequency (0-4 ordinal scale; drinking). (**D**) Within-cohort validation of AoU-trained protein-exposure scores. Scores were trained in a pooled AoU training set and evaluated in a held-out test set (circles), and then retrained after downsampling the training and test sets to match a reference ancestry’s exposure prevalence and intensity distribution (EAS-like for smoking, EUR-like for drinking) (triangles). (**E**) Exposure specificity of protein scores. For each score, the R² for its matched (on-target) exposure is plotted against the largest R² observed for any other (off-diagonal) exposure. Points above the dashed line indicate scores that predict their matched exposure more strongly than any alternative exposure. (**F**) Classification of protein scores by specificity. On-target scores had a matched-exposure R² exceeding all off-diagonal R² values. Among scores with a stronger off-diagonal association, within-class transfer scores were most strongly associated with another exposure in the same exposure class, whereas between-class transfer scores were most strongly associated with an exposure from a different class. (**G**) Correlation structure of representative protein scores across exposure targets. For each score, the strongest score-exposure correlation is colored by specificity category (on-target, within-class transfer, or between-class transfer); the full correlation matrix is shown in **fig. S9**.

We first assessed whether circulating proteins capture molecular signatures of environmental variation beyond broad geographic confounding (**Fig. 1**). For most geocoded exposures, adjustment for region of residence (Northeast, Midwest, West, or South) substantially attenuated the number of significant protein associations (**fig. S2, table S3**), reducing associations by an average of 87.8% for ambient temperature, 30.5% for SES, and 15.3% for air pollution exposures—consistent with a major contribution of geographic structure to these signals. In contrast, PM_2.5_-associated proteins increased after regional adjustment, suggesting that regional differences masked rather than explained within-region associations. Several proteins remained associated with PM_2.5_ (365d avg) after regional adjustment with prior links to respiratory disease biology, including SERPINA1 (*13*), IL1RL1 (*14*), CHI3L1 (*15*), and IL6R (*16*), all inversely associated with PM_2.5_ (**table S4**). These findings suggest the PM_2.5_-associated proteomic signature may capture biologically relevant inflammatory and airway-related variation beyond broad regional structure, though this remains correlational rather than causal evidence of mediation. Similarly, region-adjusted associations with ambient temperature implicated proteins with prior links to heat stress and heat acclimation biology (**table S4**). Notably, C1QTNF1, MOG, and SMOC1 overlap with proteins previously reported in human heat-acclimation studies (*17*). By contrast, canonical heat-shock chaperones HSPA1A and HSPB1 (*18–20*), which were significantly associated with temperature before regional adjustment, became insignificant after adjustment, suggesting that regional structure explains their apparent temperature association, while a distinct subset of heat-stress-related proteins remain robust to regional confounding (**Supplementary text**).

Second, we estimated group-stratified exposure-protein associations to test for heterogeneity across genetically inferred ancestry groups (EUR-like, AMR-like, AFR-like, EAS-like, and SAS-like) or self-reported race and/or ethnicity groups (White, Hispanic, Black, Asian, and Mixed) (**Fig. 1**). We focused primarily on stratifying by self-reported race and/or ethnicity, as environmental and social exposures are expected to be socially patterned, but retained genetically inferred ancestry for validating protein scores derived from ancestry-stratified discovery cohorts. Notably, as ancestry is a continuum, these groupings are artificial and should not be conflated with social constructs such as race and/or ethnicity; they are used to compare with existing studies (e.g. UKB). Overall, evidence of heterogeneity was limited: across more than 100 exposures, fewer than 1% of proteins per exposure showed significant heterogeneity by Cochran’s Q after Bonferroni-corrected meta-analysis, with no difference between ancestry-and race-and/or ethnicity-based stratification (median 0.06% for both; Wilcoxon P = 0.173) (**Fig. 2B, fig. S3 to S4, table S5**). The current smoking score showed the clearest pattern of ancestry-related variation, evident both across and within biobanks. The UKB-trained score, applied to AoU (**Fig. 2C, fig. S5 to S6, table S6**), performed best in EUR-like (r = 0.727) and worst in EAS-like (r = 0.417) participants, paralleling ancestry differences in smoking prevalence. Downsampling to match prevalence and cigarettes/day across groups narrowed this gap by ∼67% (EUR-like r: 0.727→ 0.528) (**table S7**). The AoU pooled-trained smoking score showed a similar pattern within AoU (**Fig. 2D, fig. S7 to S8, table S8**): performance ranged from r = 0.328 (EAS-like) to r = 0.721 (EUR-like) and was modestly stronger in lower-income and lower-education participants, paralleling roughly two-fold higher smoking prevalence and intensity in low-versus high-income groups. Downsampling non-EAS-like groups to match EAS-like smoking prevalence attenuated this gap by ∼77% (EUR-like: r = 0.722 → 0.434), reinforcing that ancestry differences in score performance–both within and across biobanks–largely reflect differences in exposure prevalence rather than reduced portability of biological signals.

The UKB-trained alcohol score also generalized to AoU, performing best in SAS-like (r = 0.300) and worst in EUR-like (r = 0.150); unlike smoking, this ordering did not consistently track alcohol-use prevalence (e.g., EAS-like performance paralleled its prevalence rank, while SAS-like did not). Among other AoU pooled-trained scores, cardiometabolic traits performed strongest and most stably (BMI r = 0.845 [0.834-0.856]; range 0.810-0.860 across ancestry), while environmental exposures (PM_2.5_, temperature) were weaker but reproducible (r = 0.193-0.321).

Third, we constructed multivariate protein scores to assess exposure specificity (**Fig. 1**, **Fig. 2E-G, table S9 to S11, fig. S9 to S10**), classifying each exposure-trained score as “on-target” (matched exposure had the highest R^2^), “off-target: within-class” or “off-target: between-class”. Trait specificity was high for anthropometry, blood pressure, lipid, liver, and kidney scores (100% on-target), moderate for ambient temperature, air pollution, and hematologic scores (75-89% on-target or within-class), and markedly weaker for SES and Tobacco/Substance scores, with only the deprivation index on-target among SES measures and just 2 of 34 Tobacco/Substance scores on-target (**table S9 to S10**). This hierarchy suggests that anthropometric and clinical biomarker scores behave as precise, exposure-specific signatures, whereas SES and substance use scores more often capture broader behavioral or systemic proteomic patterns. Because many exposures are phenotypically correlated (e.g. cigar smoking and marijuana use), some off-target associations could reflect that correlation rather than genuine cross-reactivity of the protein score itself. To distinguish these possibilities, we recalculated each score’s specificity margin after adjusting for the phenotypic correlation structure among exposures. Averaged across the 34 Tobacco/Substance scores, this adjustment attenuated the mean off-target margin (on-target - strongest off-target R^2^) from -0.11 to -0.06 and raised the number of on-target scores from 2 to 7 of 34 (**table S11, fig. S11**). The most pronounced case was the cigarette-per-day score, whose margin reversed sign after adjustment (-0.14 before to +0.23 after), such that it predicted its matched cigarette-per-day trait more strongly than any residual off-target association. These results indicate that shared behavioral structure among exposures explains some cross-prediction, but residual cross-reactivity beyond co-occurrence of exposures remains the norm rather than the exception for Tobacco/Substance scores.

### Proteomic signatures of air pollution reflect individual exposure as well as broader geographic and environmental structure

A key question in proteomic exposure modeling is whether protein scores reflect individual-level exposure variation or instead capture geographic and socioeconomic structure correlated with exposure. We addressed this directly for PM_2.5_ by evaluating the spatial distribution of the PM_2.5_ protein score, its specificity relative to related environmental and geographical scores, and its transferability across biobanks. To evaluate spatial concordance, we summarized residential PM_2.5_ estimates and PM_2.5_ protein scores to ZIP3 regions by population-density weighting and compared tertile assignments across 247 regions with complete data (**Fig. 3A**). Overall concordance between observed and predicted PM_2.5_ was 46.6%. Some local discordance was also evident (5.7% high observed/low protein score; 4.0% low observed/high protein score ZIPs), which may reflect residual geographic or socioeconomic confounding, exposure misclassification, temporal mismatch, or individual-level heterogeneity in exposure or biological response.

**Fig. 3.**
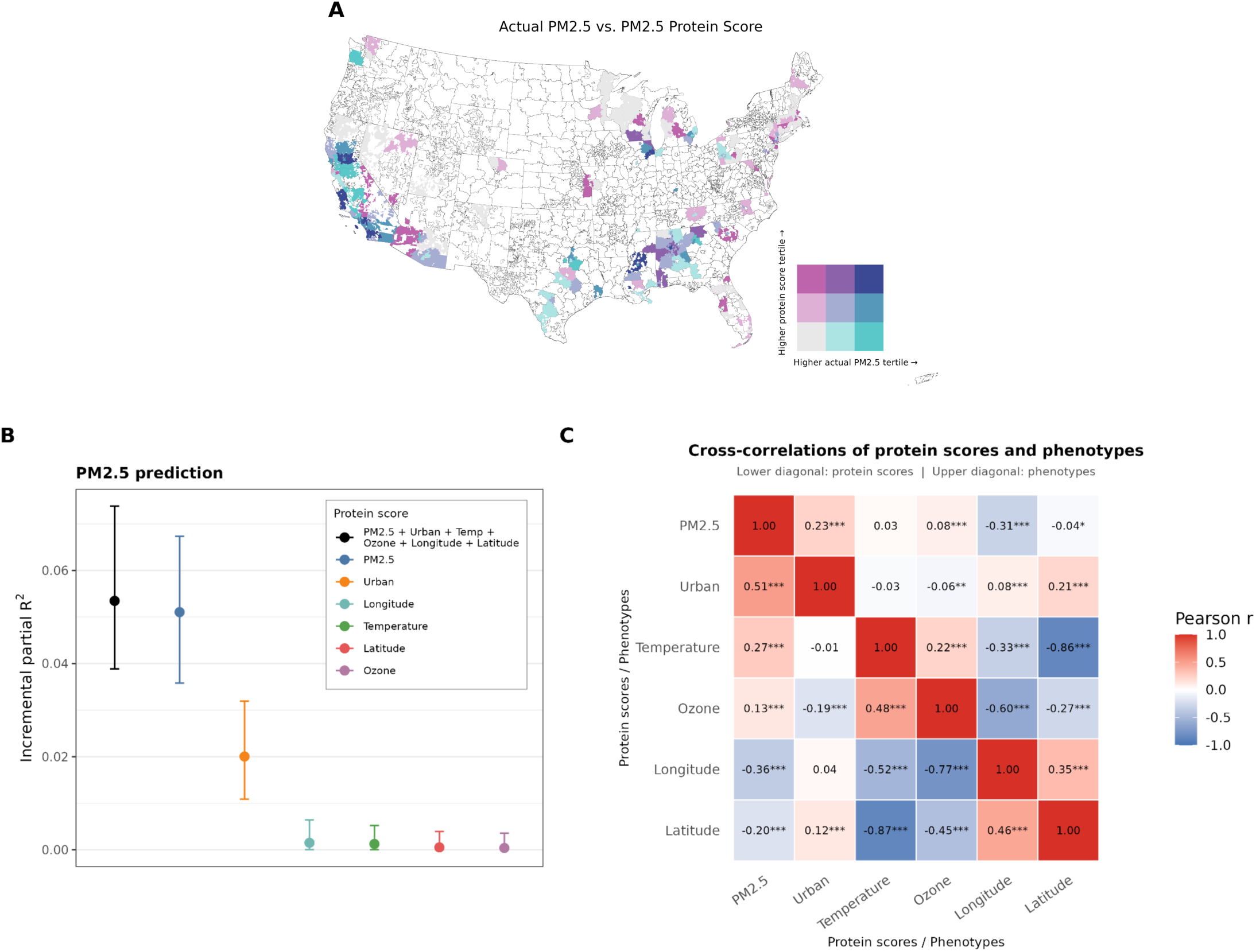
Proteomic validation of PM2.5 and urbanization exposure signatures. (**A**) Bivariate spatial map of observed annual PM2.5 and the PM2.5 protein score across ZIP3 regions. ZIP3s were classified into tertiles of observed PM2.5 and mean PM2.5 protein score among regions with complete data; outlined white regions indicate missing observed PM2.5 or protein-score data. (**B**) Incremental partial R² for the PM2.5 protein score in single-score models and in joint models including PM2.5, urbanization, ambient 365-day average temperature, 30-day average ozone, longitude, and latitude protein scores, after adjustment for age, sex, and biosample collection timing. (**C**) Correlation matrix among PM2.5, urbanization, ambient 365-day average temperature, 30-day average ozone, longitude, and latitude. Lower diagonal shows pairwise correlations between protein scores; upper diagonal shows pairwise phenotypic correlations between the corresponding observed exposures.

To assess specificity, we compared the PM_2.5_ protein scores against five additional protein scores — each trained in the held-out test set to predict a related environmental or geographic trait (urbanization, 365-day ambient temperature, 30-day ozone, longitude, and latitude) (**Fig. 3B-C, table S12**). In univariate models, the PM_2.5_ protein score was most strongly correlated with measured annual PM_2.5_ (R^2^ = 0.064) (**table S9**), with the urbanization score contributing modestly (R^2^= 0.015), and all protein scores were significantly associated with the PM_2.5_ protein score (**Fig. 3C**). In a joint model including all six scores, the PM_2.5_ score explained the largest independent fraction of PM_2.5_ variance (incremental partial R^2^ = 0.051 [0.035-0.069]), followed by a smaller contribution from the urbanization protein score (partial R^2^ = 0.020 [0.011-0.032]), and little additional information from temperature, ozone, longitude, and latitude scores (**Fig. 3B**). The full six-score model explained only modestly more variance than the PM_2.5_ alone (R^2^ = 0.053 [0.040-0.074]), indicating limited independent information from the remaining environmental and geographic scores. The higher accuracy of the PM_2.5_ score compared to all geographical and environmental correlates, indicates that it captures individual-level exposure signal beyond geographical structure alone. Nevertheless, the PM_2.5_ score was associated with a broader set of traits beyond air pollution, including inflammatory, metabolic, hematologic, renal, and hepatic markers, as well as smoking-related and socioeconomic variables (**table S9**). Rather than undermining specificity, this pattern suggests that protein scores for environmental exposures inherently encode the physiological consequences of the exposure as it occurs in the population, making them biological measures of real-world exposure patterns rather than isolated biomarkers of a single measured variable.

Cross-biobank transferability was limited, providing further evidence that PM_2.5_ protein signatures are partly geography-dependent. UKB-derived air pollution scores showed minimal predictive performance in AoU, with the strongest signal restricted to NO_2_ in AMR-like participants (incremental R^2^ = 0.050 [0.033-0.071], versus ≤0.016 in all other ancestry groups) (**fig. S5, table S6**). This association attenuated substantially after adjustment for geographic region (**fig. S12**), suggesting that it captured geographic structure rather than individual-level exposure variation alone, consistent with our within-AoU validation results. Differences in exposure definitions and temporal windows between UKB and AoU likely also contributed to this limited portability (**Supplementary text**).

### Proteomic exposure signatures reflect socially patterned metabolic context

#### Heterogeneity in BMI-associated proteomic signatures across population strata

We stratified exposure-protein association analyses across genetic ancestry and self-reported race and/or ethnicity groups to evaluate heterogeneity. Among all exposures examined, BMI showed the greatest degree of heterogeneity (**fig. S3**), with 111 (2.05%) and 116 (2.14%) proteins meeting our heterogeneity criteria (fixed-effect meta-analysis Bonferroni-corrected *P* < 0.05 and Cochran’s Q *P* < 1.66 × 10^-5^) across ancestry and race and/or ethnicity groups, respectively. Leptin was the most heterogeneous BMI-associated protein across ancestry (Cochran Q test p-value [p_het_]=1.38 × 10^-19^) and the fifth most heterogeneous across race and/or ethnicity groups (p_het_=1.97 × 10^-11^). BMI itself is highly heterogeneous across race and/or ethnicity groups, with the lowest mean and standard deviation in self-identified Asian participants (25.6 kg/m² [4.6]) to highest in self-identified Black participants (29.5 kg/m² [7.0]); comparable patterns were observed by computed genetic ancestry (**fig. S13, table S13**). This phenotypic heterogeneity extended to socioeconomic strata: BMI-protein score correlations were modestly higher in lower-SES groups (low-SES *r* = 0.850, low education *r* = 0.848;mean BMI = 29.1 kg/m² in both), than in higher-SES groups (high-income *r* = 0.829, high-education *r* = 0.824; mean BMI = 26.9 kg/m² and 26.5 kg/m², respectively). Deprivation gradients were less consistent, though correlation was still higher in the high-deprivation group (*r* = 0.864; mean BMI = 28.5 kg/m²) versus low-deprivation group (*r* = 0.847; mean BMI = 27.3 kg/m²) (**Fig. 4A**, **fig. S7, table S8**). These patterns indicate that BMI-associated protein signatures track socioeconomic exposure gradients, in addition to the variation observed across race and/or ethnicity categories, consistent with a socially patterned component to this heterogeneity. To assess whether cross-race and/or -ethnicity differences in BMI protein score performance reflected differing BMI distributions, we repeated the analysis after matching each group’s BMI distribution to that of self-identified Asian participants, who had the smallest mean BMI. BMI score correlations decreased for all groups after matching, narrowing the gap relative to the Asian reference by roughly half for self-identified Black and self-identified White participants (**fig. S14, Table S14**). Baseline T2D prevalence also differed by race and/or ethnicity, matching BMI distributions substantially narrowed this gap across groups, consistent with BMI as a substantial partial mediator of racial disparities in T2D prevalence. As discussed above with downsampling to match smoking status prevalence, protein score generalizability across race/ethnicity groups remains relatively high, though gaps persist that are likely attributable in part to differences in socioeconomic status. Consistent with this, numerous disparities by race/ethnicity are embedded directly in the EHR itself (**fig. S15**). For example, mean EHR completeness was highest among Black and White participants and markedly lower among Asian participants. Black and Hispanic participants additionally reported substantially higher rates of public versus private insurance and greater difficulty affording specialist care than White or Asian participants.

**Fig. 4.**
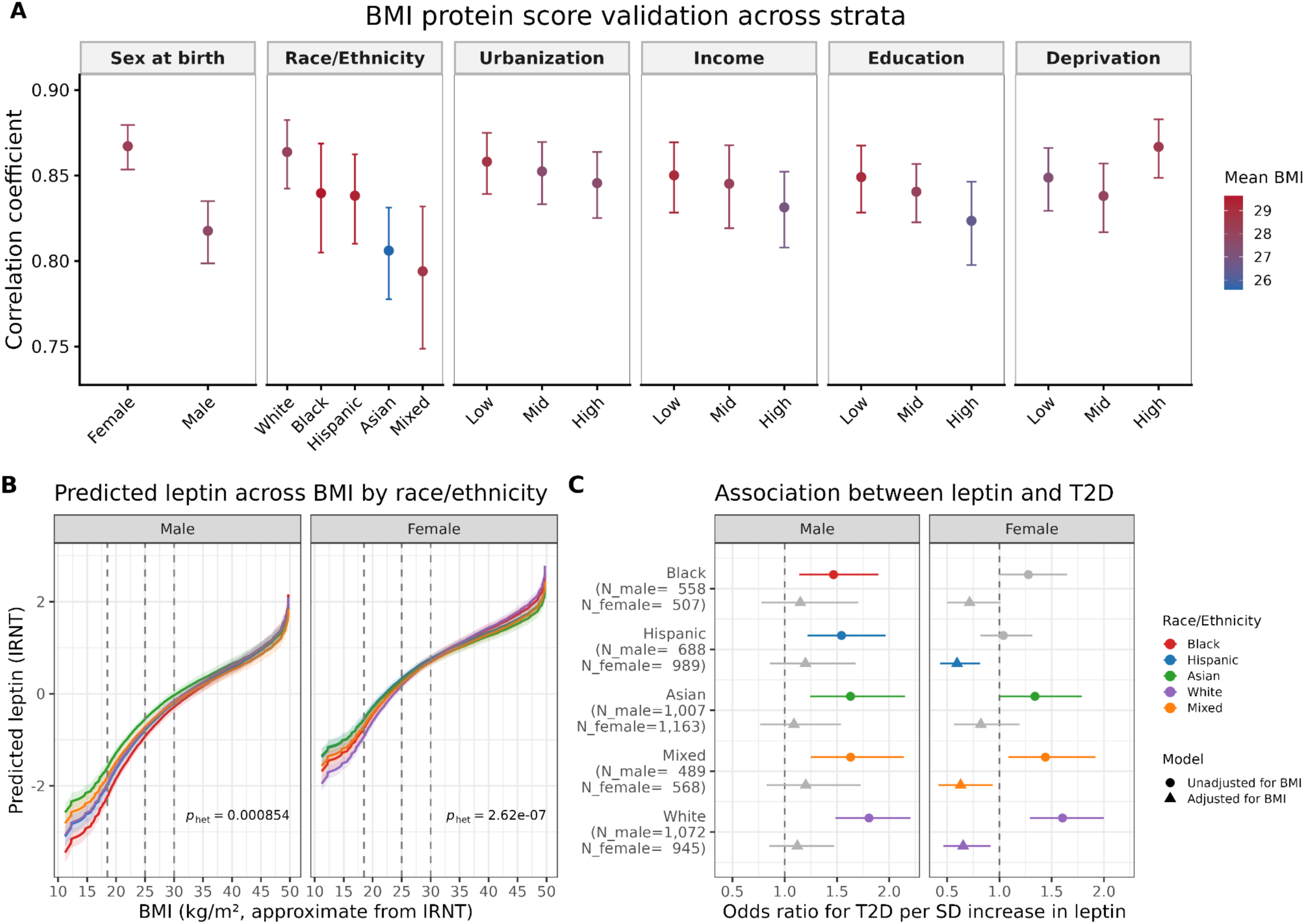
Cross-race and/or -ethnicity heterogeneity in BMI-based associations with leptin in male and female. (**A**) BMI protein score validation across AoU test set strata defined by sex, race and/or ethnicity, urbanization, income, education, and deprivation. (**B**) Predicted leptin levels across BMI by race and/or ethnicity, based on interaction models. Vertical dashed lines indicate conventional BMI thresholds (18.5, 25, and 30 kg/m²). (**C**) Association of leptin with T2D after BMI adjustment. Points show ancestry-specific estimates for the association of leptin with T2D before (circles) and after BMI adjustment (triangles). N shown on the y-axis corresponds to the BMI-adjusted model.

The magnitude and ranking of BMI-leptin associations was sensitive to the BMI transformation applied (raw values, z scores, and inverse rank normal transformation (IRNT), underscoring the need for caution when interpreting relative effect sizes across populations (**fig. S13, Supplementary text**). BMI was positively associated with leptin in all race and/or ethnicity groups, with substantial heterogeneity in effect sizes (p_het_ = 1.97 × 10^-11^), strongest in self-identified Black and self-identified White individuals and attenuated in self-identified Asian participants, with comparable patterns by genetic ancestry (**Fig. 4B**, **fig. S13**). This heterogeneity was more pronounced in females than males (p_het_ = 2.62 × 10^-7^ for female and 8.54 × 10^-4^ for male), with comparable sex-stratified patterns across race and/or ethnicity and genetic ancestry groups.

We next assessed whether leptin contributes to T2D risk independently of BMI (**Fig. 4E, table S15**). In males, leptin was positively associated with T2D risk across all race and/or ethnicity groups prior to BMI adjustment, but these associations were fully attenuated toward the null after adjustment, indicating that leptin provides no information about T2D risk beyond BMI in men. In females, the pattern was more complex: leptin was associated with T2D risk in the self-identified White, Mixed, and Asian groups before adjustment, but BMI adjustment unmasked a significant inverse association between leptin and T2D risk in most race and/or ethnicity groups, suggesting that leptin may mark a protective process once overall adiposity is accounted for — consistent with a previously reported sex-differential pattern of sign-reversal after adiposity adjustment (*21*) (**table S15**, **fig. S16**). These findings indicate that the leptin-T2D association is largely a proxy for adiposity as captured by BMI, with a possible protective signal in females that warrants further investigation.

### Proteomic signatures distinguish related behavioral exposures

Cigarette smoking and marijuana use were highly prevalent in the AoU proteomic cohort, with substantial heterogeneity across demographic and socioeconomic strata (**fig. S17 to S21, table S16 to S17, and Supplementary text**). Overall, 47.3% (3,979/8,418) of participants reported ever using marijuana and 34.8% (2,926/8,418) reported ever smoking cigarettes, with substantial co-use: 70.9% of smokers also used marijuana, while 47.9% of marijuana users also smoked cigarettes (**Fig. 5A**, **fig. S22**). Substance use prevalence varied by age, sex, race and/or ethnicity, socioeconomic status, and geography, consistent with established U.S. social patterning (*22*) and differences in regulatory environments (*23*, *24*).

**Fig. 5.**
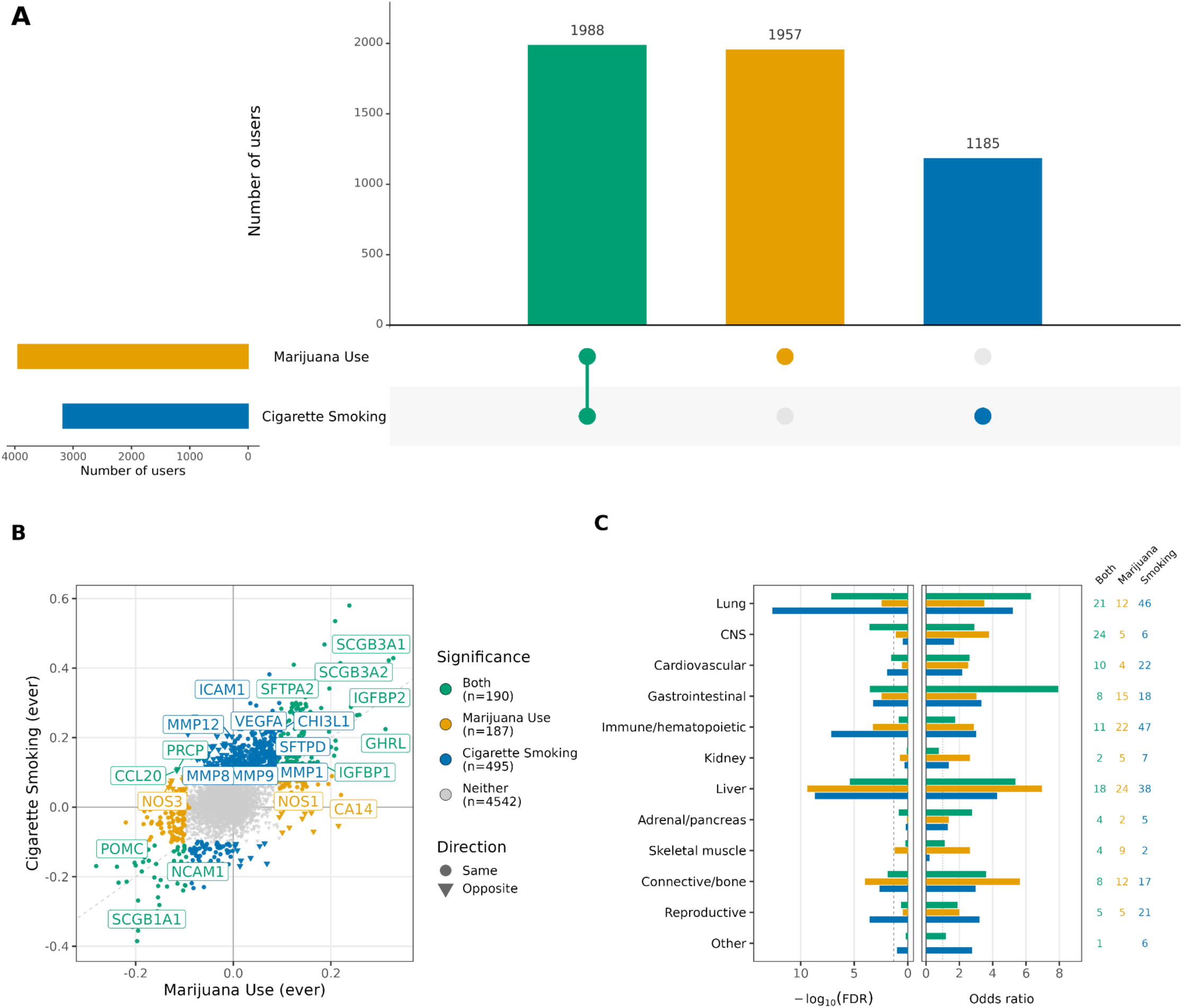
Marijuana use and cigarette smoking share an inhalation-associated proteomic signature but differ in exposure-specific peripheral effects. (**A**) UpSet plot illustrating co-use patterns between marijuana ever use and cigarette ever smoking in AoU proteomics set. (**B**) Comparison of beta coefficients in linear regression of ever marijuana use (x axis) and cigarette smoking (y axis) for top-associated proteins (P_bonf_<0.05) from marijuana use (377 proteins) and cigarette smoking (685 proteins). Proteins are colored by classification: cigarette smoking-predominant effects (blue), marijuana use-predominant effects (yellow), and effects shared between both exposures (green) (**table S17**). Gray points indicate proteins not significantly associated with either exposure (P_bonf_ ≥ 0.05). (**C**) Tissue enrichment of the three protein sets from panel B (cigarette smoking-only, marijuana use-only, and shared) relative to proteins associated with neither exposure. For each tissue, proteins were classified as tissue-elevated or not, and each target group was compared against the neither-exposure group using one-sided Fisher’s exact tests. P-values were adjusted using the Benjamini-Hochberg procedure within each comparison across tissues (**table S18**).

#### Shared and distinct proteomic associations of marijuana use and cigarette smoking

At the individual protein level, marijuana use and cigarette smoking shared 190 Bonferroni-significant proteins (**Fig. 5B, table S18**), with the strongest tissue enrichment in the lung (-log_10_FDR = 7.14, OR = 6.3) (**Fig. 5C, table S19**) across trait definitions (**fig. S23 to S24**), supporting a shared physiological response to inhalational exposure. Several shared proteins have prior biological and genetic support linking them to smoking-related or neurobehavioral processes. Shared proteins included lung-enriched secretory proteins (SCGB1A1, SCGB3A1/SCGB3A2, SFTPA2) with established roles in airway epithelial function and known alteration by cigarette smoke (*25*, *26*). NCAM1 was also strongly associated with both exposures (**table S4**); rs7935745 in *NCAM1* is a well-replicated locus for smoking initiation, nicotine dependence, and smoking motives (*27–29*), and NCAM1 has an established role in synaptic plasticity (*30–32*). Additional shared proteins enriched in liver, gastrointestinal, and central nervous system (CNS) tissues included metabolic and neuroendocrine regulators (GHRL, POMC, IGFBP1, IGFBP2) (**table S17 to S18**), suggesting a neuroendocrine axis linking central appetite regulation with peripheral metabolic signaling (*33*, *34*), consistent with prior experimental evidence (*35–37*). We note that tissue enrichment does not necessarily correspond to the tissues most highly represented or most abundantly measured on the Olink panel itself (**Supplementary text**). A subset of shared proteins showed opposing directions of effect between marijuana use and cigarette smoking, most notably the nitric oxide synthase NOS3 (eNOS), which was inversely associated with marijuana use but positively associated with current cigarette smoking (**table S18, fig. S25, Supplementary text**), consistent with distinct, well-characterized effects of cigarette smoke versus cannabinoids on nitric oxide signaling pathways (*38–42*).

Proteins significant only for cigarette smoking (n=495) were enriched for pathways related to lung injury, extracellular matrix remodeling, and inflammation, including matrix metalloproteinases (MMP1, MMP8, MMP9, MMP12), vascular and endothelial markers (ICAM1, VEGFA), and surfactant-related proteins (SFTPD), consistent with established smoking-related pulmonary pathology (*43–45*). CHI3L1 (YKL-40), a well-established biomarker of inflammation and tissue remodeling, elevated in smokers and associated with chronic lung disease (*46*, *47*) was also among these (**table S18**). Indeed these cigarette smoking proteins are highly enriched in the lung tissue (-log_10_FDR = 12.65, OR = 5.20) (**Fig. 5C**), consistent across alternative trait definitions (**table S19, fig. S23 to S24**).

Fewer proteins were uniquely associated with marijuana use (n=187) than with cigarette smoking, despite comparable statistical power in this analysis, suggesting a narrower proteomic signature for marijuana use rather than a detection-power limitation. These proteins were most enriched in liver, connective/bone and immune/hematopoietic tissues, with a small subset (including CA14, previously implicated in schizophrenia (*48*)) also enriched in CNS regions (**Fig. 5C**, **table S19**). Among marijuana-specific proteins, NOS1 (nNOS) showed a pattern consistent with direct cannabinoid biology, given the established role of CB1 receptor activation in modulating nNOS-dependent nitric oxide signaling (*49*, *50*) (**Supplement text**).

SES-stratified analyses (income, deprivation, education), which showed a larger and more strongly lung-enriched shared proteomic signature in lower-SES groups—potentially reflecting greater co-use prevalence and cumulative exposure (**fig. S26 to S28, table S20 to S22, Supplementary text**).

Comparing these proteomic findings against published GWAS of smoking initiation (*27*) and cannabis use (*51*), together with tissue and cell-type enrichment analyses (**fig. S29 to S30, table S23 to S24, Supplementary text**), genetic susceptibility was most strongly linked to neuronal loci (e.g., *CADM2, NCAM1*) and cell types, whereas proteomic signatures predominantly reflected peripheral physiological responses—airway epithelial injury, systemic metabolic regulation, and immune signaling. These results support a model in which genetic variation primarily shapes susceptibility to substance-use initiation, while the circulating proteome captures the integrated physiological consequences of exposure across organ systems.

## Discussion

Plasma proteins could provide an objective molecular record of environmental exposure, but their value depends on three properties: consistency across populations, specificity across exposures, and reproducibility across cohorts. We tested all three in 9,561 All of Us participants profiled for 108 environmental, behavioral, socioeconomic, anthropometric, and clinical exposures. Associations were largely shared across ancestry and race and/or ethnicity strata, protein scores retained specificity across major exposure classes, and scores trained in UK Biobank transferred to All of Us for biologically proximal exposures. Yet score performance was not uniformly portable–evaluating generalizability requires joint consideration of biological consistency alongside exposure prevalence, social context, and cohort structure.

Fewer than 1% of proteins per exposure showed significant heterogeneity across race and/or ethnicity and ancestry groups, suggesting that many proteomic responses to environmental and behavioral exposures are broadly shared. Yet, portability still requires calibration across race and/or ethnicity, sex, age, and social context, echoing lessons from polygenic scores (*52*, *53*). BMI was the clearest exception, showing the greatest heterogeneity of any exposure tested. Leptin was most heterogeneous across ancestries, with trajectories diverging nonlinearly across ancestry, race and/or ethnicity, and sex, and sensitive to how BMI was scaled.

Type 2 diabetes analyses reinforced the pattern: BMI-associated diabetes risk differed across ancestry and race and/or ethnicity, whereas leptin’s association was largely BMI-dependent, with residual sex-specific effect. We interpret BMI-linked proteomic signals as indexing socially and biologically patterned metabolic context rather than adiposity alone. We stress that race and/or ethnicity is not interchangeable with ancestry and that both are useful: ancestry is a continuum that approximates population genomic structure, whereas self-identified race and/or ethnicity captures social and structural processes that genetics does not.

An important caveat for our many cross-ancestry comparisons is Jensen’s fallacy: the mistaken inference that because a trait is heritable within a population, differences in that trait between populations must also be genetic (*54*). Our own estimates illustrate why this distinction matters: SNP-heritability for height ranged from *h^2^* = 0.395 in the AMR-like group to *h^2^* = 0.602 in the EAS-like group, yet this within-population heritability does not indicate that the corresponding between-population difference in height, or in the protein signatures we study here, have a genetic versus environmental or cultural basis (*55*). The same caution applies to our broader cross-ancestry and cross-race and/or -ethnicity comparisons of exposure-protein associations.

At the individual-protein level, many exposures shared associated proteins, consistent with pleiotropic inflammatory, metabolic, renal, hematologic, and airway responses. At the aggregated level, however, protein scores were largely exposure-specific across anthropometry, blood pressure, lipid, liver, and kidney domains. Specificity broke down where exposures are themselves entangled: tobacco and substance-use largely cross-predicted one another, reflecting real co-use patterns and shared inhalational biology. The PM_2.5_ score captured within-region variation and outperformed related environmental and geographic scores, yet also correlated with urbanization, socioeconomic status, inflammation, and smoking–reflecting challenges ahead for disentangling effects of pollution, deprivation, healthcare access, and comorbid disease risk that characterize environmental health disparities.

Cross-biobank transfer was strong for exposures with direct, repeated physiological effects. UKB-trained scores for current smoking and alcohol use transferred cleanly to AoU across genetic ancestry groups, suggesting that reproducible biological responses extend beyond the cohort in which they were developed. Air-pollution scores showed more limited transferability, likely reflecting cohort differences in geography, spatial resolution, and exposure modeling rather than a fundamental biological limit. These results highlight a distinctive contribution of AoU: its diversity across genetic ancestry, self-identified race and/or ethnicity, socioeconomic context, and geography provides a critical test of whether molecular exposure signatures generalize across groups historically underrepresented in biomedical research. Large resources such as deCODE, Fenland, UK Biobank, and FinnGen have transformed population proteomics (*6*, *10*, *11*, *56*), but are comparatively homogeneous in ancestral, social, and healthcare-system context. AoU therefore provides a complementary setting for distinguishing shared biological response from context-specific exposure histories.

Several limitations apply. Most analyses were cross-sectional, limiting temporal and causal inference. Protein signatures may reflect exposure effects, correlated exposures, compensatory physiology, subclinical disease, treatment, or reverse causation. Exposure phenotypes derived from surveys, EHRs, and geocoded residential data are subject to measurement error, recall bias, temporal mismatch, and spatial misclassification. We are largely missing individual mobility, indoor exposures, occupational exposures, or historical residential trajectory phenotypes. Results may also differ across tissues, assay platforms, longitudinal sampling designs, and disease-enriched cohorts.

The most important next step is moving beyond association. A rapidly growing literature linking circulating proteins to disease risk (*2*, *3*, *5*), and a smaller set of studies has begun building proteomic exposure biomarkers (*1*, *6*, *7*). The 108 exposures examined here provide initial inputs for that framework. Extending proteomic exposure profiling to a broader set of environmental, occupational, and infectious exposures, including viral and other pathogen exposure histories, which may carry long-term relevance for chronic disease risk, could substantially expand the utility of this approach. Calibrating them to the social, epidemiologic, and biobank contexts in which they are applied will be essential to using them responsibly–such as disentangling how much of an observed disparity reflects differential exposure burden versus differential access to care– rather than reproducing existing measurement inequities.

In conclusion, plasma proteomic signatures capture a broad range of environmental and behavioral exposures with generally limited cross-group heterogeneity, substantial exposure class specificity, and transferability for strong and biologically proximal exposure signatures. These findings establish proteomic exposure scores as a promising tool for exposure measurement, biobank harmonization, and health disparities research, while underscoring that their responsible use will require careful calibration and causal framework moving forward.

## Data Availability

This study used data from the All of Us Research Program Controlled Tier Dataset version 8 for all genetic and phenotype analyses. These version 8 data were paired with proteomic data released in version 9, with the analytic sample defined based on participants available in version 8. These data are available to authorized users on the Researcher Workbench. Code developed for this project in AoU is available as a featured workspace (https://www.researchallofus.org/research-project-directory/?snapshot=105587).

## Acknowledgements

We gratefully acknowledge *All of Us* participants for their contributions, without whom this research would not have been possible. We also thank the National Institutes of Health’s *All of Us* Research Program for making available the participant data, samples, and cohort examined in this study.

## Data and Code Availability

This study used data from the *All of Us* Research Program’s Controlled Tier Dataset version 8 for all genetic and phenotype analyses. These version 8 data were paired with proteomic data released in version 9, with the analytic sample defined based on participants available in version 8. These data are available to authorized users on the Researcher Workbench. Code developed for this project in AoU is available as a featured workspace (https://www.researchallofus.org/research-project-directory/?snapshot=105587).

## Methods

### All of Us Data

The All of Us Research Program (AoU), launched by National Institute Health in May 2018, represents a longitudinal cohort study with the goal of engaging at least 1 million participants encompassing diverse ancestral backgrounds. By leveraging comprehensive data collection including biospecimens, health questionnaires, electronic health records and physical measurements, AoU aims to advance precision medicine and enhance overall human health (*57*). Participants, aged 18 years and older, are recruited from over 340 centers with informed consent. As of February 2025, a subset of around 414,830 participants has undergone whole genome sequencing (WGS).

We used AoU plasma proteomics for this study. The AoU proteomics cohort measured 5,416 proteins across 10,170 individuals who were selected to maximize ancestral diversity using the Olink Explore 5K platform (AoU CDEv9, 2026). The proteomic data were released in version 9 and were paired with genetic and phenotypic data from version 8 for the present analyses(*59*). We used All of Us (AoU)-provided genetic ancestry classifications, which are based on principal components (PCs) computed from WGS data and describe genetic similarity to reference populations from the 1000 Genomes Project and Human Genome Diversity Project (1KGP-HGDP)(*58*). We refer to these groups as African (AFR-like), Americas (AMR-like), East Asian (EAS-like), European (EUR-like), Middle Eastern (MID-like), and South Asian (SAS-like); individuals not classified into one of these six groups, or reflecting admixture, are designated as remaining (REM) by AoU. Our sample included 1,865 AFR-like, 2,034 AMR-like, 1,382 SAS-like, 1,308 EAS-like, 2,999 EUR-like and 406 MID-like individuals. Self-identified race and/or ethnicity was assessed using AoU’s "Race: What Race Ethnicity" survey variable (concept ID 1586140), a multi-select item from AoU’s Basics survey with response options Asian, Black, Hispanic, MENA (Middle Eastern or North African), NHPI (Native Hawaiian or Pacific Islander), White, and "None of these." Participants selecting exactly one of White, Black, Asian, or Hispanic were assigned to that category; participants selecting MENA, NHPI, or "None of these" (alone or in combination) were grouped as Other; and participants selecting more than one race and/or ethnicity category were classified as Mixed. Consequently, our Hispanic category reflects only participants who identified as Hispanic without concurrently selecting an additional race and/or ethnicity category; participants identifying as Hispanic in combination with another category were classified as Mixed. Individuals were assigned to biological sex (male or female) using the AoU’s “Sex at Birth” variable (Concept ID: 1585845). For proteomics, normalized protein expression (NPX) values were obtained from AoU following quality control and normalization procedures(*59*).

We used environment measurements from the AoU from individual-level survey data. The measurements include lifestyle factors such as alcohol consumption and smoking usage. We incorporated geocoded data of multiple socioeconomic indices, air pollution, and ambient temperature, which aggregate local income, employment, education, and housing to encompass potential latent features of lifestyle.

### Preprocessing exposure data

We evaluated a broad set of health-related exposures and traits encompassing behavioral, socioeconomic, environmental, anthropometric, clinical, and biomarker domains. This exposure framework reflects domains commonly evaluated in prior social and environmental determinants of health and exposomic studies (*1*), expanded to incorporate additional environmental variables available through AoU’s geocoded residential data (air pollution, ambient temperature, and neighborhood-level socioeconomic indices). The socioeconomic measures, including the deprivation index, education, income, were obtained from the socioeconomic status data available within the Controlled Tier dataset, which are externally sourced from the U.S. Census American Community Survey. For brevity, these variables are collectively referred to as “exposures” throughout the manuscript, although several traits, including anthropometric measures and clinical biomarkers, are influenced by both environmental and genetic factors. The final set comprised 108 exposure variables spanning tobacco and substance use (n=50, representing 15 distinct tobacco, alcohol, and substance-use constructs), anthropometry (n=3), blood pressure (n=2), socioeconomic characteristics (n=4), ambient temperature (n=4), air pollution (n =8, representing 6 distinct pollutants), hematologic biomarkers (n=18), glycemic biomarkers (n=3), lipid biomarkers (n=4), liver biomarkers (n=6), lung (n=1), thyroid (n=3), and kidney biomarkers (n=2) (**table S2**). The number of exposure variables exceeds the number of underlying exposure constructs because several domains were represented by multiple related measures, including lifetime-use, current-use, and exclusive-use tobacco and substance phenotypes, as well as short-term (30-day) and long-term (365-day) environmental exposure averages.

Exposure variables were assembled from multiple AoU data resources. Tobacco, alcohol, and recreational drug use phenotypes were derived from responses to the Lifestyle survey and included lifetime (“ever”), current-use, and exclusive-use measures. Anthropometric traits, blood pressure measurements, and clinical biomarkers were obtained from the All of Us electronic health record (EHR) and Labs & Measurements data resources. Selection of clinical and biomarker traits was informed by the exposure framework used by Carrasco-Zanini et al.(*1*) for proteome-wide association analyses and corresponding variables were identified and curated from available All of Us survey, measurement, and EHR resources. Socioeconomic and contextual measures were derived from geocoded neighborhood-level datasets linked to participants’ residential locations.

Binary exposures were analyzed as indicator variables. Continuous exposures were screened for extreme values, with observations exceeding 6 median absolute deviations from the median excluded prior to analysis and were subsequently inverse rank-normal transformed (IRNT). Sample size varied across exposures according to data availability, and analyses were conducted using all non-missing observations for each exposure. Detailed descriptions of exposure definitions, data sources, completeness, and distributions are provided in **table S2**.

Environmental exposures were assigned at the 3-digit ZIP code (ZIP3) level using a population-weighted aggregation of census tract-level information. ZIP3 geographies were constructed by dissolving 2010 ZIP Code Tabulation Areas (ZCTAs) to their three-digit prefix. Population weights linking tracts to ZIP3 areas were derived using the 2010 Census ZCTA-tract relationship file, which provides population counts for the intersection between each census tract and ZCTA. For each tract *t* and ZIP3 region *j*, weights were calculated as the proportion of the ZIP3 population residing within the tract-ZIP3 intersection and normalized such that the weights summed to one within each ZIP3 region. Detailed descriptions of the ambient temperature, air pollution, neighborhood socioeconomic datasets, and binary exposure definitions are provided in the **Materials and Methods**.

### Preprocessing plasma proteins

We used NPX values that accounted for batch effects and other technical covariates. Detailed descriptions of quality control procedures and the assessment of assay repeatability are provided in AoU, 2026(*59*). Samples with missing measurements for any assay, those identified as outliers using connectivity scores, and those did not have WGS were excluded. At the assay level, duplicated assays were excluded. Finally, protein expression values were transformed using IRNT prior to downstream analyses. 9,561 x 5,414 complete set.

### Individual Exposure-protein association analyses

We first evaluated associations between environmental exposures and individual plasma proteins to characterize exposure-related proteomic signals and assess their consistency across population groups. For each exposure, we performed univariate linear regression analyses in which each normalized plasma protein level (P) was regressed on the exposure of interest (E) and a set of covariates (C): *P_i_* = *β*_0_ + *β*_1_*E_i_* + *γ^T^C_i_* + *ε_i_*. All models were adjusted for age at blood collection, sex, calendar year, month, and time of blood draw, based on prior evidence that these factors influence protein expression variability. For exposures derived from geocoded data, we additionally adjusted for geographic region (NorthEast/MidWest/Western/Southern) based on state of residence to account for broad spatial variation. Analyses were conducted both in the full pooled sample and within strata defined by population groups.

To assess consistency across groups, we compared exposure-protein regression coefficients (*β*_1_) from group-stratified models using heterogeneity tests. Fixed-effects meta-analysis was performed across the group strata. Meta-analysis was conducted only for exposure-protein associations with valid regression estimates in at least two group strata. Associations were classified as heterogeneous if they were significant in the fixed-effects meta-analysis (Bonferroni-corrected p-value <0.05) and exhibited significant between-group heterogeneity according to Cochran’s Q test (p-value < 1.66 × 10⁻⁵, corresponding to 0.05 divided by 3,011 estimated independent proteins). The number of independent proteins was estimated using maximal independent set pruning of a protein correlation graph, where proteins were connected if their pairwise correlation exceeded r^2^ = 0.1.

### Aggregate Exposure protein score construction and evaluation

To assess the aggregate predictive capacity of proteomic data, we constructed protein-based exposure scores using multivariate models. The dataset was randomly partitioned into training (70%) and test (30%) sets, with stratification by population group to preserve group representation in both subsets.

For each exposure, we trained penalized regression models in the training set to predict exposure levels from the full set of proteins: 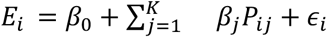 We used Lasso regression for variable selection and shrinkage, with hyperparameters tuned via cross-validation. Models were trained using pooled models including all individuals (10-fold cross-validation) and group-stratified models trained separately within each population group (5-fold cross-validation). The resulting models generated exposure-specific protein scores as weighted linear combinations of protein levels. For each individual i, the protein score was defined as: 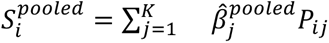. Protein-based exposure scores were evaluated in the independent test set. For each exposure, we assessed how well the proteomic scores predicted observed exposure levels under both pooled and group-stratified testing frameworks.

For continuous exposures, we fit linear regression models: *E_i_* = *α*_0_ + *α*_1_ *S_i_* + *ε_i_* and quantified predictive performance using the coefficient of determination (R^2^). For binary or categorical exposures, we fit logistic regression models: *logit*[*Pr*(*E_i_* = 1)] = *α*_0_ + *α*_1_ *S_i_* + *ε_i_* and evaluated performance using the area under the receiver operating characteristic curve (AUC). In addition, to evaluate the specificity of protein scores, we performed cross-exposure prediction analyses in which each exposure-specific protein score was used to predict other exposures.

To assess the extent to which protein-based predictions reflected exposure-related biology rather than confounding factors, we conducted sequential covariate adjustment analyses in the test set. For each exposure and group, we fit models of the form: *E_i_* = *α*_0_ + *α*_1_ *S_i_* + *γ^T^C_i_* + *ε_i_*, where *C_i_* included the same covariates used in the exposure-protein association analyses. We compared prediction performance before and after covariate adjustment to quantify attenuation in model performance attributable to confounding.

### Cross score-exposure correlations

To evaluate cross-exposure prediction and exposure specificity, each exposure-trained protein score was assessed against all exposure targets in the held-out test set. For binary exposure targets, we calculated the point-biserial correlation between the continuous protein score and the 0/1-coded exposure indicator. This is equivalent to the Pearson correlation between a continuous variable and a binary variable coded as 0 and 1. For continuous exposure targets, we calculated the Pearson correlation between the protein score and the continuous exposure value. For all score-target pairs, we report both the signed correlation coefficient, r, and the squared correlation, R^2^, to summarize directionality and magnitude, respectively. Pairwise correlations were computed using complete observations for the corresponding score-target pair.

To quantify exposure specificity, we compared the matched, diagonal score-exposure R^2^ with the maximum off-diagonal R^2^ across all non-matched exposure targets. Scores were classified as “on-target” if the matched exposure had a higher R^2^ than all off-diagonal targets. Scores for which an off-diagonal target exceeded the matched exposure were classified as “within-class” if the strongest off-diagonal target belonged to the same exposure class, or “between-class” if the strongest off-diagonal target belonged to a different exposure class. For phenotypic exposure-exposure correlations, binary-binary pairs were evaluated as Pearson correlations between two 0/1-coded variables, equivalent to the phi coefficient; binary-continuous pairs were evaluated using point-biserial correlations; and continuous-continuous pairs were evaluated using Pearson correlations. As a sensitivity analysis, we modeled off-diagonal score-exposure R^2^ as a function of phenotypic exposure-exposure R^2^ across all off-diagonal pairs. Residual cross-prediction was defined as the observed off-diagonal score-exposure R^2^ minus the value expected from phenotypic R^2^. We then compared each score’s matched, diagonal R^2^ with its maximum residual off-diagonal cross-prediction to assess whether exposure specificity persisted after accounting for phenotypic correlation among exposures.

### Evaluation of UK Biobank-trained protein-exposure scores in All of Us

We evaluated the transferability of UK Biobank-trained protein-exposure scores in the All of Us Research Program. We focused on externally trained LASSO-based protein scores for current smoking and current alcohol drinking. For each exposure, external UKB score weights and model intercepts were applied to normalized AoU proteomic measurements using proteins that overlapped between the UKB score model and the AoU proteomic platform. The protein-exposure score for each participant was calculated as the sum of the model intercept and the weighted protein abundances, using the published UKB LASSO coefficient for each overlapping protein(*7*). AoU exposure phenotypes were harmonized to the corresponding UKB exposure definitions. Current smoking was evaluated using the binary AoU current smoking phenotype. Current alcohol drinking was defined using AoU alcohol questionnaire data, where users were participants who reported lifetime alcohol participation and non-never drinking in the past year, and non-users were participants who reported no lifetime alcohol participation. Participants with missing or ambiguous alcohol-drinking status were excluded from the corresponding analysis.

Score performance was evaluated in the full proteomics AoU analysis set and stratified by genetically inferred ancestry. Analyses were performed separately in EUR-like, AMR-like, AFR-like, EAS-like, and SAS-like groups. For each exposure and ancestry group, discrimination was quantified using the AUCs, with 95% confidence intervals estimated using DeLong’s method. AUCs were calculated using the external UKB protein-exposure score as the predictor and the harmonized AoU exposure phenotype as the binary outcome, adjusting for age, sex, collection year, month of sample collection, and time of day of sample collection. Full descriptions of the prevalence-matched sensitivity analysis are provided in the **Materials and Methods**.

### Tissue enrichment for Proteomic signatures of behavioral exposures

We performed GTEx-based tissue enrichment analysis using GTEx v10 median gene expression values. The GTEx v10 gene median TPM matrix was read into R and converted to a SummarizedExperiment object with genes as rows and tissues as columns. Gene symbols from the GTEx Description column were used to match Olink protein identifiers. Rows with missing or invalid gene identifiers were removed, and duplicate gene symbols were collapsed by retaining the maximum median TPM across duplicate rows for each tissue.

Tissue-elevated gene sets were defined using the teGeneRetrieval() function from the TissueEnrich R package(*60*), which applies a Human Protein Atlas-style tissue-specificity classification to expression matrices. We used a four-fold expression threshold, a maximum of five tissues for group-enriched genes, and a minimum expression threshold of 1 TPM. Genes classified as Tissue-Enriched, Group-Enriched, or Tissue-Enhanced were considered tissue-elevated. Non-tissue-elevated categories, including Expressed-In-All, Mixed, and Not-Expressed, were not used as tissue-specific sets.

For each pair of traits, proteins were classified into four mutually exclusive groups: associated with both traits, associated with the first trait only, associated with the second trait only, or associated with neither trait. Gene symbols from the Olink protein panel were matched to the GTEx/TissueEnrich reference gene set, and proteins without a matching GTEx annotation were excluded. Overall, 5,331 of 5,414 (98.5%) unique Olink gene symbols were represented in the GTEx/TissueEnrich reference. For each tissue, we compared each target group (Both, trait 1 only, and trait 2 only) against the Neither group – comprising all remaining Olink panel proteins not significantly associated with either trait – using one-sided Fisher’s exact tests. For each tissue, proteins were classified as tissue-elevated or not tissue-elevated according to the GTEx/TissueEnrich tissue-specific gene sets. Genes represented in the GTEx/TissueEnrich reference but not classified as tissue-elevated for a given tissue remained in the denominator as non-tissue-elevated genes. To assess whether tissue-enrichment results reflected true biological enrichment rather than differential detectability on the Olink panel, we tested whether liver tissue-elevated status predicted significant associations with marijuana use or cigarette smoking, independent of protein abundance (**Supplementary text**).

For each test, we reported the odds ratio, p-value, target-group and comparator-group denominators, and the numbers of tissue-elevated proteins in each group. P-values were adjusted using the Benjamini-Hochberg false discovery rate (FDR) procedure both within each trait-pair/comparison across tissues and globally across all tissue enrichment tests.

### GWAS validation for Proteomic signatures of behavioral exposures

To provide genetic context for the proteomic findings, we examined publicly available genome-wide association study (GWAS) summary statistics for cigarette smoking initiation and lifetime cannabis use. Summary statistics for smoking initiation were obtained from the GWAS meta-analysis reported by Saunders et al. (2022)(*27*), comprising individuals of European ancestry (n = 2,669,029). Summary statistics for lifetime cannabis use were obtained from the Bright et al. (2026)(*51*) European-ancestry GWAS (total effective sample size: 258,823). Only variants present in both datasets were retained for downstream comparisons. To facilitate comparison between traits, effect estimates were harmonized to the same effect allele, and genome-wide significant loci were annotated to the nearest protein-coding gene using standard genomic annotation.

To characterize the tissues and cell types implicated by genetic susceptibility to smoking initiation and lifetime cannabis use, we performed gene-property and cell-type enrichment analyses using FUMA v1.6.x (Functional Mapping and Annotation of GWAS(*61*)). Independent genome-wide significant variants (P < 5 × 10⁻⁸) were mapped to genes using the default FUMA SNP2GENE pipeline. Tissue-specific expression enrichment was evaluated using GTEx (v8), and cell-type specificity analyses were performed using the precomputed single-cell RNA sequencing reference datasets implemented in FUMA (including Tabula Muris FACS). Enrichment significance was assessed using the default MAGMA gene-property framework implemented in FUMA, and tissues or cell types with FDR-adjusted P < 0.05 were considered significantly enriched.

### SNP-heritability estimation

We estimated SNP-heritability (*h^2^*) for 108 exposure and biomarker traits across five ancestry groups in the All of Us v8 cohort (AFR-like N=74,575; AMR-like N=70,626; EAS-like N=9,706; EUR-like N=75,000; SAS-like N=5,368) using GCTA-GREML(*62*) (v1.94.1) with ancestry-stratified genomic relationship matrices built from unrelated individuals. All models adjusted for age, age^2^, sex, and 16 genetic PCs. Significance of each *h^2^* estimate was assessed via GCTA’s likelihood-ratio test and corrected for multiple testing (540 tests) using Benjamini-Hochberg FDR. Estimates falling outside the valid (0,1) range were excluded from summary statistics.

