## Supplementary Figures for "Proteomic Signatures Capture Environmental Exposures Across Diverse Populations"

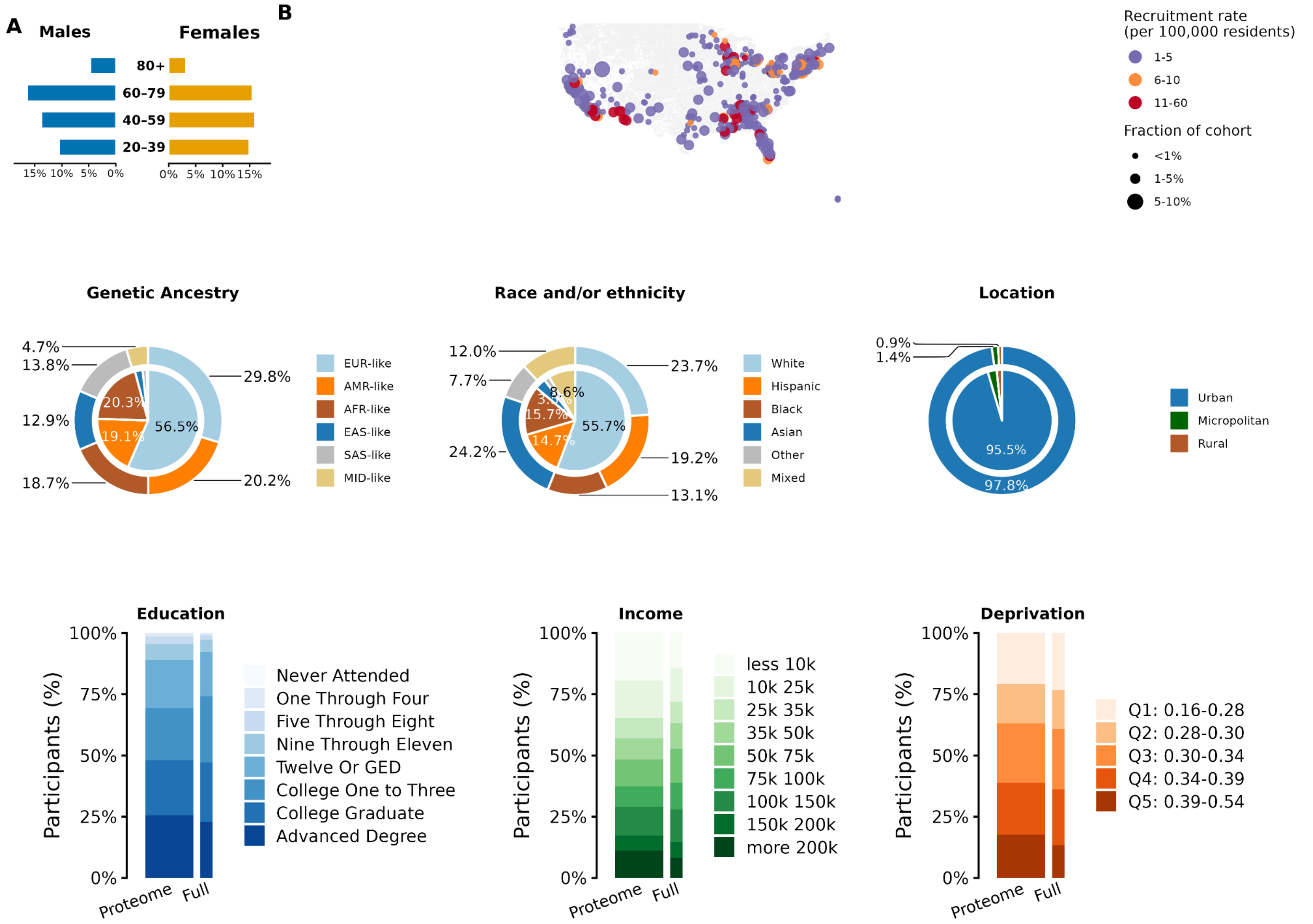


**Fig. S1. Geographic recruitment and demographic composition of the All of Us cohort: comparison between the Proteomics subset and the full cohort.** (A) Sex and age distributions for males and females. (B) The main map shows the AoU proteomics participants recruitment rates per 100,000 residents across the United States, while the inset map shows the same metric for the full AoU cohort. (C) Pie charts compare distributions of genetic ancestry, race and/or ethnicity, and location, where the outer ring denotes the proteomics AoU subset and the inner filled pie denotes the full AoU cohort. (D) Stacked bars summarize income, education, and deprivation among proteomics participants. The first (thicker) bar is the proteomics AoU subset and the second (thinner) bar is the full AoU cohort.

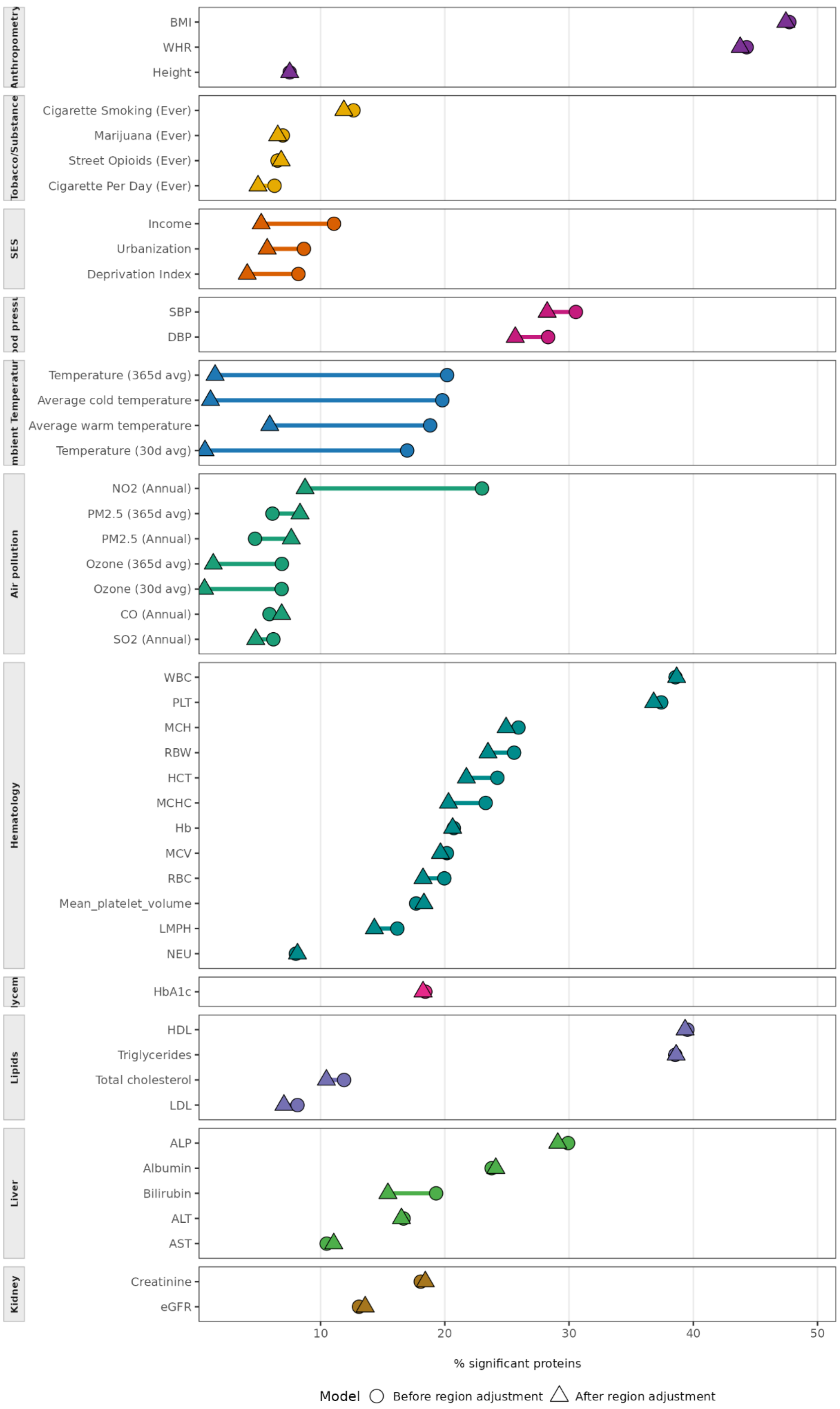


**Fig. S2. Proportion of significant proteins across geocoded exposures before and after adjusting for geographical region (NorthEast/MidWest/Western/Southern).**

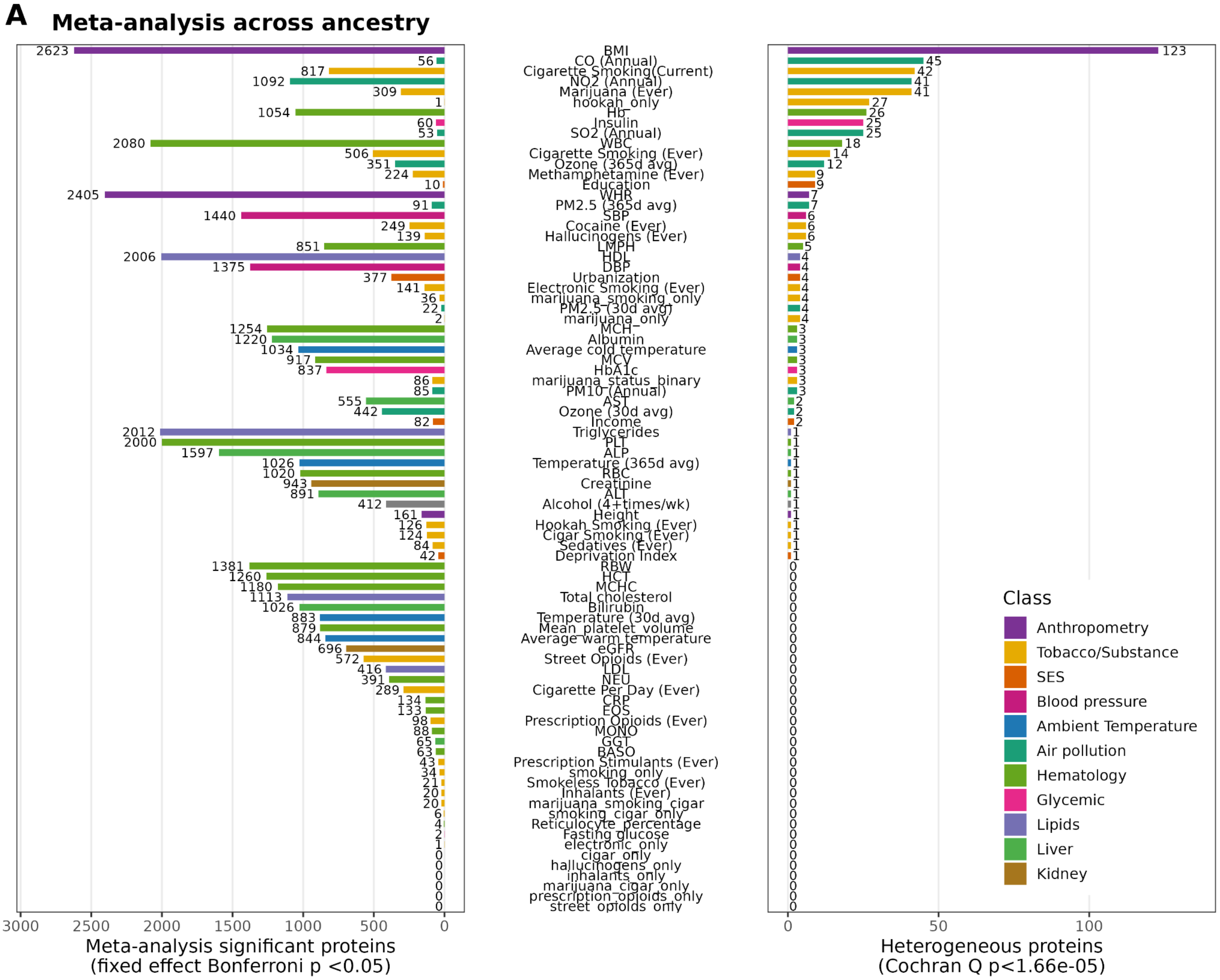
A
Meta-analysis across ancestry
BMI
CO (Annual)
Cigarette Smoking(Current)
NO2 (Annual)
Marijuana (Ever)
hookah_only
Hb
Insulin
SO2 (Annual)
WBC
Cigarette Smoking (Ever)
Ozone (365d avg)
Methamphetamine (Ever)
Education
WHR
PM2.5 (365d avg)
SBP
Cocaine (Ever)
Hallucinogens (Ever)
LMPH
HDL
DBP
Urbanization
Electronic Smoking (Ever)
marijuana_smoking_only
PM2.5 (30d avg)
marijuana_only
MCH
Albumin
Average cold temperature
MCV
HbA1c
marijuana_status_binary
PM10 (Annual)
AST
Ozone (30d avg)
Income
Triglycerides
PLT
ALP
Temperature (365d avg)
RBC
Creatinine
ALT
Alcohol (4+times/wk)
Height
Hookah Smoking (Ever)
Cigar Smoking (Ever)
Sedatives (Ever)
Deprivation Index
RBW
HCT
MCHC
Total cholesterol
Bilirubin
Temperature (30d avg)
Mean_platelet_volume
Average warm temperature
eGFR
Street Opioids (Ever)
LDL
NEU
Cigarette Per Day (Ever)
CRP
EOS
Prescription Opioids (Ever)
MONO
GGT
BASO
Prescription Stimulants (Ever)
smoking_only
Smokeless Tobacco (Ever)
Inhalants (Ever)
marijuana_smoking_cigar
smoking_cigar_only
Reticulocyte_percentage
Fasting glucose
electronic_only
cigar_only
hallucinogens_only
inhalants_only
marijuana_cigar_only
prescription_opioids_only
street_opioids_only
Meta-analysis significant proteins
(fixed effect Bonferroni p <0.05)
Heterogeneous proteins
(Cochran Q p<1.66e-05)
Class
Anthropometry
Tobacco/Substance
SES
Blood pressure
Ambient Temperature
Air pollution
Hematology
Glycemic
Lipids
Liver
Kidney

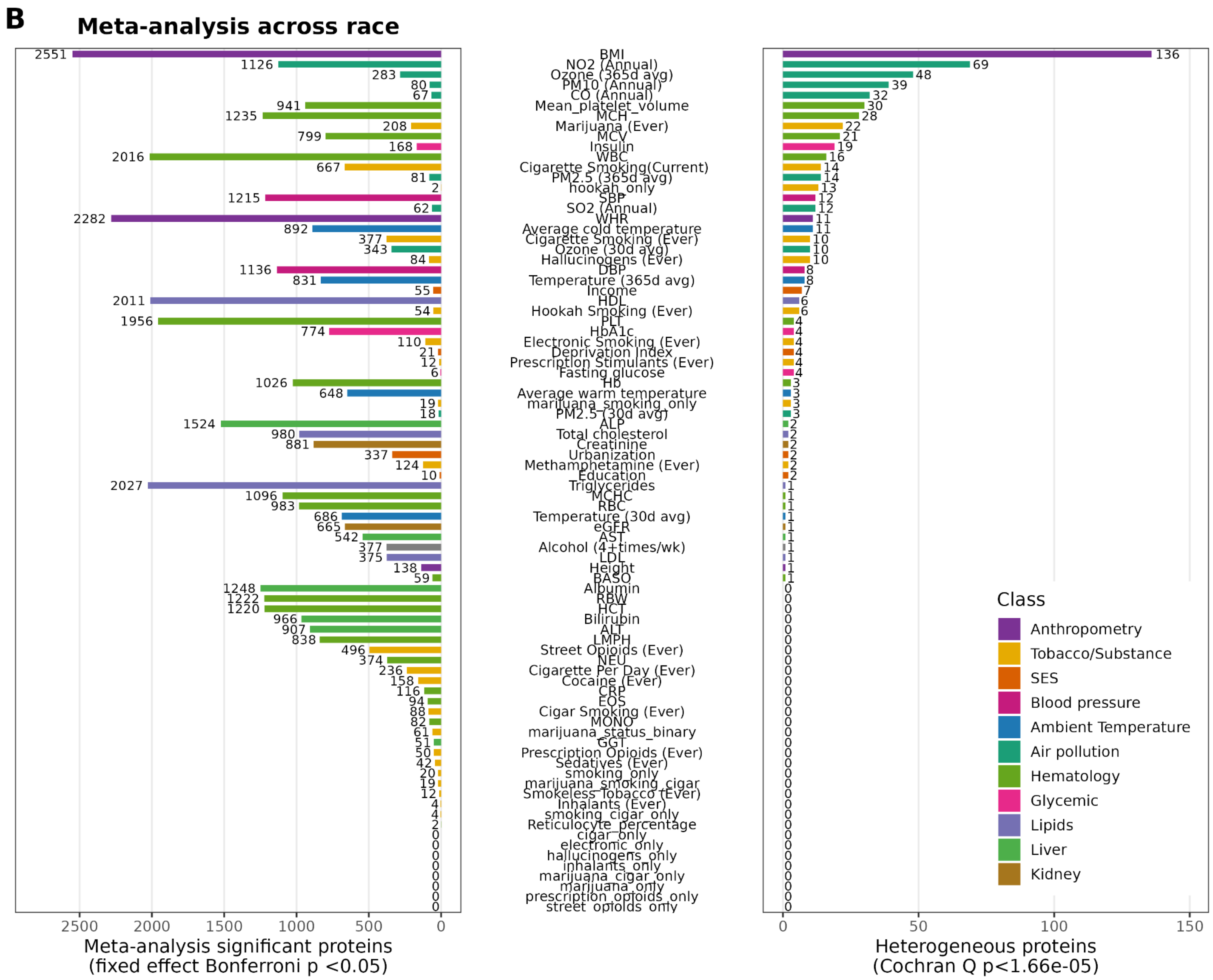


**Fig. S3. Number of heterogeneous proteins.** (A) across genetic ancestry and (B) self-identified race and/or ethnicity groups. Significant meta-analysis across ancestry groups (fixed-effect Bonferroni-adjusted P < 0.05) as well as heterogeneity between groups (Cochran's Q P < 0.05/3011 [number of independent proteins]).

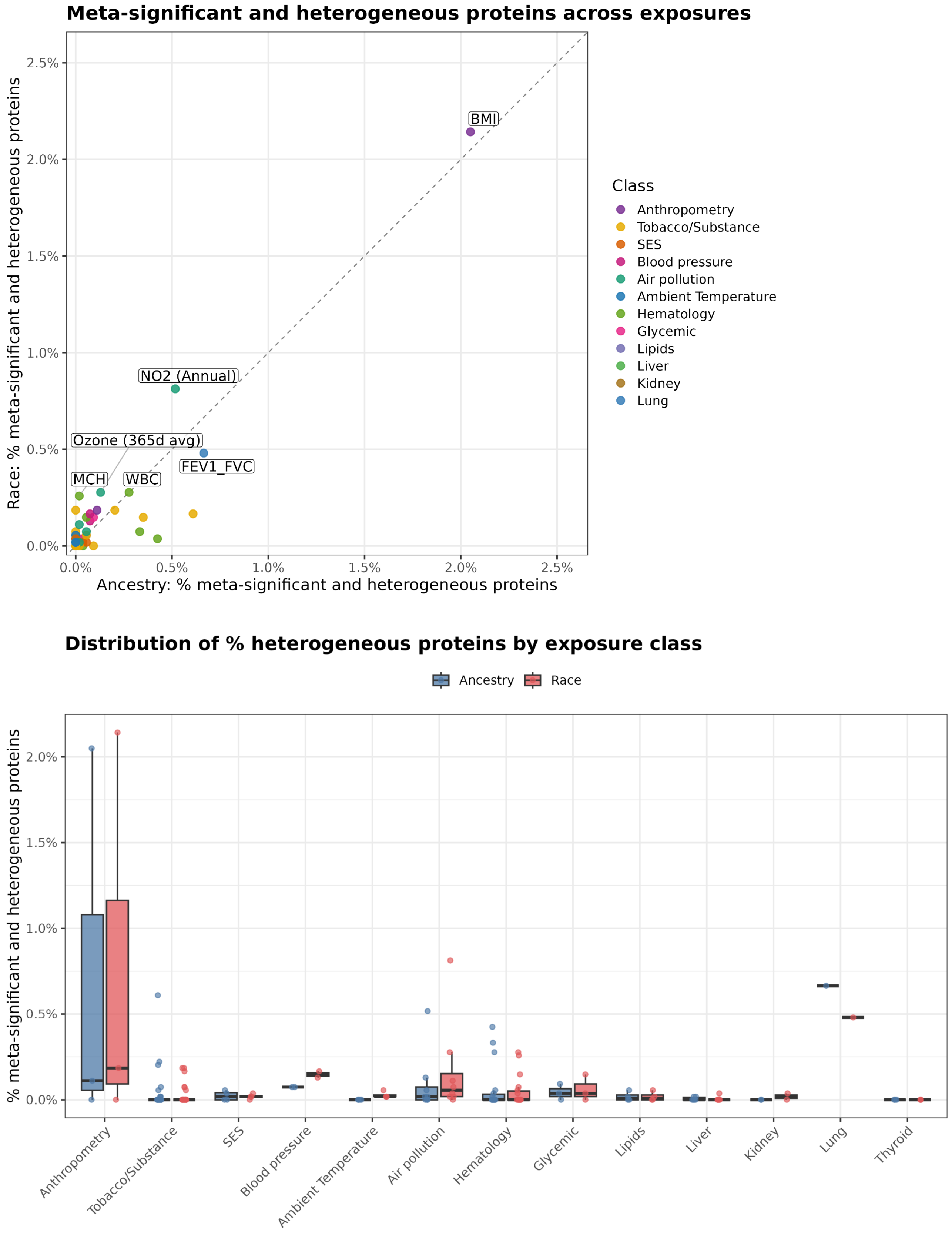


**Fig. S4. Percentage of proteins for each exposure with heterogeneous effects across ancestry and race and/or ethnicity group.**

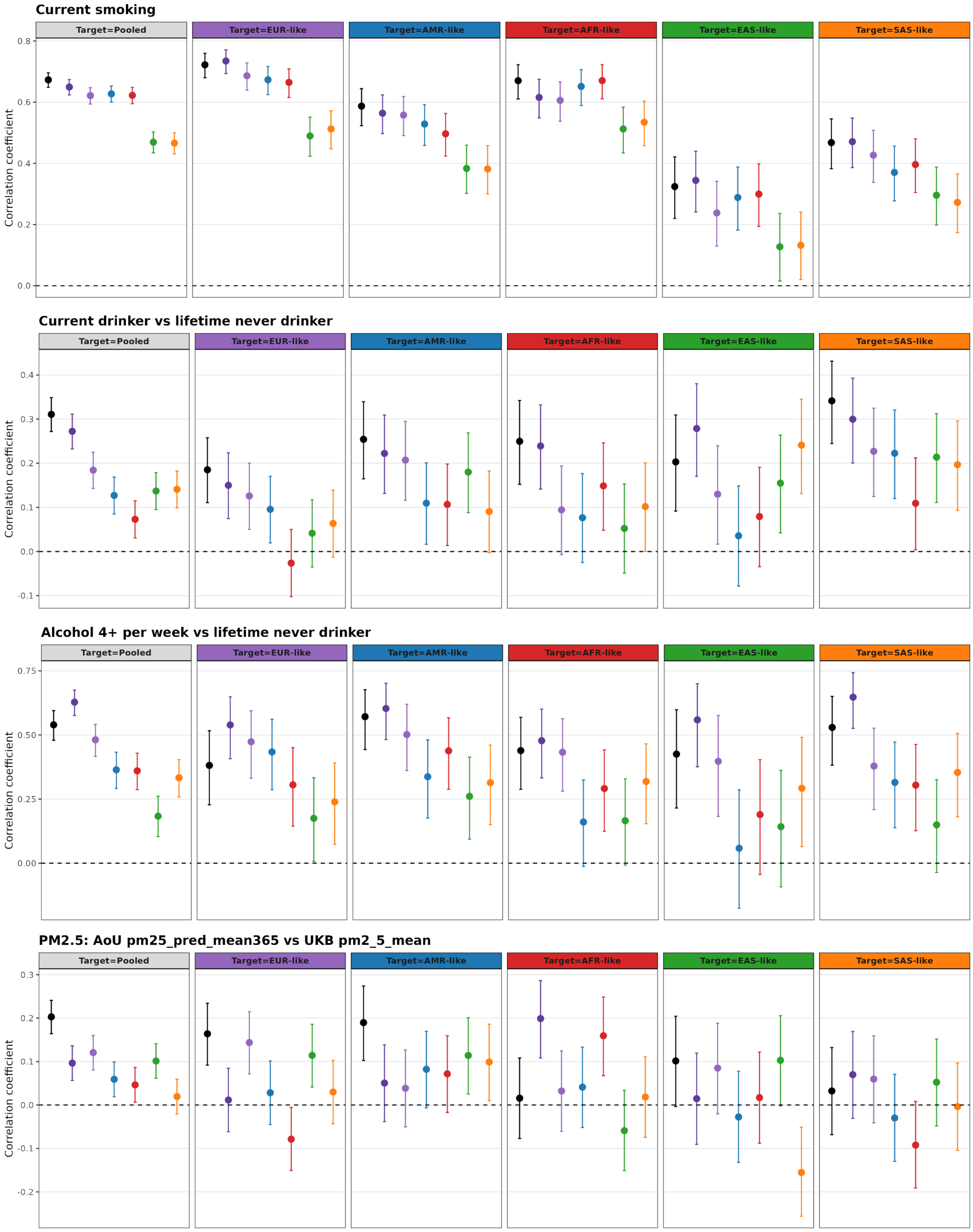
Current smoking
Target=Pooled
Target=EUR-like
Target=AMR-like
Target=AFR-like
Target=EAS-like
Target=SAS-like
Correlation coefficient
0.8
0.6
0.4
0.2
0.0
Current drinker vs lifetime never drinker
Target=Pooled
Target=EUR-like
Target=AMR-like
Target=AFR-like
Target=EAS-like
Target=SAS-like
Correlation coefficient
0.4
0.3
0.2
0.1
0.0
-0.1
Alcohol 4+ per week vs lifetime never drinker
Target=Pooled
Target=EUR-like
Target=AMR-like
Target=AFR-like
Target=EAS-like
Target=SAS-like
Correlation coefficient
0.75
0.50
0.25
0.00
PM2.5: AoU pm25_pred_mean365 vs UKB pm2_5_mean
Target=Pooled
Target=EUR-like
Target=AMR-like
Target=AFR-like
Target=EAS-like
Target=SAS-like
Correlation coefficient
0.3
0.2
0.1
0.0
-0.1
-0.2

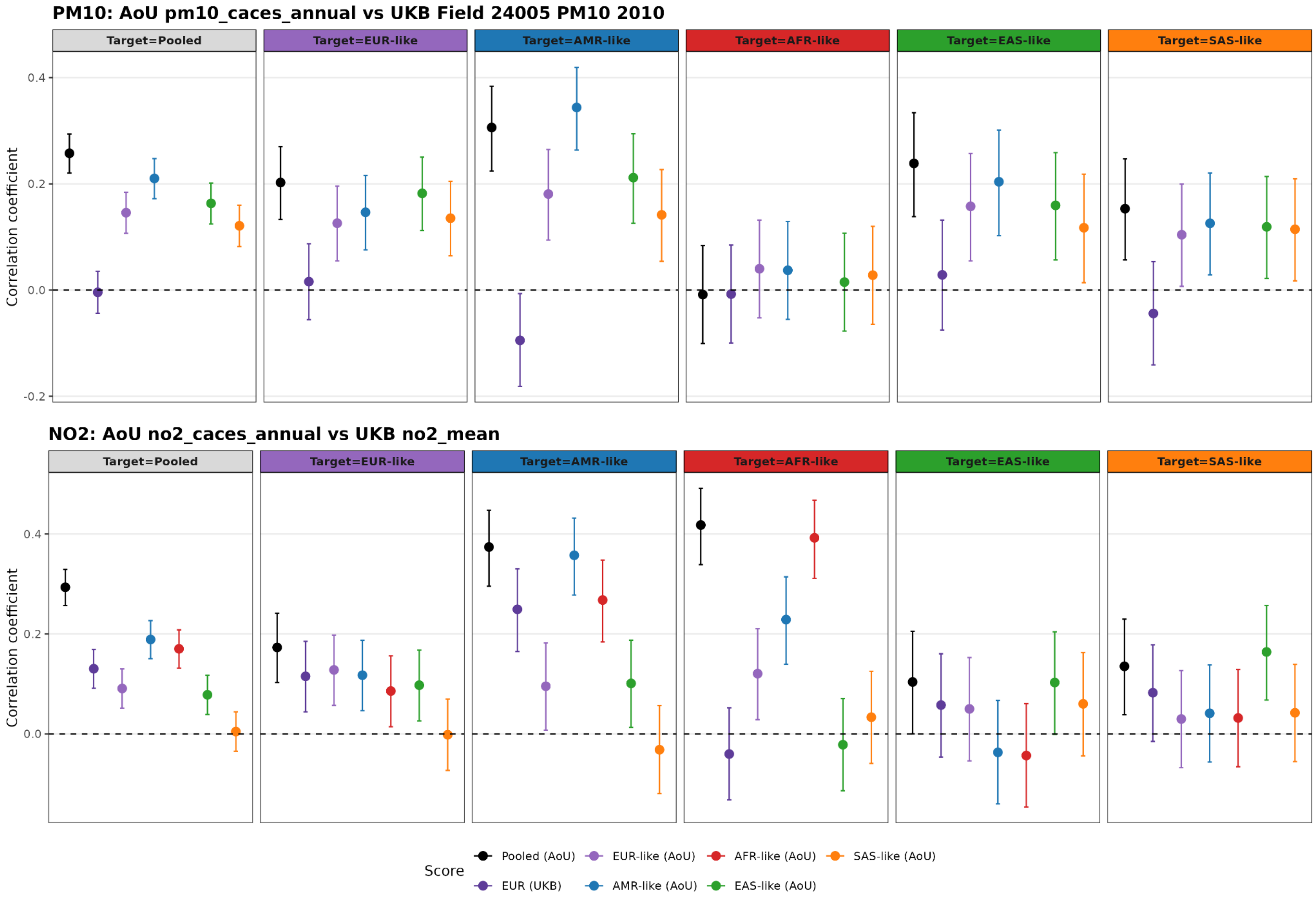


**Fig. S5. Validation of UKB-trained, pooled- and ancestry-stratified AoU-trained protein scores for smoking alcohol, PM2.5, PM10, and NO2 in AoU test set ancestry groups.**

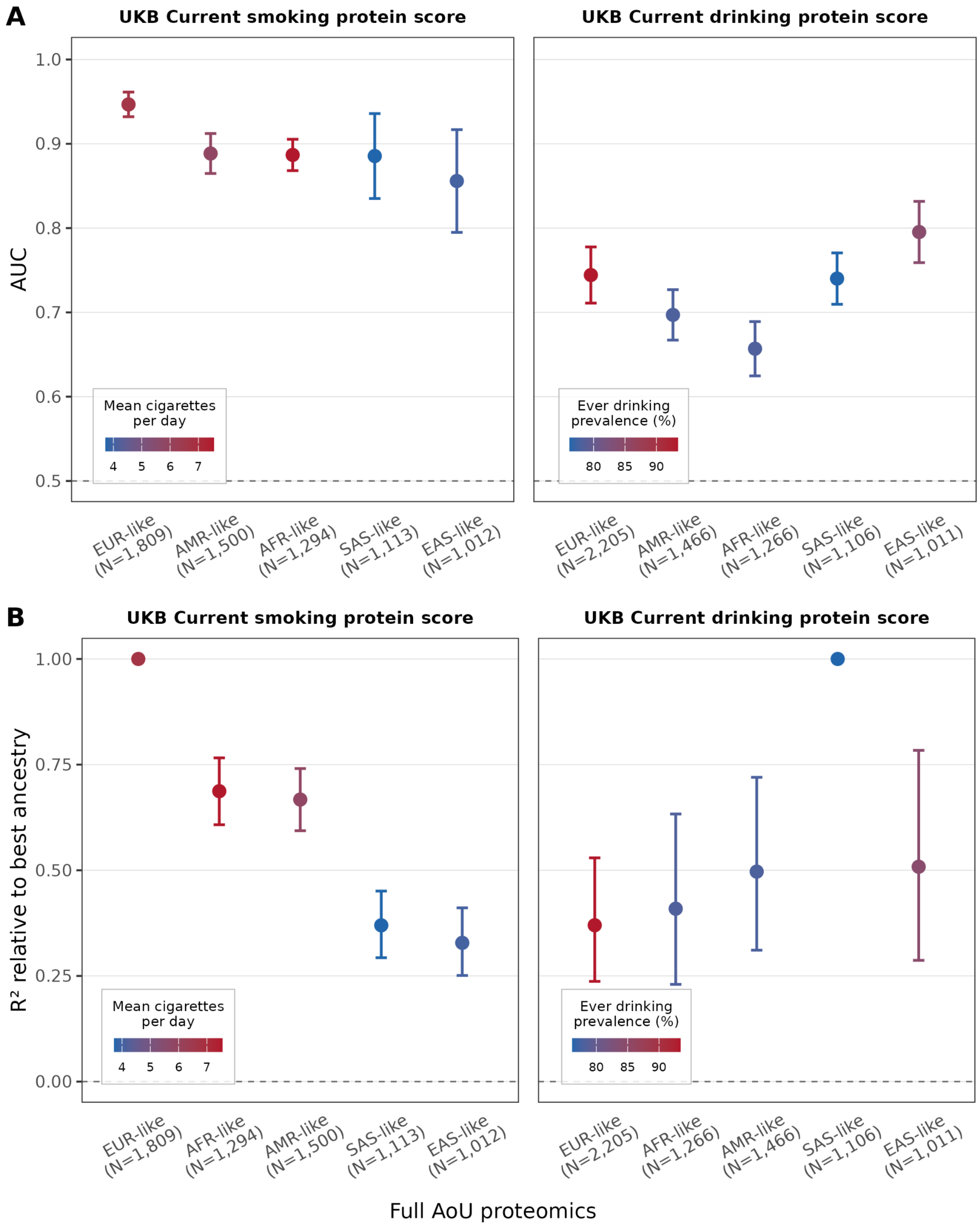


**Fig. S6. Cross-cohort validation of UK Biobank (UKB)-trained protein-exposure scores for smoking and alcohol drinking in the All of Us (AoU) full proteomics cohort.** UKB-trained scores for current smoking and current alcohol drinking were applied to AoU participants and evaluated within genetically inferred ancestry groups. Performance is shown as the area under the curve (AUC) (A) and the point-biserial correlation coefficient ($R^2$) between the protein score and exposure status (B). Point colors indicate ancestry-specific mean cigarette per day (smoking panel) and ever-drinking prevalence (alcohol panel).

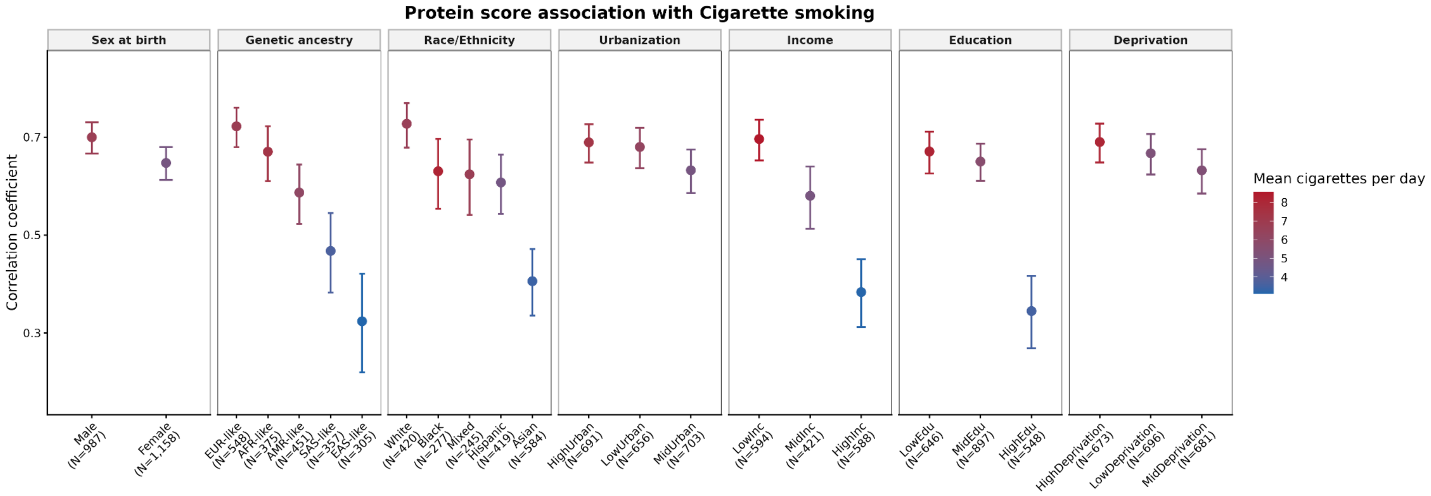
Protein score association with Cigarette smoking
Sex at birth
Genetic ancestry
Race/Ethnicity
Urbanization
Income
Education
Deprivation
Correlation coefficient
0.7
0.5
0.3
Mean cigarettes per day
8
7
6
5
4
Male (N=987)
Female (N=1,158)
EUR-like (N=548)
AFR-like (N=375)
AMR-like (N=451)
SAS-like (N=357)
EAS-like (N=305)
White (N=420)
Black (N=277)
Mixed (N=245)
Hispanic (N=419)
Asian (N=584)
HighUrban (N=691)
LowUrban (N=656)
MidUrban (N=703)
LowInc (N=594)
MidInc (N=421)
HighInc (N=588)
LowEdu (N=646)
MidEdu (N=897)
HighEdu (N=548)
HighDeprivation (N=673)
LowDeprivation (N=696)
MidDeprivation (N=681)

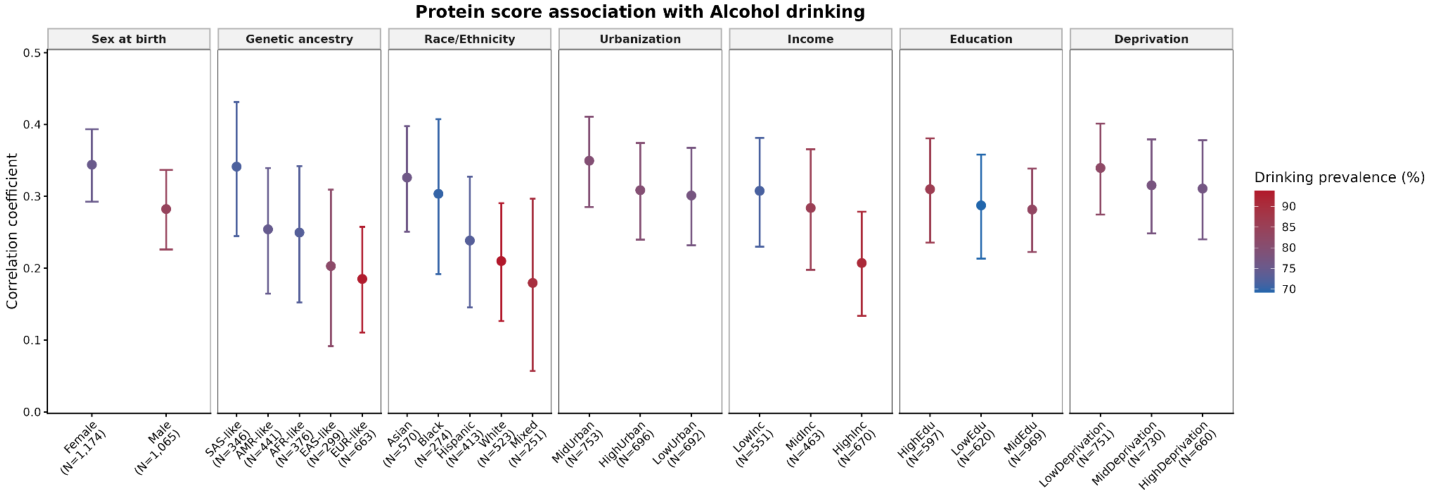
Protein score association with Alcohol drinking
Sex at birth
Genetic ancestry
Race/Ethnicity
Urbanization
Income
Education
Deprivation
Correlation coefficient
0.5
0.4
0.3
0.2
0.1
0.0
Drinking prevalence (%)
90
85
80
75
70
Female (N=1,174)
Male (N=1,065)
SAS-like (N=346)
AMR-like (N=441)
AFR-like (N=376)
EAS-like (N=299)
EUR-like (N=663)
Asian (N=570)
Black (N=274)
Hispanic (N=413)
White (N=523)
Mixed (N=251)
MidUrban (N=753)
HighUrban (N=696)
LowUrban (N=692)
LowInc (N=551)
MidInc (N=463)
HighInc (N=670)
HighEdu (N=597)
LowEdu (N=620)
MidEdu (N=969)
LowDeprivation (N=751)
MidDeprivation (N=730)
HighDeprivation (N=660)

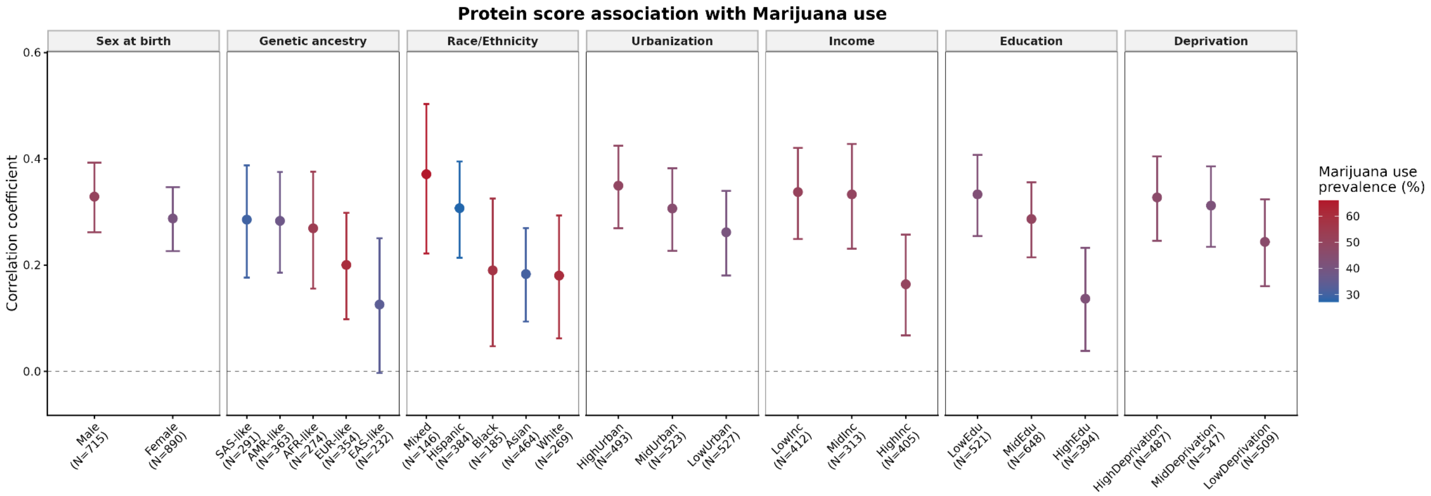
Protein score association with Marijuana use
Sex at birth
Genetic ancestry
Race/Ethnicity
Urbanization
Income
Education
Deprivation
Correlation coefficient
0.6
0.4
0.2
0.0
Marijuana use prevalence (%)
60
50
40
30
Male (N=715)
Female (N=890)
SAS-like (N=291)
AMR-like (N=363)
AFR-like (N=274)
EUR-like (N=354)
EAS-like (N=232)
Mixed (N=146)
Hispanic (N=384)
Black (N=185)
Asian (N=464)
White (N=269)
HighUrban (N=493)
MidUrban (N=523)
LowUrban (N=527)
LowInc (N=412)
MidInc (N=313)
HighInc (N=405)
LowEdu (N=521)
MidEdu (N=648)
HighEdu (N=394)
HighDeprivation (N=487)
MidDeprivation (N=547)
LowDeprivation (N=509)

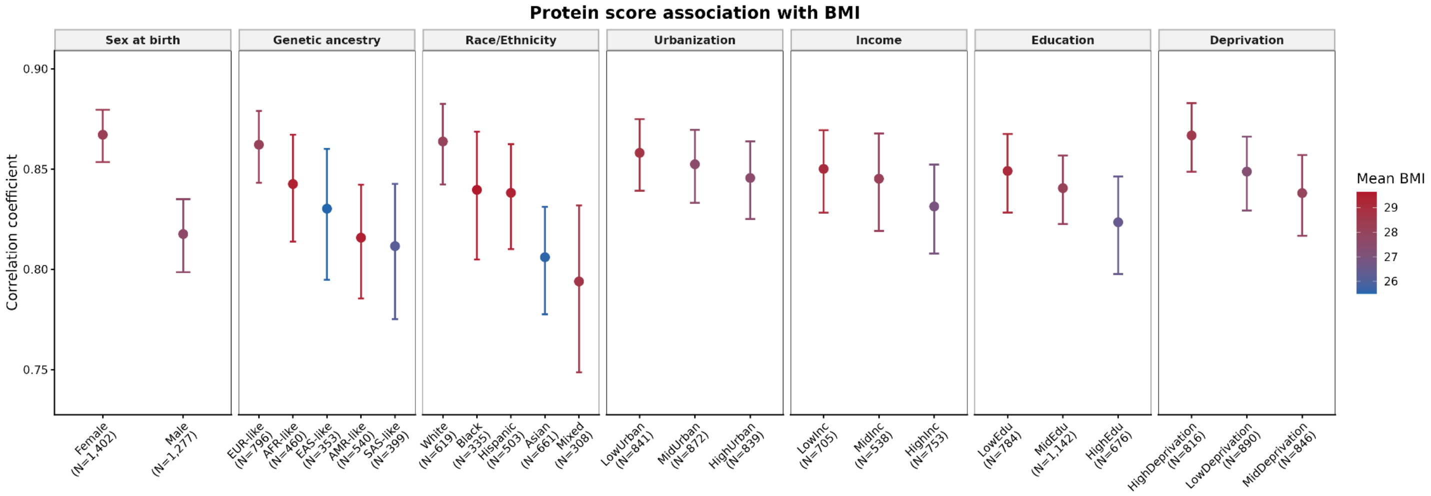
Protein score association with BMI
Sex at birth
Genetic ancestry
Race/Ethnicity
Urbanization
Income
Education
Deprivation
Correlation coefficient
Mean BMI
Female (N=1,402)
Male (N=1,277)
EUR-like (N=796)
AFR-like (N=460)
EAS-like (N=353)
AMR-like (N=540)
SAS-like (N=399)
White (N=619)
Black (N=335)
Hispanic (N=503)
Asian (N=661)
Mixed (N=308)
LowUrban (N=841)
MidUrban (N=872)
HighUrban (N=839)
LowInc (N=705)
MidInc (N=538)
HighInc (N=753)
LowEdu (N=784)
MidEdu (N=1,142)
HighEdu (N=676)
HighDeprivation (N=816)
LowDeprivation (N=890)
MidDeprivation (N=846)

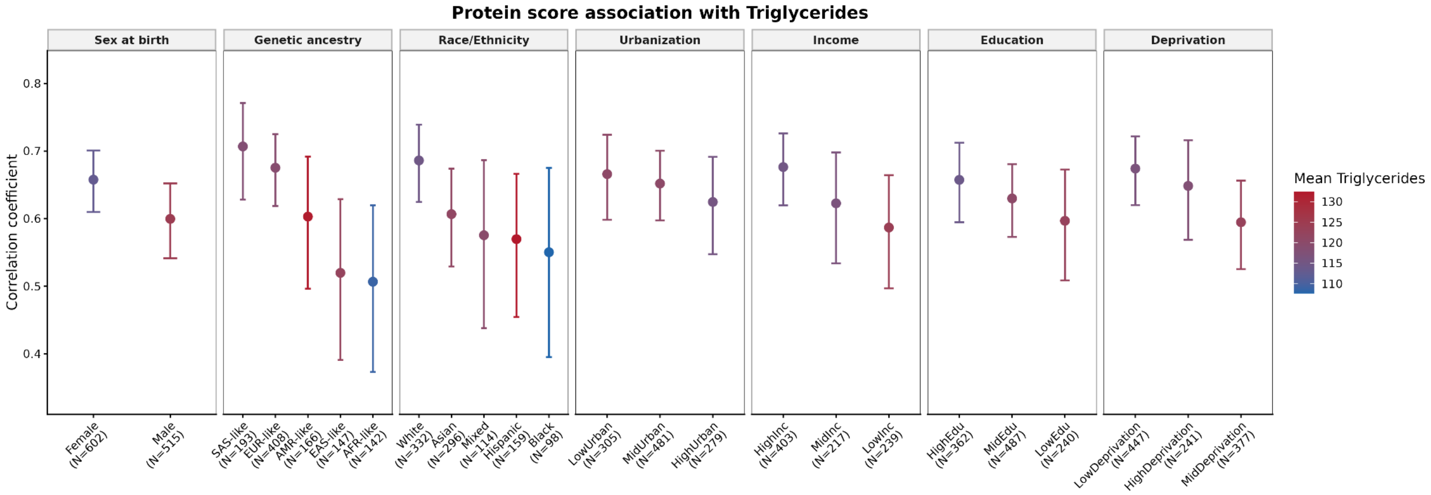
Protein score association with Triglycerides
Sex at birth
Genetic ancestry
Race/Ethnicity
Urbanization
Income
Education
Deprivation
Correlation coefficient
Mean Triglycerides
Female (N=602)
Male (N=515)
SAS-like (N=193)
EUR-like (N=408)
AMR-like (N=166)
EAS-like (N=147)
AFR-like (N=142)
White (N=332)
Asian (N=296)
Mixed (N=114)
Hispanic (N=159)
Black (N=98)
LowUrban (N=305)
MidUrban (N=481)
HighUrban (N=279)
HighInc (N=403)
MidInc (N=217)
LowInc (N=239)
HighEdu (N=362)
MidEdu (N=487)
LowEdu (N=240)
LowDeprivation (N=447)
HighDeprivation (N=241)
MidDeprivation (N=377)

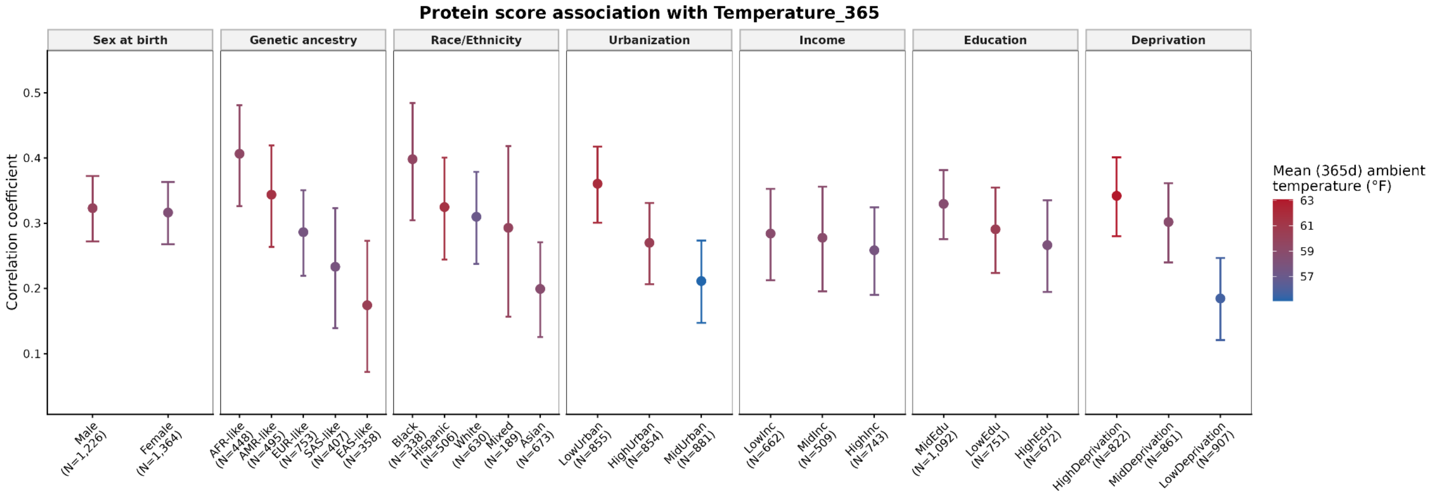
Protein score association with Temperature_365
Sex at birth
Genetic ancestry
Race/Ethnicity
Urbanization
Income
Education
Deprivation
Correlation coefficient
Mean (365d) ambient temperature (°F)
Male (N=1,226)
Female (N=1,364)
AFR-like (N=448)
AMR-like (N=495)
EUR-like (N=753)
SAS-like (N=407)
EAS-like (N=358)
Black (N=338)
Hispanic (N=506)
White (N=630)
Mixed (N=189)
Asian (N=673)
LowUrban (N=855)
HighUrban (N=854)
MidUrban (N=881)
LowInc (N=662)
MidInc (N=509)
HighInc (N=743)
MidEdu (N=1,092)
LowEdu (N=751)
HighEdu (N=672)
HighDeprivation (N=822)
MidDeprivation (N=861)
LowDeprivation (N=907)

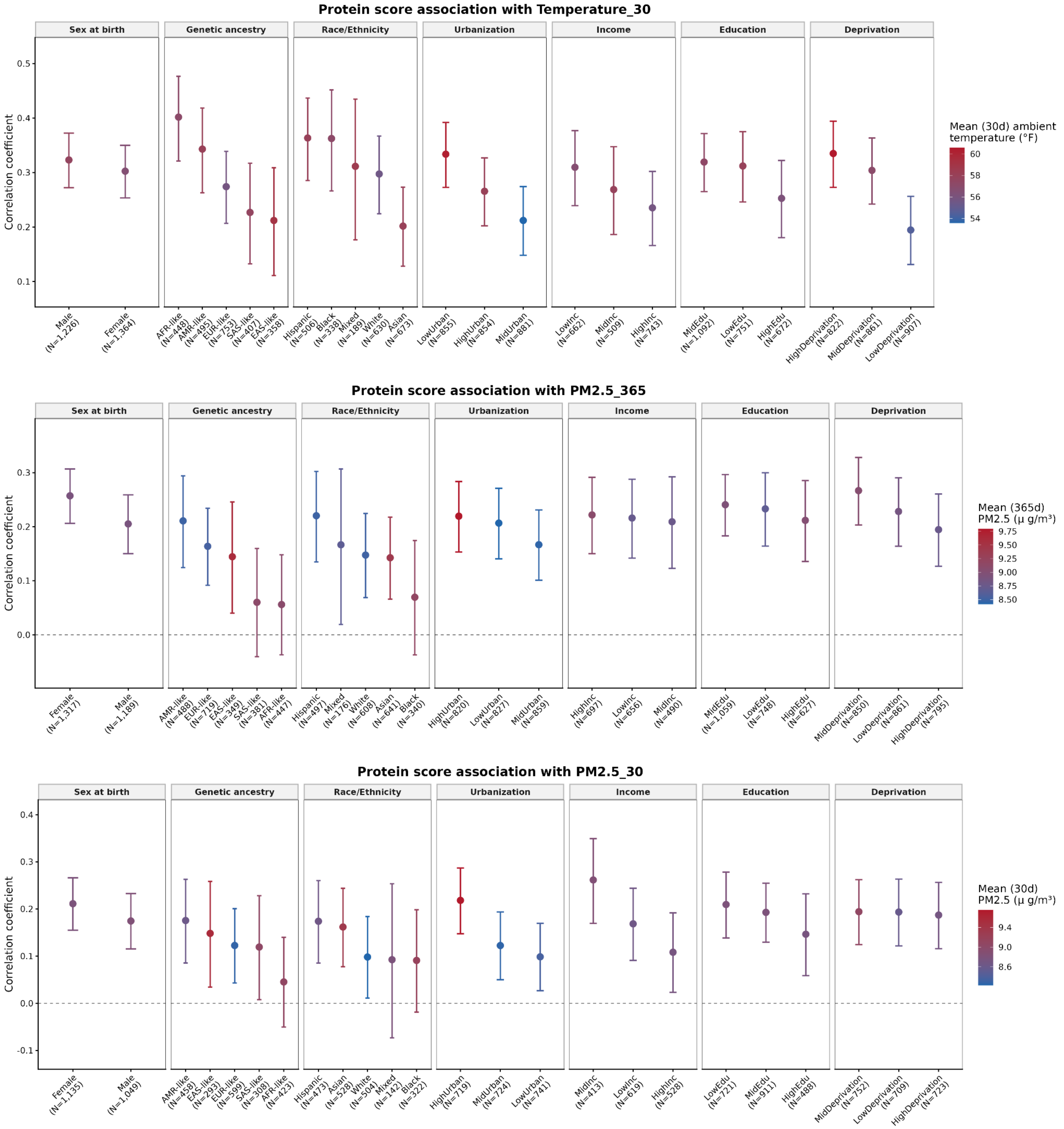


**Fig. S7. Validation of AoU protein scores for smoking, alcohol drinking, marijuana use, body mass index, triglycerides, ambient temperature, and PM2.5 across AoU test set strata defined by sex, ancestry, race and/or ethnicity, urbanization, income education, and deprivation.**

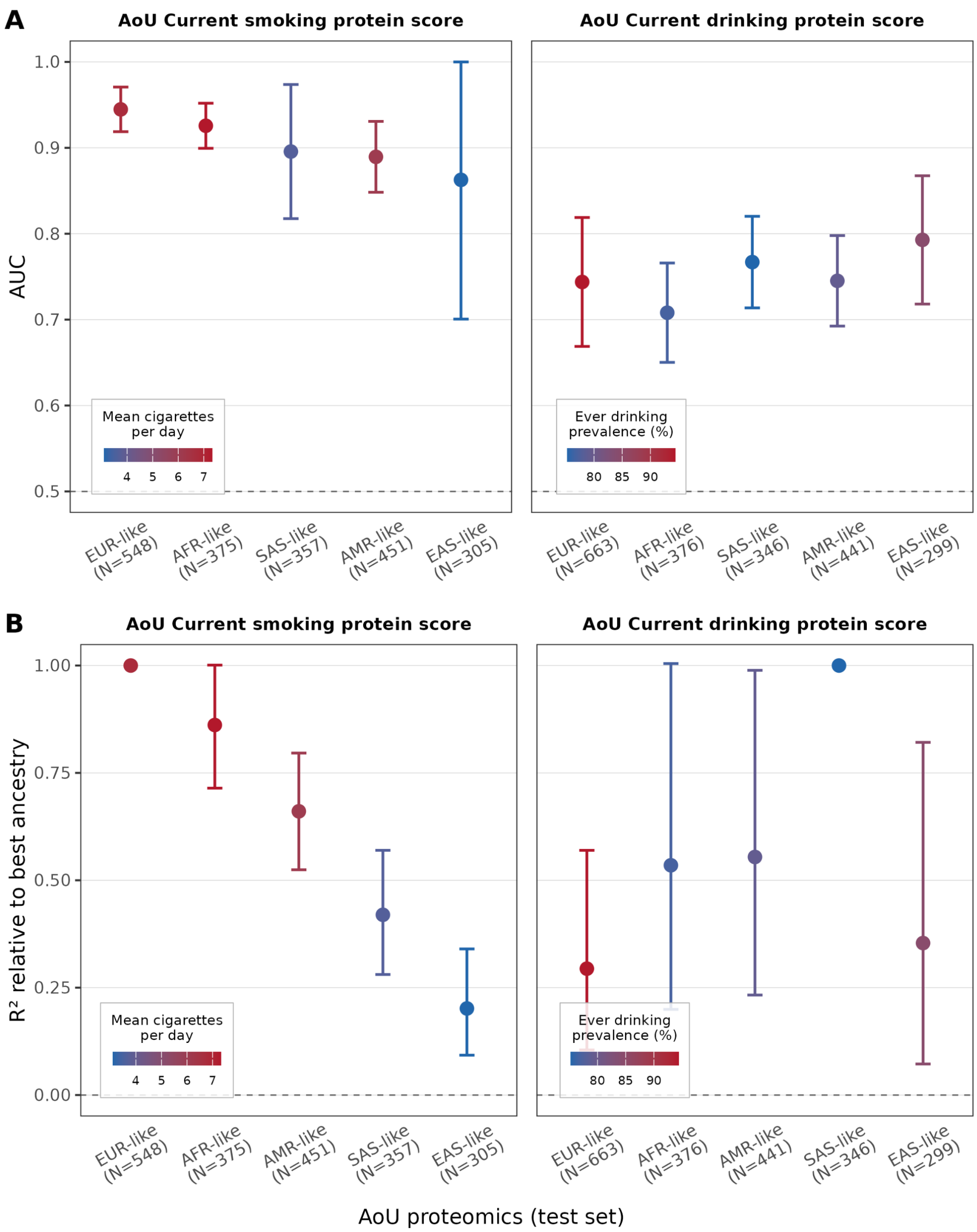


**Fig. S8. Within-cohort validation of AoU-trained protein-exposure scores for smoking and alcohol drinking. Scores were trained in a pooled AoU training set and evaluated in a held-out test set.** Performance is shown as the area under the curve (AUC) (A) and the point-biserial correlation coefficient ($R^2$) between the protein score and exposure status (B). Point colors indicate ancestry-specific mean cigarette per day (smoking panel) and ever-drinking prevalence (alcohol panel).

Score-exposure correlation (PS_targets_correlation_heatmap.png)

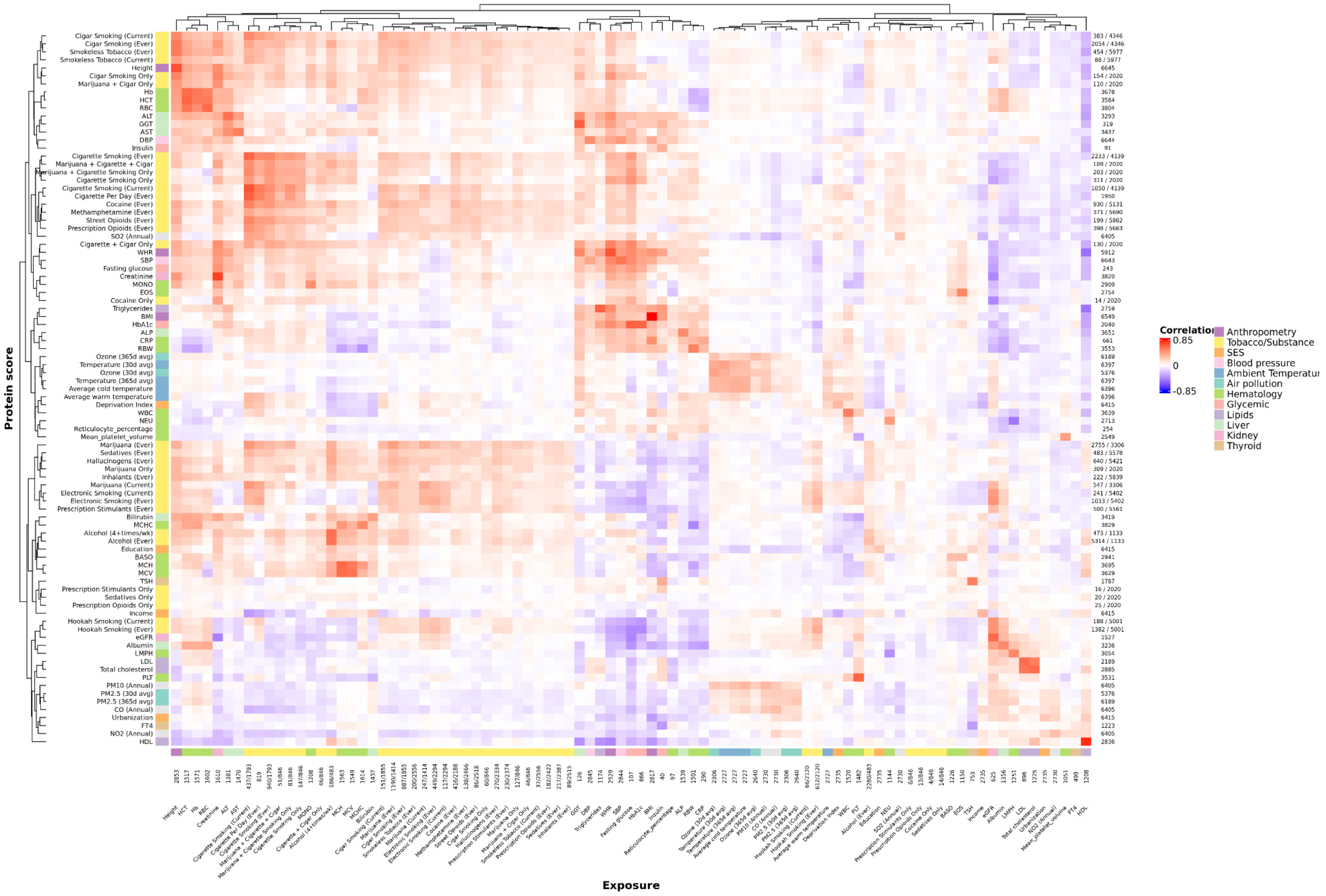


**Fig. S9. Correlation matrix between protein scores and exposures (table S9).**

Exposure-exposure correlation (phenotypic_exposure_correlation_heatmap_clustered.png)

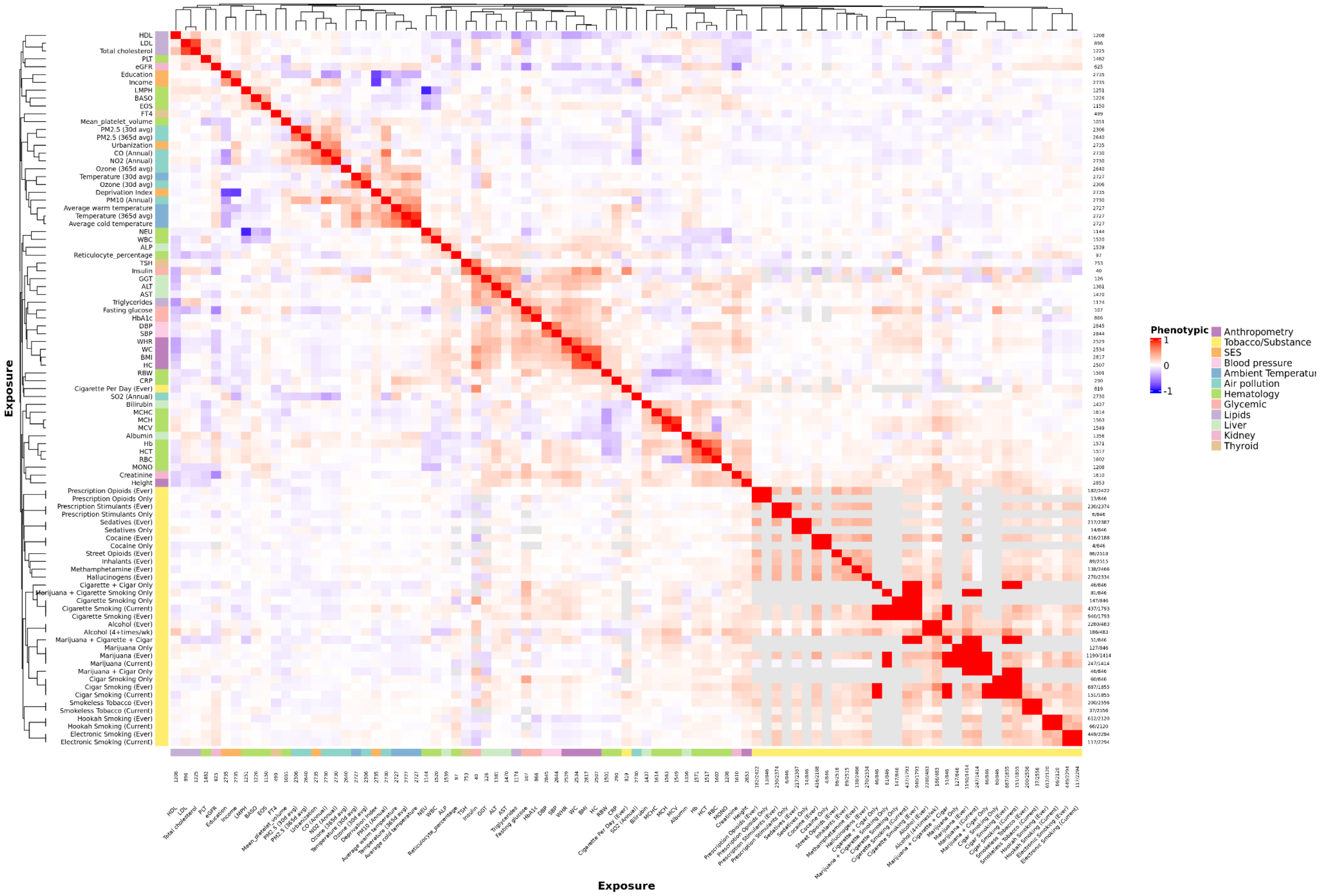


**Fig. S10. Correlation matrix between exposures and exposures (table S10).**

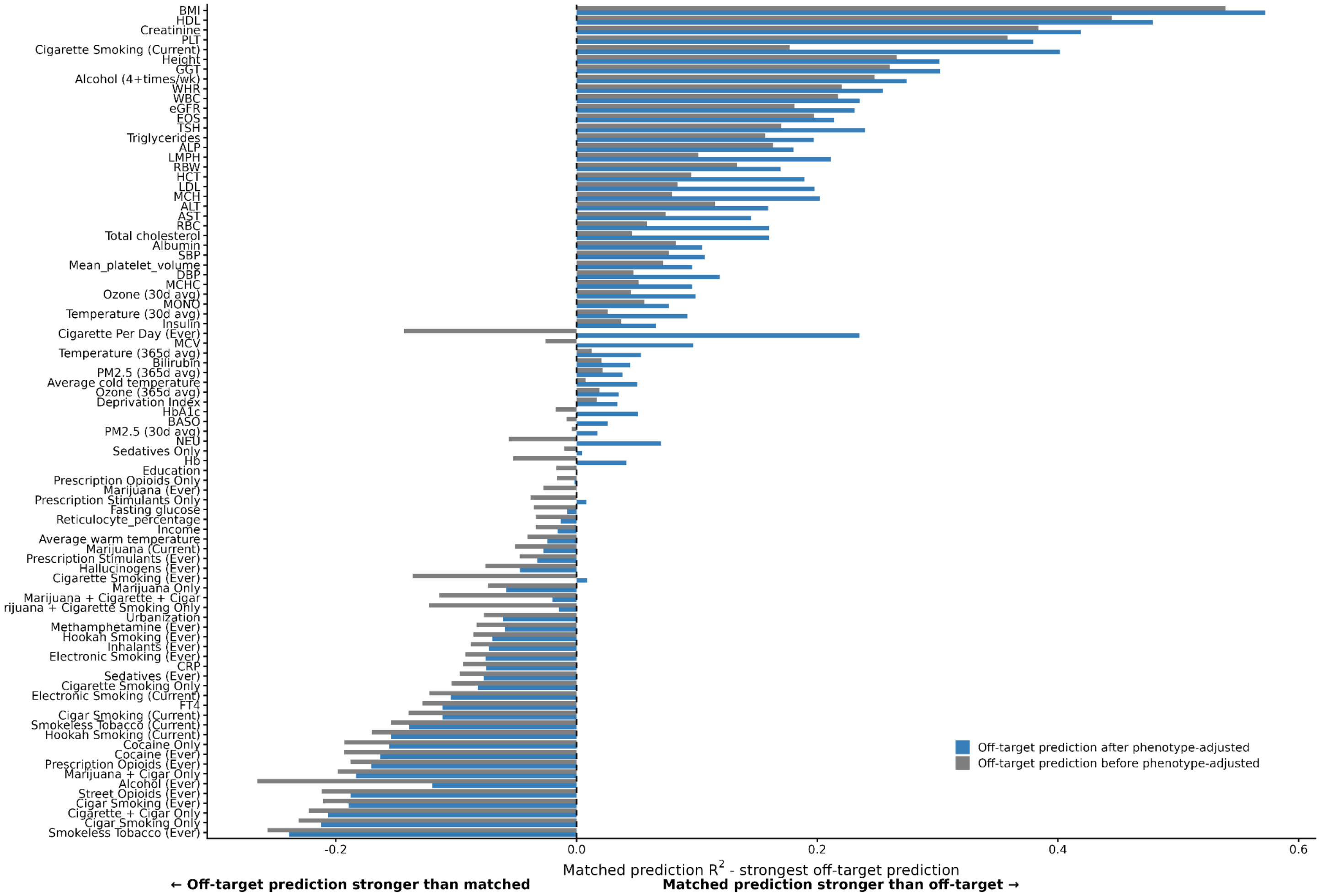


**Fig. S11. Exposure specificity before and after adjustment for phenotypic correlation.** Positive values indicate that the matched exposure was predicted more strongly than the strongest off-target exposure. Negative values indicate stronger off-target than matched prediction. The phenotype-adjusted value uses off-target prediction residuals after regressing off-target score-exposure $R^2$ on phenotypic exposure-exposure $R^2$. We first fit a regression across all off-diagonal pairs, $R^2(PS_i, E_i) = \beta_0 + \beta_1 R^2(E_i, E_j) + \epsilon_{ij}$. For each off-diagonal pair, residual cross-prediction was defined as the observed score-exposure $R^2$ minus the score-exposure $R^2$ predicted from the phenotypic exposure-exposure $R^2$. We then compared each score's matched $R^2$ with its largest residual off-target prediction.

Off-target prediction before phenotype-adjustment = $R^2(PS_i, E_i) - \max_{j \neq i} R^2(PS_i, E_j)$ .

Off-target prediction after phenotype-adjustment = $R^2(PS_i, E_i) - \max_{j \neq i}\{R^2(PS_i, E_j) - \hat{E}\,[\,R^2(PS_i, E_j) | R^2(E_i, E_j)]\}$ .

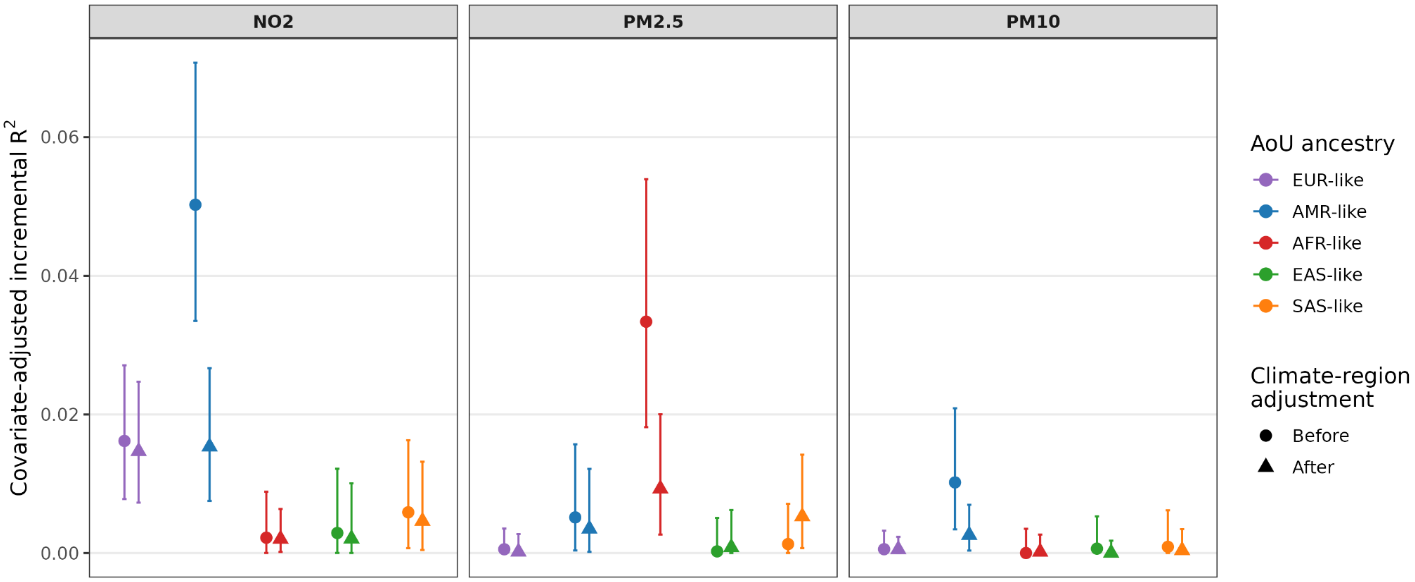


**Fig. S12. UKB air-pollution protein score performance across AoU ancestry groups adjusted for age, sex, and blood collection time and additional adjustment for region.**

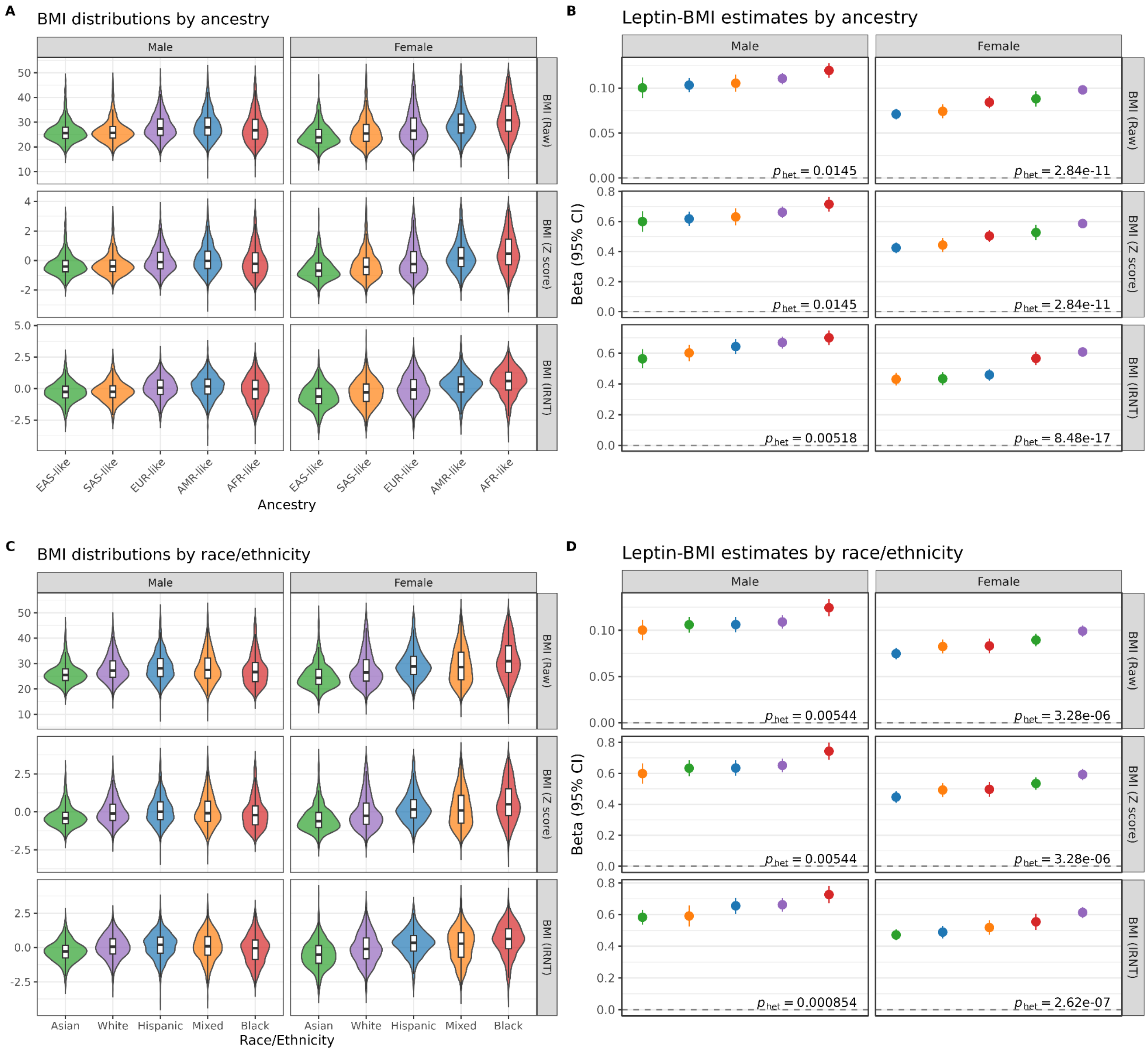


**Fig. S13. Cross-ancestry and cross-race and/or ethnicity heterogeneity in BMI associations with leptin in male and female.** (A). Distributions of body mass index (BMI) across genetic ancestry groups shown for raw values, z score, and inverse rank normal transformed (IRNT) scales. (B). Ancestry-specific associations between BMI and circulating leptin levels. Points represent regression coefficients (β) from linear models of leptin on BMI, adjusted for age, sex, biosample collection year, month, and time; $p_{\text{het}}$ indicate heterogeneity test results. (C). Distributions of body mass index (BMI) across self-reported race and/or ethnicity groups shown for raw values, z score, and inverse rank normal transformed (IRNT) scales. (D). Race and/or ethnicity-specific associations between BMI and circulating leptin levels. Points represent regression coefficients (β) from linear models of leptin on BMI, adjusted for age, sex, biosample collection year, month, and time; $p_{\text{het}}$ indicate heterogeneity test results.

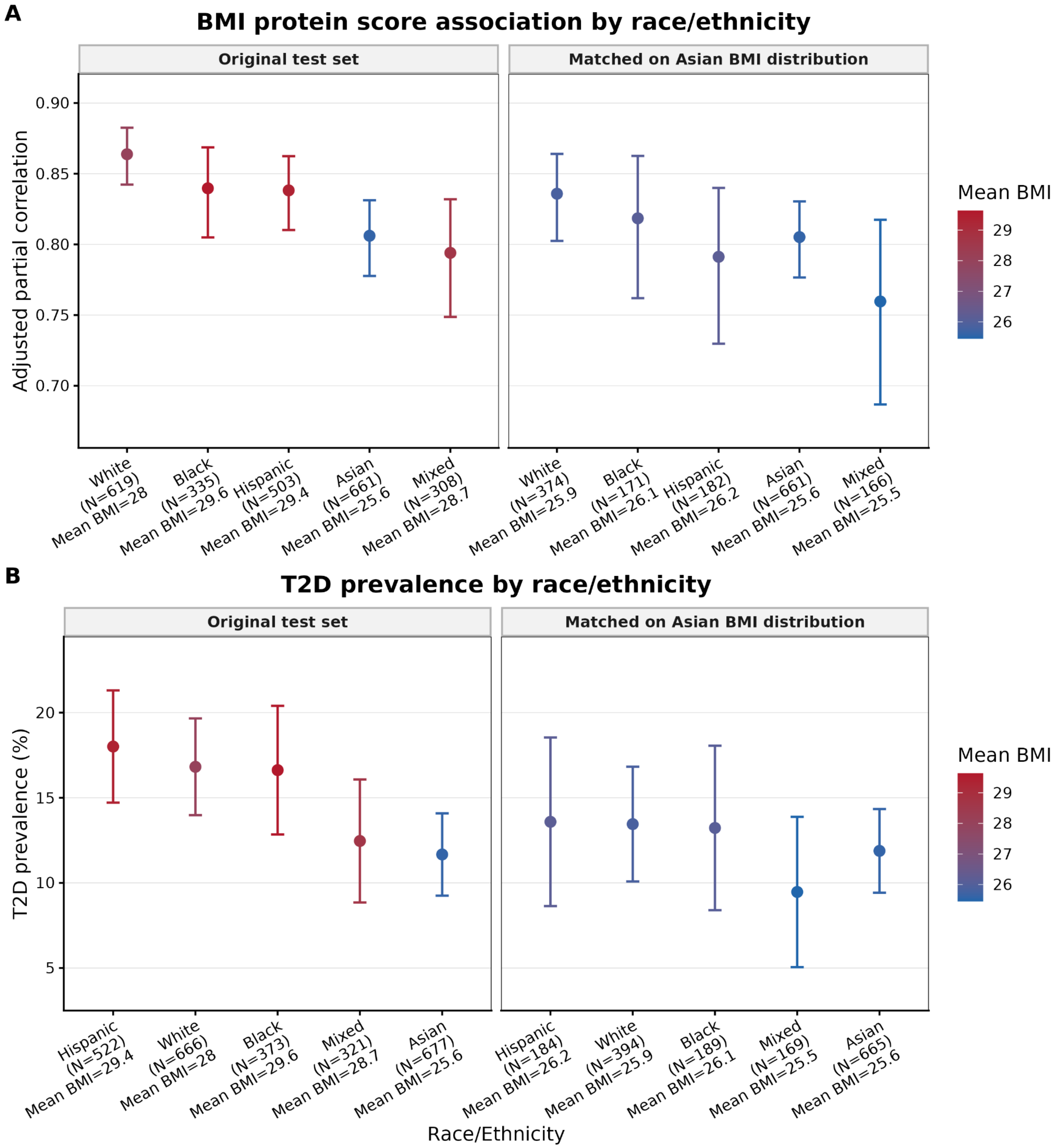
A
BMI protein score association by race/ethnicity
Original test set
Matched on Asian BMI distribution
Adjusted partial correlation
0.90
0.85
0.80
0.75
0.70
White (N=619) Mean BMI=28
Black (N=335) Mean BMI=29.6
Hispanic (N=503) Mean BMI=29.4
Asian (N=661) Mean BMI=25.6
Mixed (N=308) Mean BMI=28.7
White (N=374) Mean BMI=25.9
Black (N=171) Mean BMI=26.1
Hispanic (N=182) Mean BMI=26.2
Asian (N=661) Mean BMI=25.6
Mixed (N=166) Mean BMI=25.5
Mean BMI
29
28
27
26
B
T2D prevalence by race/ethnicity
Original test set
Matched on Asian BMI distribution
T2D prevalence (%)
20
15
10
5
Hispanic (N=522) Mean BMI=29.4
White (N=666) Mean BMI=28
Black (N=373) Mean BMI=29.6
Mixed (N=321) Mean BMI=28.7
Asian (N=677) Mean BMI=25.6
Hispanic (N=184) Mean BMI=26.2
White (N=394) Mean BMI=25.9
Black (N=189) Mean BMI=26.1
Mixed (N=169) Mean BMI=25.5
Asian (N=665) Mean BMI=25.6
Race/Ethnicity
Mean BMI
29
28
27
26

**Fig. S14. BMI distribution-matched analysis across race and/or ethnicity groups.** (A) Performance of the BMI protein score, measured by adjusted partial correlation with observed BMI, across racial/ethnic groups in the original test set (left) and after matching to the Asian BMI distribution (right). (B) Prevalence of type 2 diabetes (T2D, %) across racial/ethnic groups in the original test set (left) and after matching on Asian BMI distribution (right). Points represent point estimates, error bars indicate 95% confidence intervals, and point colors reflect the mean BMI for each group. Asian participants served as the reference distribution and were partitioned into quintile bins extended to $-\infty$ and $+\infty$. Non-Asian participants were stratified into these bins and randomly subsampled to achieve uniform bin sizes constrained by the smallest group count. The complete Asian reference and matched non-Asian groups were re-analyzed for BMI protein score performance, BMI-leptin associations, and T2D prevalence relative to unmatched baseline estimates (table S14).

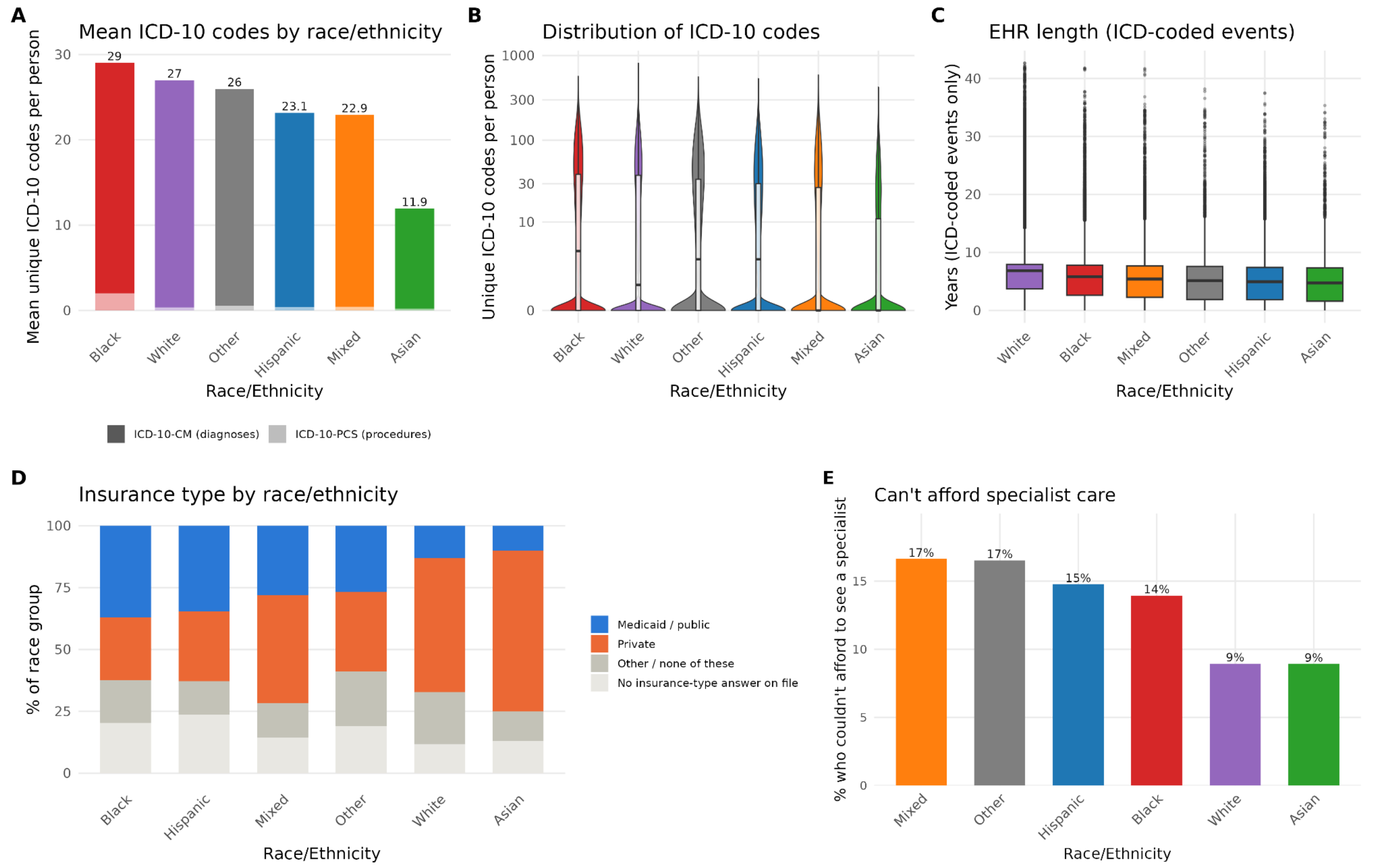


**Fig. S15. Self-reported race and/or ethnicity-stratified patterns in EHR completeness, specialist-care burden, insurance coverage, and EHR history length among All of Us participants (Black=97,795, Hispanic=91,606, Asian=21,826, White=346,965, Mixed=46,886, Other=17,369).** (A) Mean number of unique ICD-10-CM (diagnosis) and ICD-10-PCS (procedure) codes per person, stacked by code type, for each self-identified race and/or ethnicity group. The number above each bar gives the total mean code count. (B) Distribution of the same total unique-code count per person by race/ethnicity group. (C) Mean length of EHR history by race/ethnicity group, restricted to the interval between each person's earliest and latest ICD-10-coded (diagnosis or procedure) encounter, shown as a boxplot. (D) Distribution of self-reported insurance type by race/ethnicity group. Medicaid/public coverage takes priority when a respondent reported both Medicaid and a private plan; "Private" combines employer/union and individually purchased coverage. (E) Percentage of survey respondents in each race/ethnicity group who reported being unable to afford a visit to a medical specialist in the past 12 months (Healthcare Access & Utilization survey).

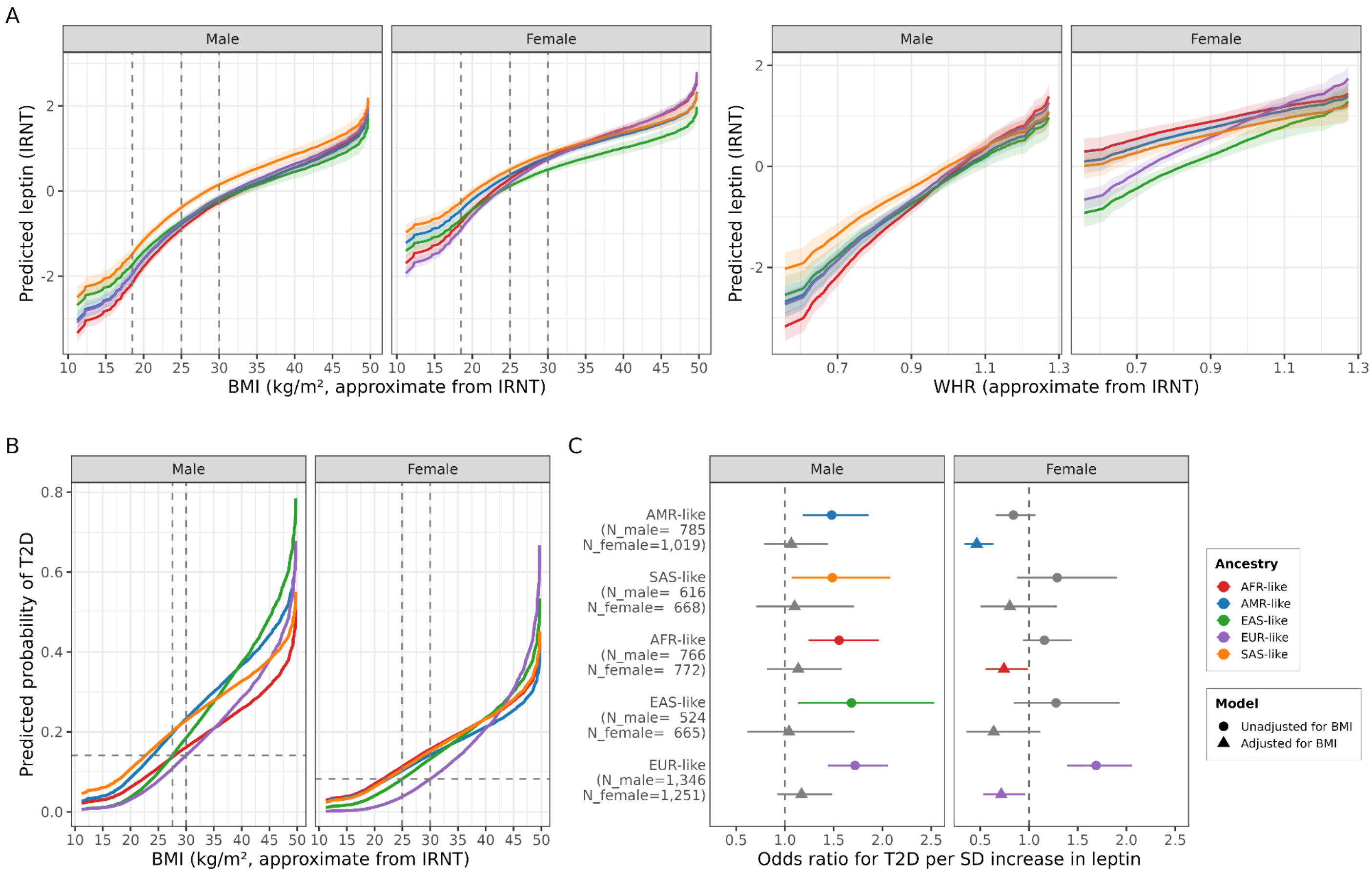


**Fig. S16. Cross-ancestry heterogeneity in body mass index (BMI)- and waist-to-hip ratio (WHR)-based associations with leptin in male and female.** (A) Predicted leptin levels across BMI (left) and WHR (right) by ancestry, based on interaction models. Vertical dashed lines indicate conventional BMI thresholds (18.5, 25, and 30 kg/m²). (B) Relationship between BMI and type 2 diabetes (T2D) risk, classified by ancestry groups. T2D risk was assessed in a logistic regression model, controlling for BMI, age, sex, biosample collection year, month, and time. The vertical dashed lines indicate the BMI thresholds needed to develop an equivalent risk of T2D in the White and Asian groups. (C) Association of leptin with T2D after BMI adjustment. Points show ancestry-specific estimates for the association of leptin with T2D before (circles) and after BMI adjustment (triangles). N shown on the y-axis corresponds to the BMI-adjusted model.

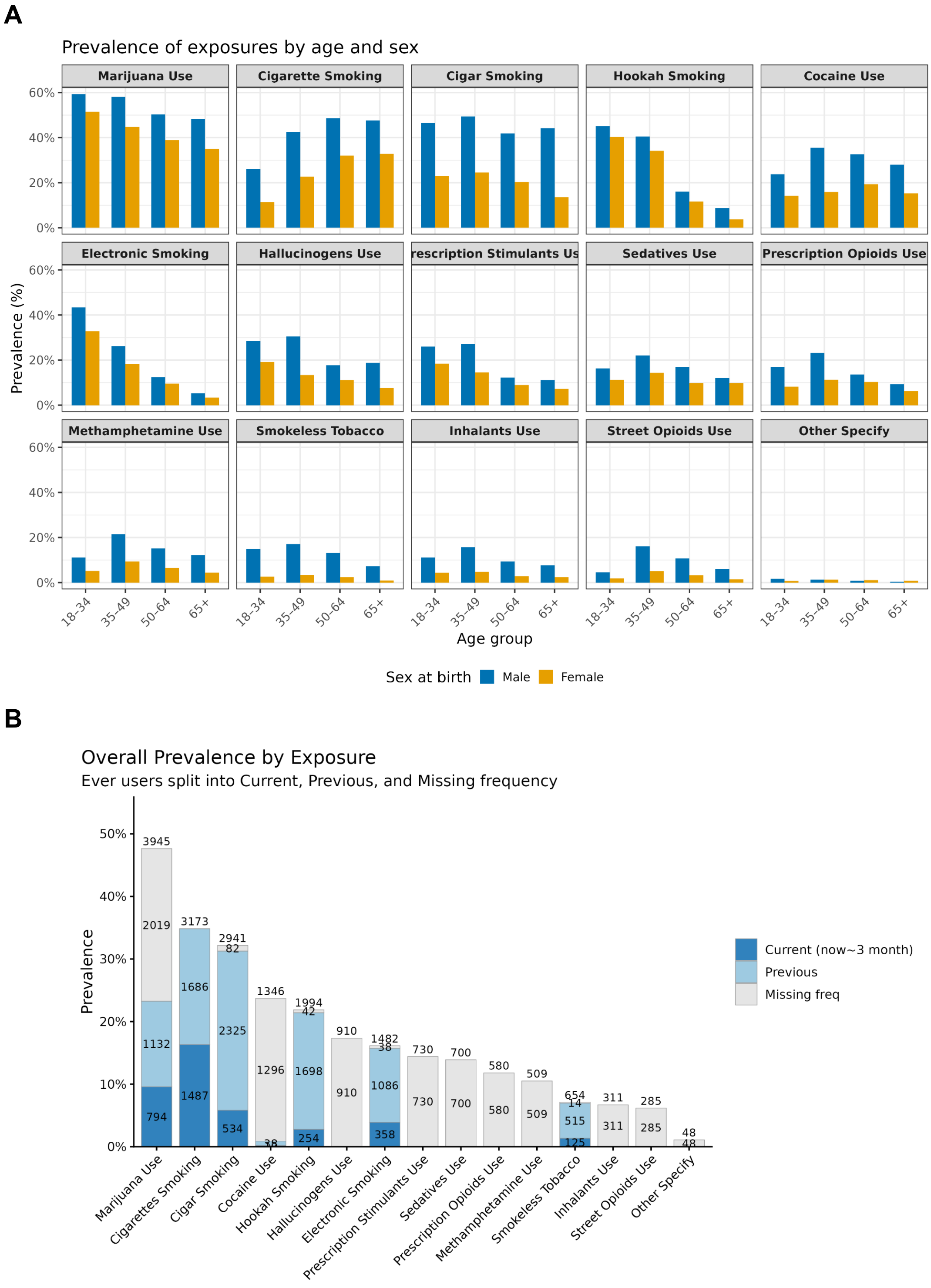


**Fig. S17.** A. Prevalence of tobacco/ substance use by age and sex. B. Overall prevalence of tobacco/ substance use by age and sex across socioeconomic status groups (tertile income, deprivation, and income).

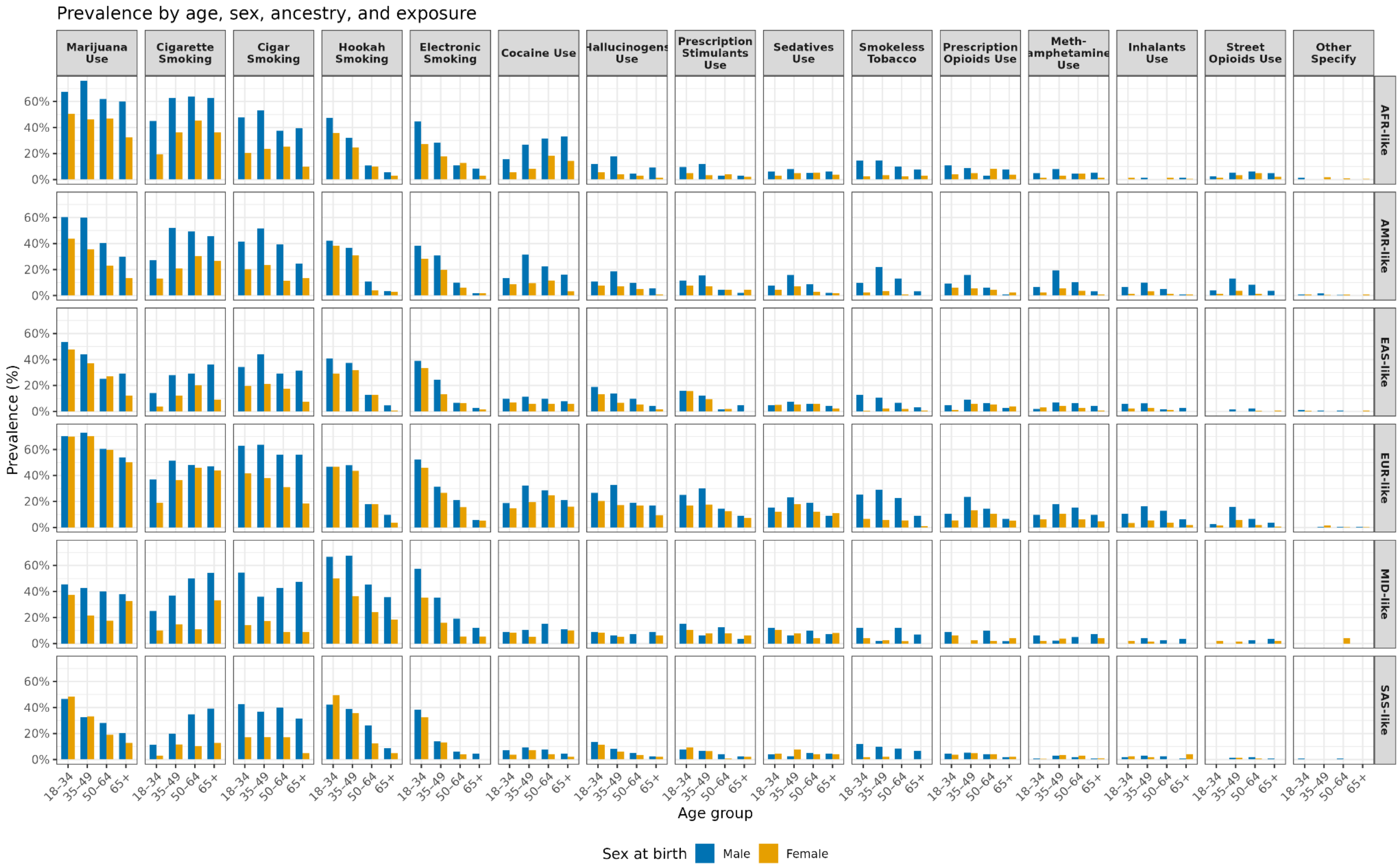


**Fig. S18. Prevalence of tobacco/ substance use by age and sex across genetic ancestry groups.**

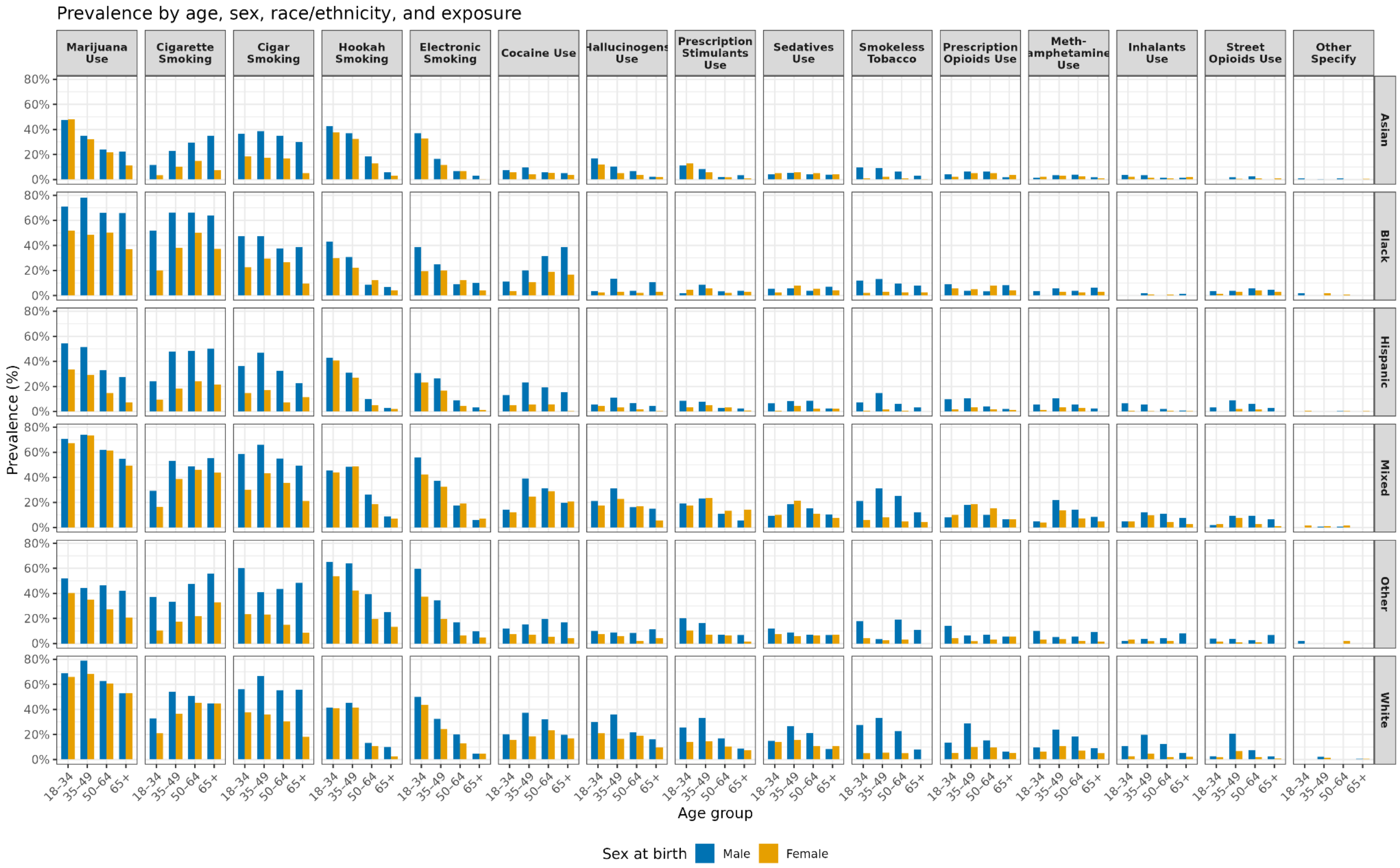


**Fig. S19. Prevalence of tobacco/ substance use by age and sex across self-reported race and/or ethnicity groups.**

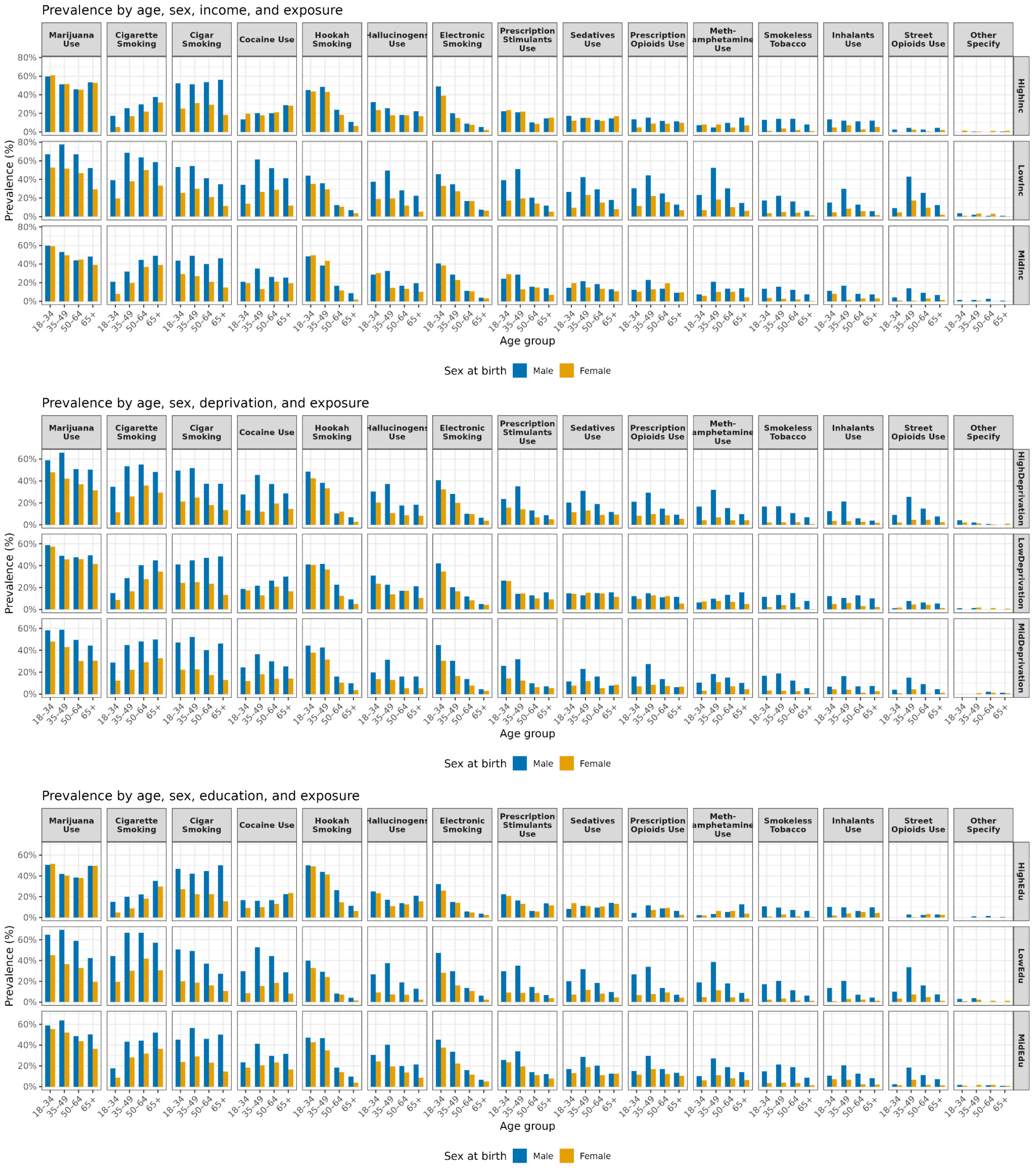


**Fig. S20. Prevalence of tobacco/ substance use by age and sex across socioeconomic status groups (tertile income, deprivation, and income).**

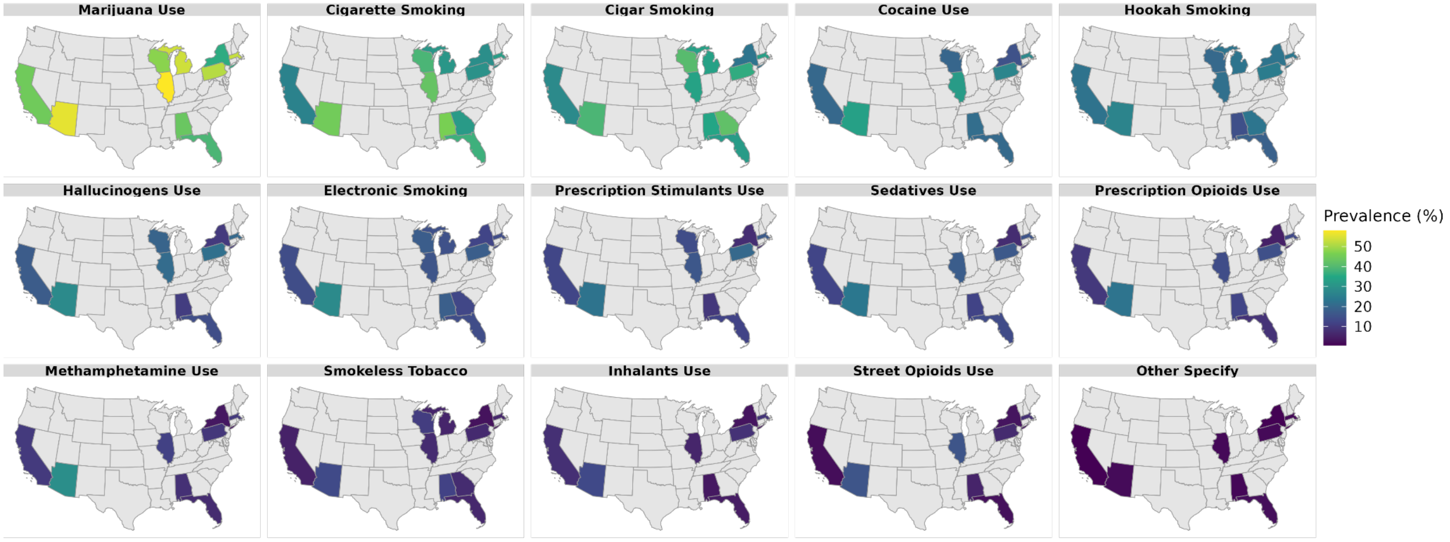


**Fig. S21. Geographic variation in exposure prevalence across U.S. regions. Color intensity shows the prevalence of tobacco/ substance use by states of residence in AoU proteomics data.**

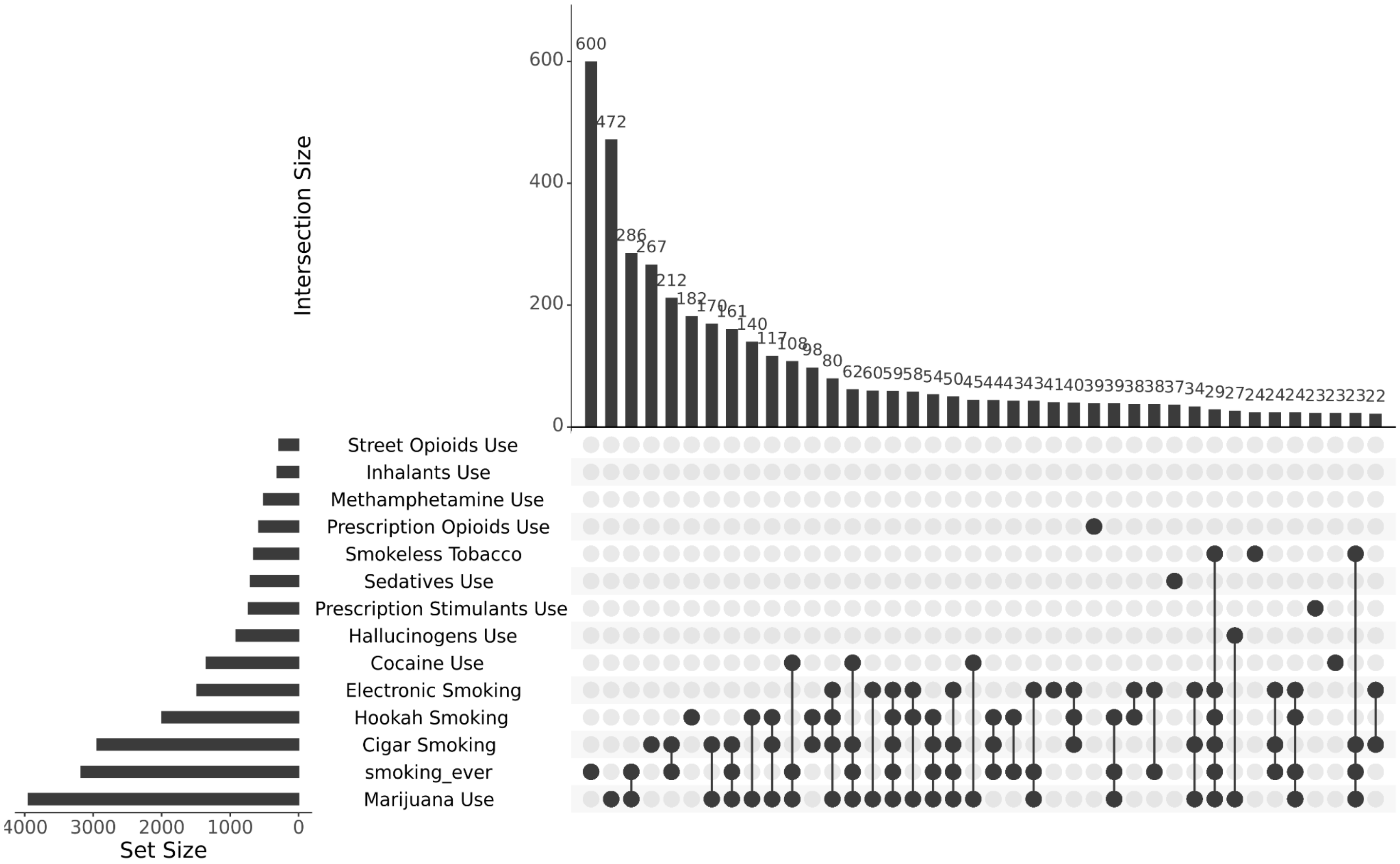


**Fig. S22. UpSet plot illustrating co-use patterns across multiple substance exposures in AoU proteomics set.**

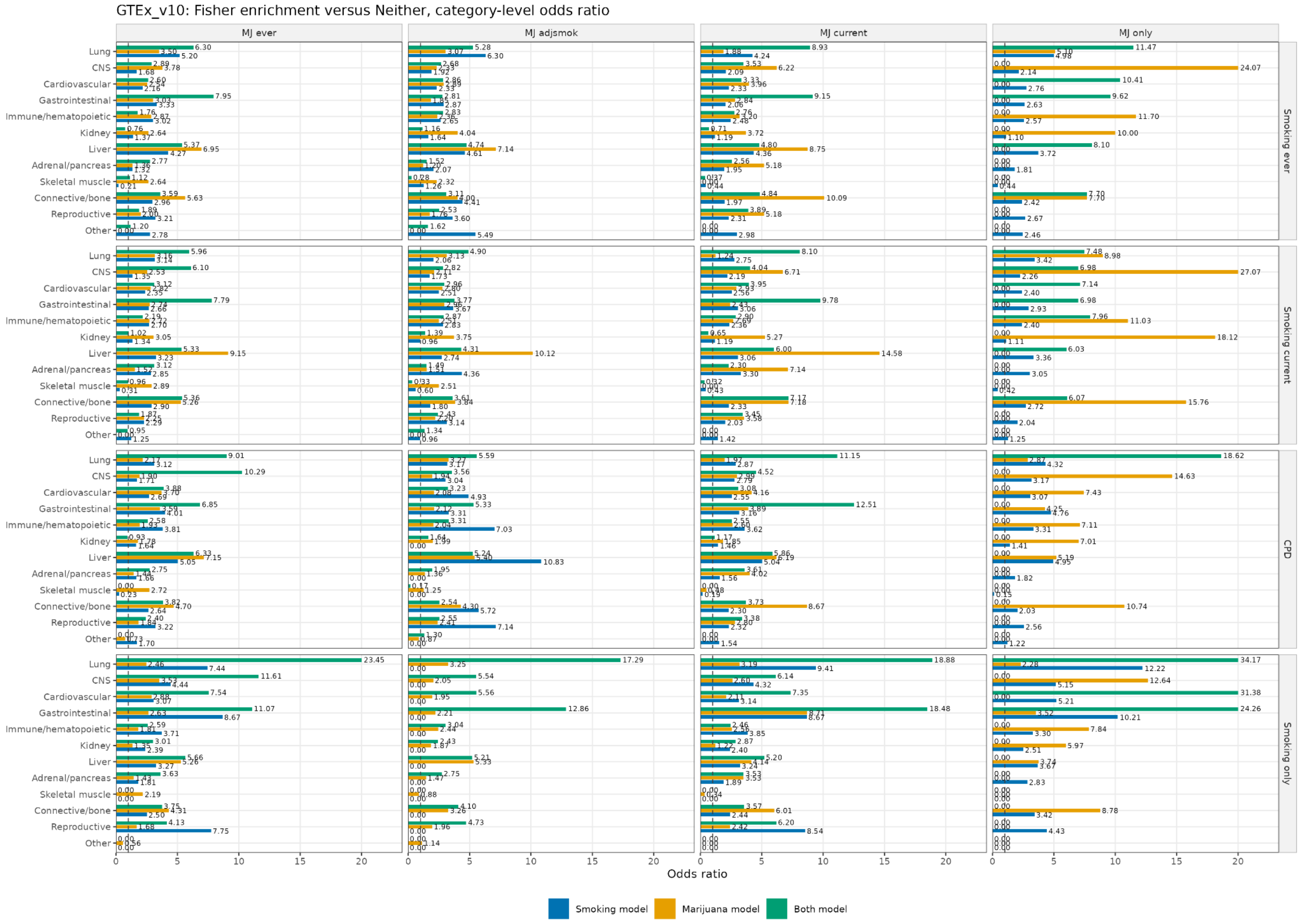


**Fig. S23. GTEx v10 tissue enrichment of proteins significant with either cigarette smoking or marijuana use.** Odds ratio. 4 tiers of cigarette smoking variables (row) is evaluated across ever smoking, current smoking, cigarette per day (CPD), and smoking-only. 4 tiers of marijuana use variables (column) is evaluated across ever use, marijuana use adjusted for smoking, current use, and marijuana-only. Each category shows the most significant tissue by FDR. Dashed line indicates OR = 1. Bars capped at OR = 20.

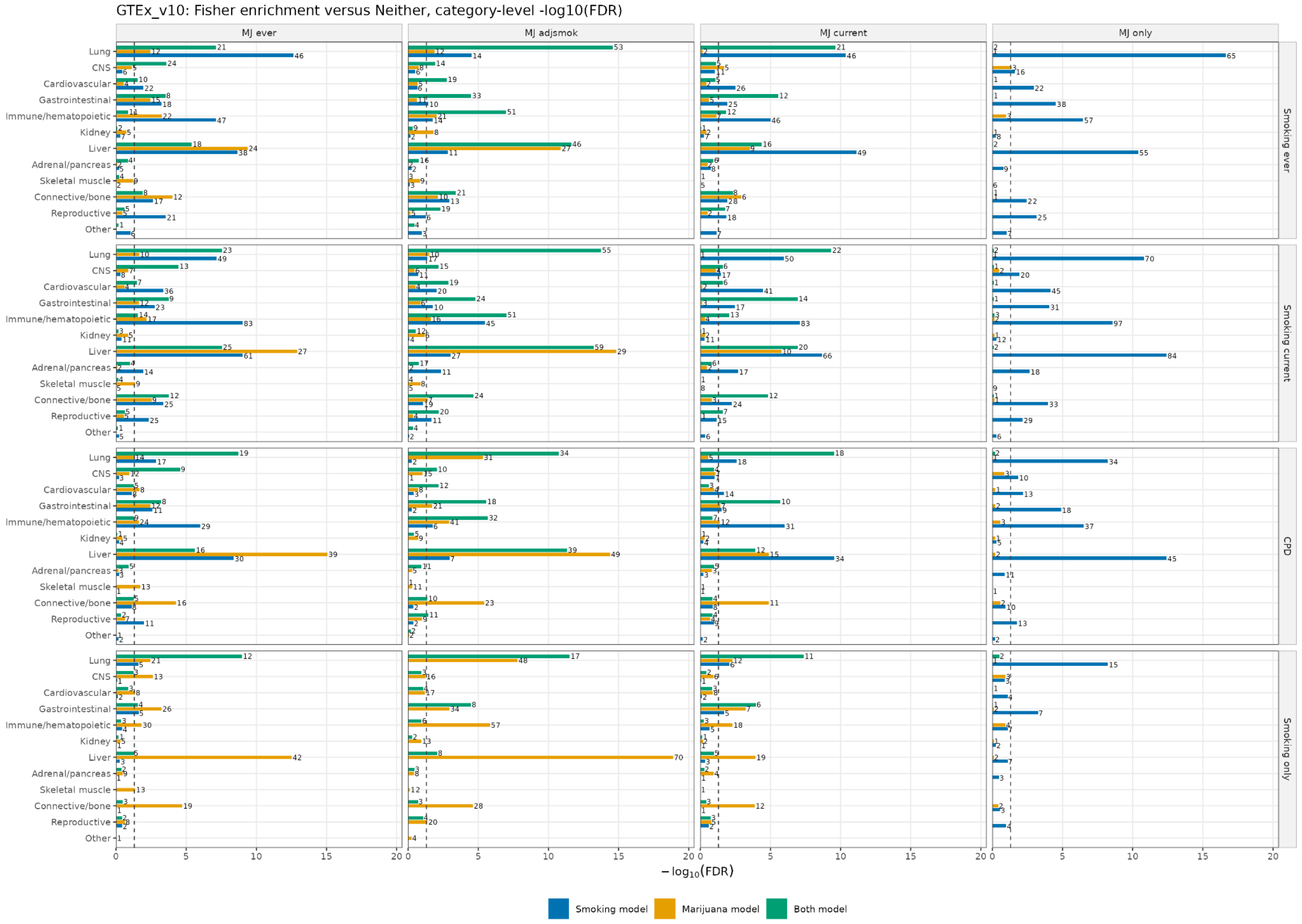


**Fig. S24. GTEx tissue enrichment of proteins significant with either cigarette smoking or marijuana use.** -log10(FDR). 4 tiers of cigarette smoking variables (row) is evaluated across ever smoking, current smoking, cigarette per day (CPD), and smoking-only. 4 tiers of marijuana use variables (column) is evaluated across ever use, marijuana use adjusted for smoking, current use, and marijuana-only. Each category shows the most significant tissue in that category. Text labels show target-group tissue-specific protein count.

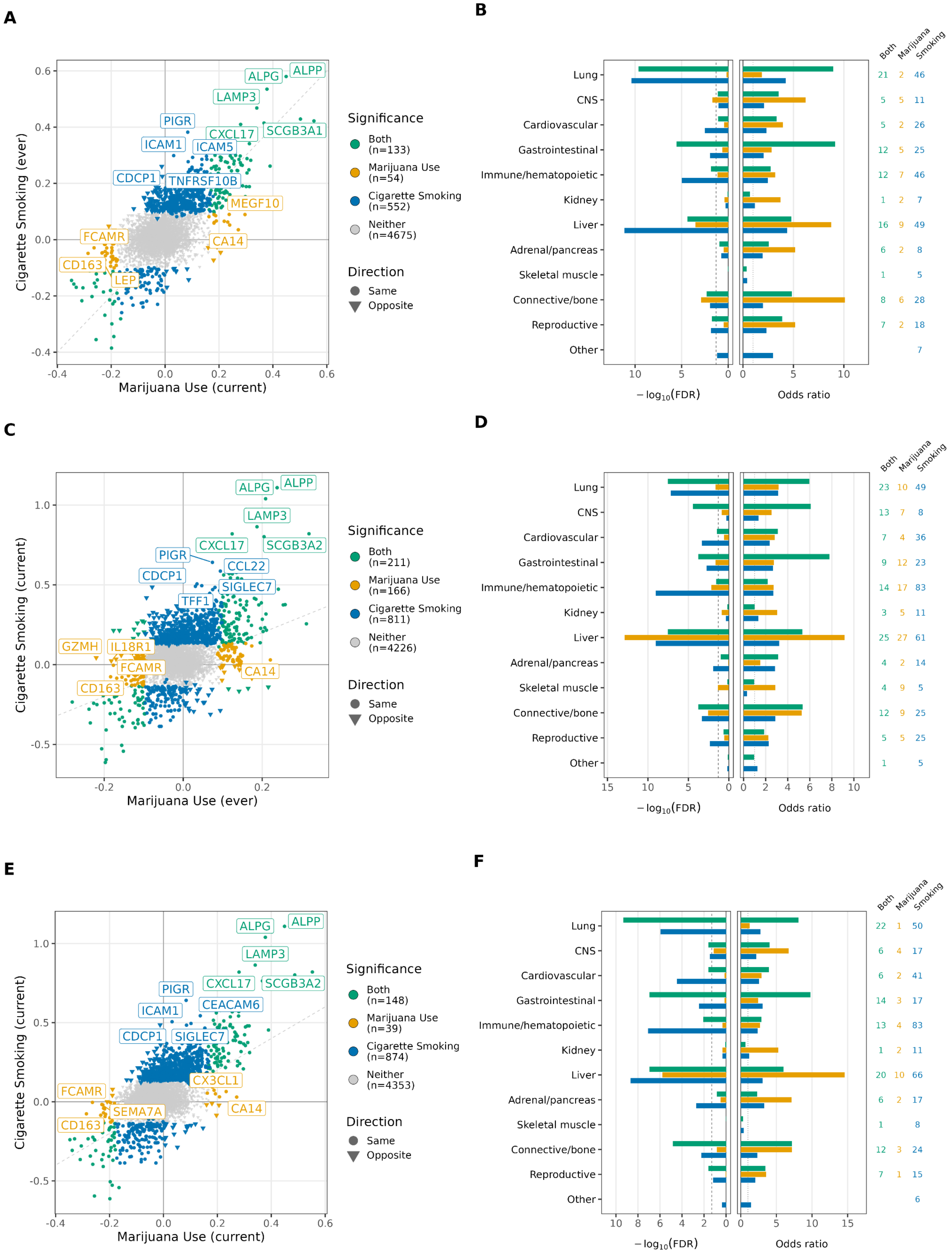
A
Cigarette Smoking (ever)
Marijuana Use (current)
ALPG
ALPP
LAMP3
PIGR
CXCL17
SCGB3A1
ICAM1
ICAM5
CDCP1
TNFRSF10B
MEGF10
FCAMR
CA14
CD163
LEP
Significance
Both (n=133)
Marijuana Use (n=54)
Cigarette Smoking (n=552)
Neither (n=4675)
Direction
Same
Opposite
B
Both
Marijuana
Smoking
Lung
CNS
Cardiovascular
Gastrointestinal
Immune/hematopoietic
Kidney
Liver
Adrenal/pancreas
Skeletal muscle
Connective/bone
Reproductive
Other
$-\log_{10}(\mathrm{FDR})$
Odds ratio
C
Cigarette Smoking (current)
Marijuana Use (ever)
ALPG
ALPP
LAMP3
CXCL17
SCGB3A2
PIGR
CCL22
CDCP1
SIGLEC7
TFF1
GZMH
IL18R1
FCAMR
CA14
CD163
Significance
Both (n=211)
Marijuana Use (n=166)
Cigarette Smoking (n=811)
Neither (n=4226)
Direction
Same
Opposite
D
Both
Marijuana
Smoking
$-\log_{10}(\mathrm{FDR})$
Odds ratio
E
Cigarette Smoking (current)
Marijuana Use (current)
ALPG
ALPP
LAMP3
PIGR
CXCL17
SCGB3A2
ICAM1
CEACAM6
CDCP1
SIGLEC7
CX3CL1
FCAMR
SEMA7A
CA14
CD163
Significance
Both (n=148)
Marijuana Use (n=39)
Cigarette Smoking (n=874)
Neither (n=4353)
Direction
Same
Opposite
F
Both
Marijuana
Smoking
$-\log_{10}(\mathrm{FDR})$
Odds ratio

**Fig. S25. Classification of the protein effects of marijuana use and cigarette smoking.** (left panel) Comparison of beta coefficients from linear regression models of current marijuana use (x axis) and current cigarette smoking (y axis) for top-associated proteins ($P_{bonf} < 0.05$). Proteins are colored by classification: cigarette smoking-predominant effects (blue), marijuana use-predominant effects (yellow), and effects shared between both exposures (green) (table S17). Gray points indicate proteins not significantly associated with either exposure ($P_{bonf} \geq 0.05$). (right panel) Tissue enrichment of the three protein sets from panel D (cigarette smoking-only, marijuana use-only, and shared) relative to proteins associated with neither exposure, using the same GTEx v10/TissueEnrich-based approach and Fisher's exact testing described in panel D (table S18). **A-B.** Marijuana Use (Current) vs. Cigarette smoking (Ever). **C-D.** Marijuana Use (Ever) vs. Cigarette smoking (Current). **E-F.** Marijuana Use (Current) vs. Cigarette smoking (Current).

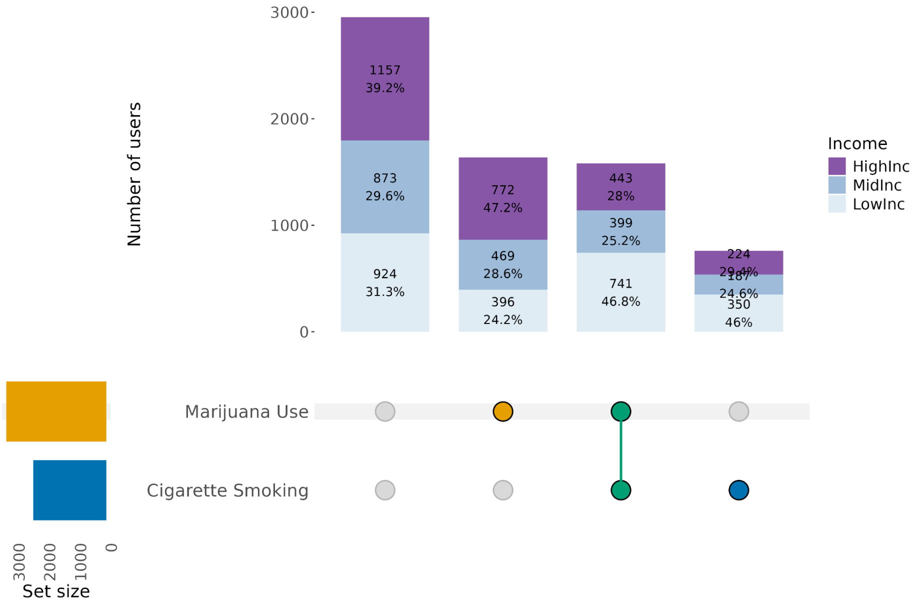
Number of users
3000
2000
1000
0
1157
39.2%
873
29.6%
924
31.3%
772
47.2%
469
28.6%
396
24.2%
443
28%
399
25.2%
741
46.8%
224
29.4%
187
24.6%
350
46%
Income
HighInc
MidInc
LowInc
Marijuana Use
Cigarette Smoking
3000
2000
1000
0
Set size

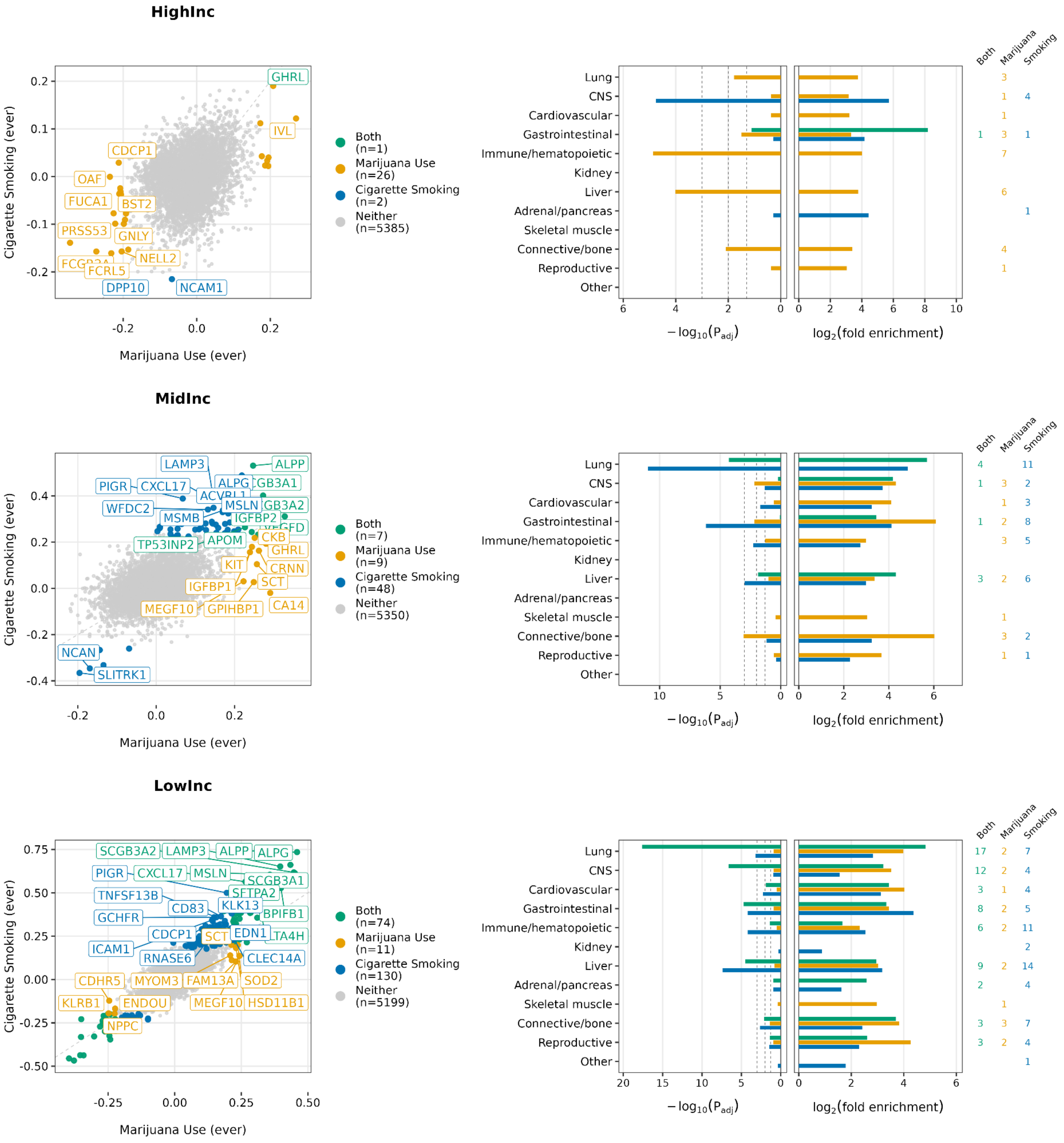
HighInc
Cigarette Smoking (ever)
Marijuana Use (ever)
GHRL
IVL
CDCP1
OAF
FUCA1
BST2
PRSS53
GNLY
NELL2
FCRL5
DPP10
NCAM1
Both (n=1)
Marijuana Use (n=26)
Cigarette Smoking (n=2)
Neither (n=5385)
Lung
CNS
Cardiovascular
Gastrointestinal
Immune/hematopoietic
Kidney
Liver
Adrenal/pancreas
Skeletal muscle
Connective/bone
Reproductive
Other
$-\log_{10}(P_{adj})$
$\log_2$(fold enrichment)
Both
Marijuana
Smoking
MidInc
LAMP3
ALPP
PIGR
CXCL17
ALPG
WFDC2
MSLN
MSMB
IGFBP2
TP53INP2
APOM
CKB
GHRL
KIT
CRNN
IGFBP1
SCT
MEGF10
GPIHBP1
CA14
NCAN
SLITRK1
Both (n=7)
Marijuana Use (n=9)
Cigarette Smoking (n=48)
Neither (n=5350)
LowInc
SCGB3A2
LAMP3
ALPP
ALPG
PIGR
CXCL17
MSLN
SCGB3A1
TNFSF13B
SFTPA2
CD83
KLK13
GCHFR
BPIFB1
CDCP1
SCT
EDN1
ICAM1
RNASE6
CLEC14A
CDHR5
MYOM3
FAM13A
SOD2
KLRB1
ENDOU
MEGF10
HSD11B1
NPPC
Both (n=74)
Marijuana Use (n=11)
Cigarette Smoking (n=130)
Neither (n=5199)

**Fig. S26. Classification of the protein effects of marijuana use and cigarette smoking across income groups.**

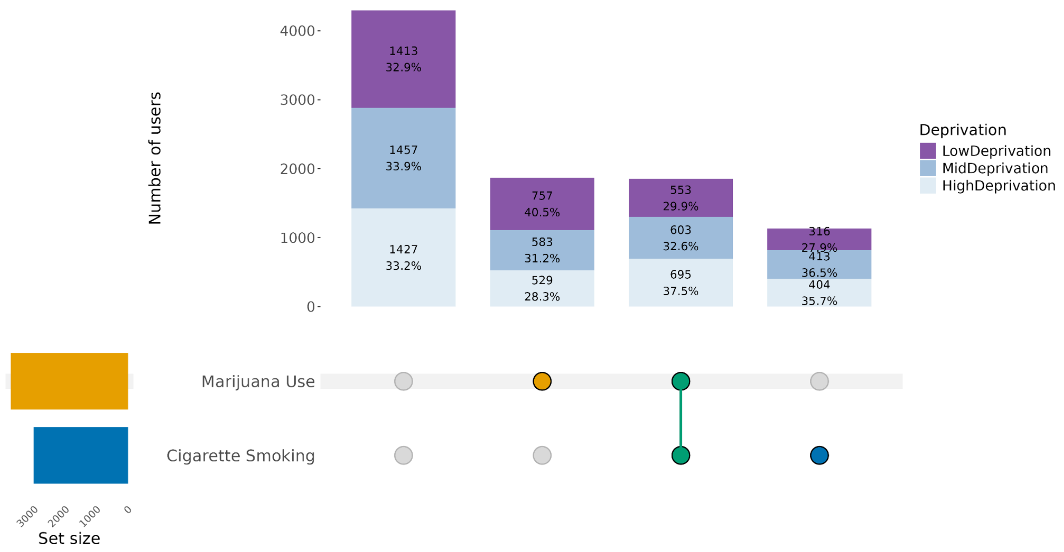
Number of users
4000
3000
2000
1000
0
1413
32.9%
1457
33.9%
1427
33.2%
757
40.5%
583
31.2%
529
28.3%
553
29.9%
603
32.6%
695
37.5%
316
27.9%
413
36.5%
404
35.7%
Deprivation
LowDeprivation
MidDeprivation
HighDeprivation
Marijuana Use
Cigarette Smoking
3000
2000
1000
0
Set size

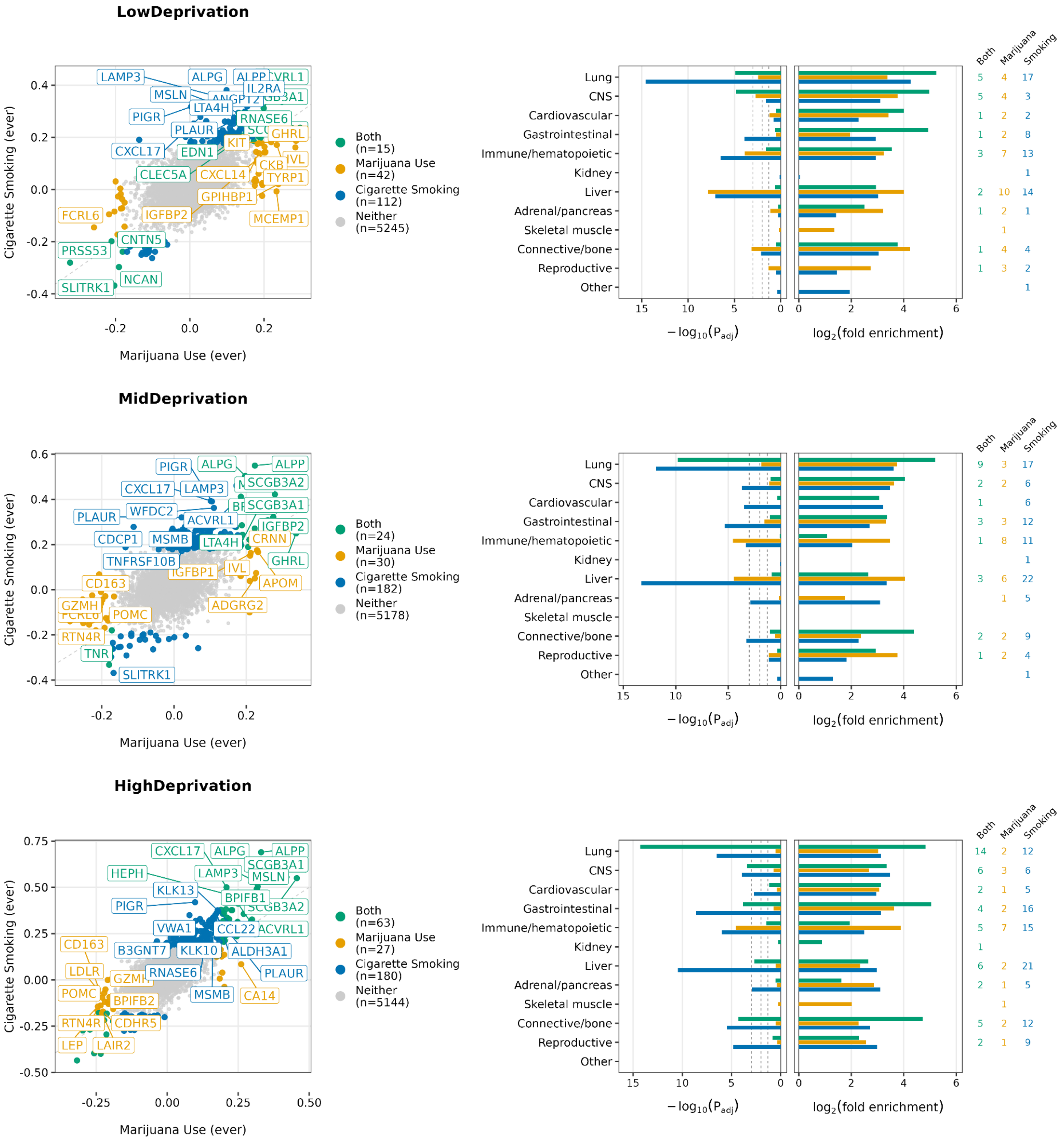

LowDeprivation
Cigarette Smoking (ever)
Marijuana Use (ever)
LAMP3
ALPG
ALPP
ACVRL1
MSLN
IL2RA
LTA4H
PIGR
RNASE6
PLAUR
GHRL
CXCL17
EDN1
KIT
CKB
IVL
CLEC5A
CXCL14
TYRP1
GPIHBP1
FCRL6
IGFBP2
MCEMP1
PRSS53
CNTN5
SLITRK1
NCAN
Both
(n=15)
Marijuana Use
(n=42)
Cigarette Smoking
(n=112)
Neither
(n=5245)
Lung
CNS
Cardiovascular
Gastrointestinal
Immune/hematopoietic
Kidney
Liver
Adrenal/pancreas
Skeletal muscle
Connective/bone
Reproductive
Other
$-\log_{10}(P_{adj})$
$\log_2$(fold enrichment)
Both
Marijuana
Smoking
MidDeprivation
PIGR
ALPG
ALPP
CXCL17
LAMP3
SCGB3A2
PLAUR
WFDC2
SCGB3A1
ACVRL1
IGFBP2
CDCP1
MSMB
LTA4H
CRNN
TNFRSF10B
GHRL
IGFBP1
IVL
CD163
APOM
GZMH
POMC
ADGRG2
RTN4R
TNR
SLITRK1
Both
(n=24)
Marijuana Use
(n=30)
Cigarette Smoking
(n=182)
Neither
(n=5178)
HighDeprivation
CXCL17
ALPG
ALPP
SCGB3A1
HEPH
LAMP3
MSLN
KLK13
BPIFB1
PIGR
SCGB3A2
VWA1
CCL22
ACVRL1
CD163
B3GNT7
KLK10
ALDH3A1
LDLR
RNASE6
PLAUR
POMC
MSMB
CA14
BPIFB2
RTN4R
CDHR5
LEP
LAIR2
Both
(n=63)
Marijuana Use
(n=27)
Cigarette Smoking
(n=180)
Neither
(n=5144)

**Fig. S27. Classification of the protein effects of marijuana use and cigarette smoking across deprivation groups.**

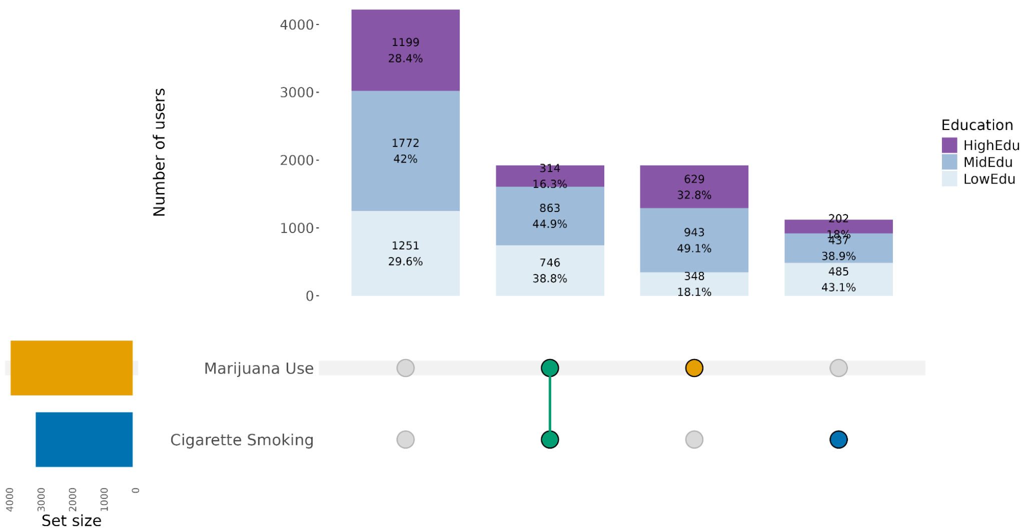
Number of users
4000
3000
2000
1000
0
1199
28.4%
1772
42%
1251
29.6%
314
16.3%
863
44.9%
746
38.8%
629
32.8%
943
49.1%
348
18.1%
202
18%
437
38.9%
485
43.1%
Education
HighEdu
MidEdu
LowEdu
Marijuana Use
Cigarette Smoking
Set size

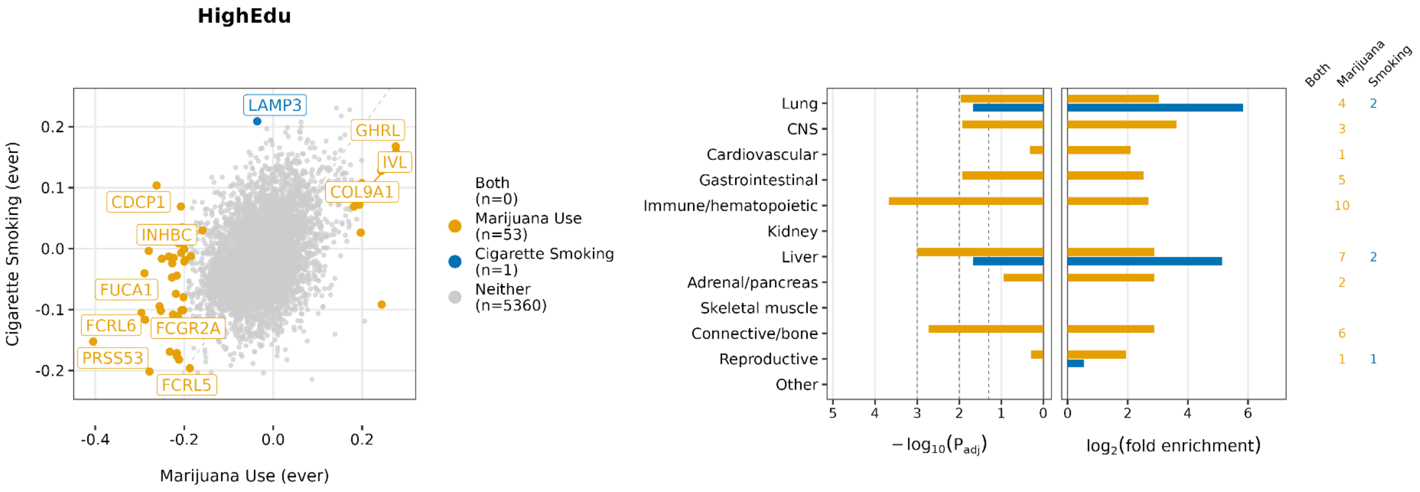
HighEdu
Cigarette Smoking (ever)
Marijuana Use (ever)
LAMP3
GHRL
IVL
COL9A1
CDCP1
INHBC
FUCA1
FCRL6
FCGR2A
PRSS53
FCRL5
Both (n=0)
Marijuana Use (n=53)
Cigarette Smoking (n=1)
Neither (n=5360)
Lung
CNS
Cardiovascular
Gastrointestinal
Immune/hematopoietic
Kidney
Liver
Adrenal/pancreas
Skeletal muscle
Connective/bone
Reproductive
Other
$-\log_{10}(P_{adj})$
$\log_2$(fold enrichment)
Both
Marijuana
Smoking

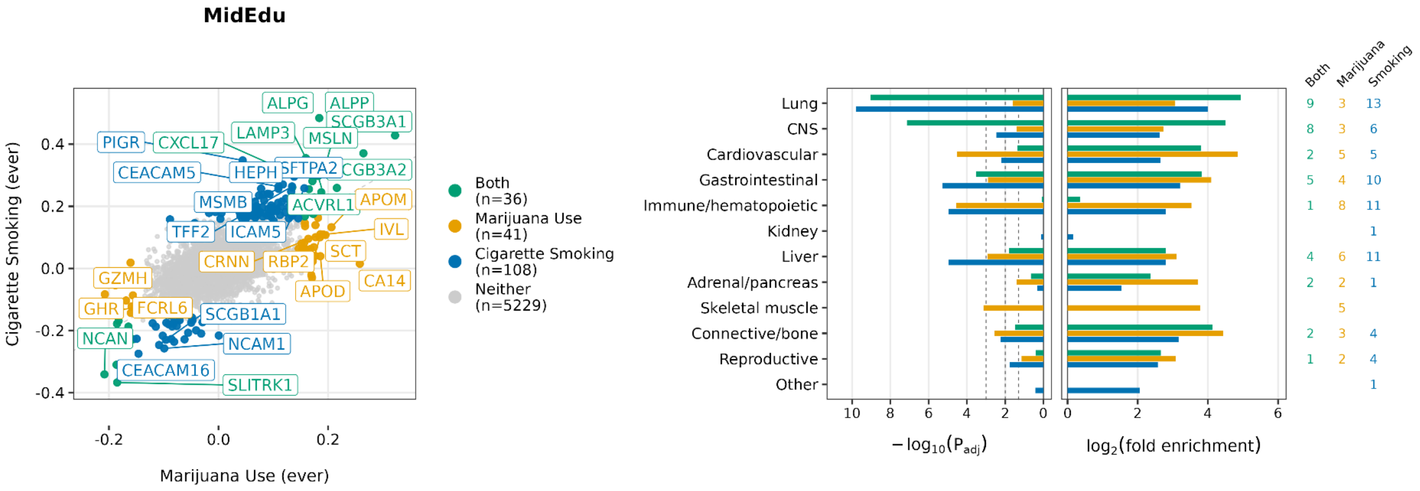
MidEdu
Cigarette Smoking (ever)
Marijuana Use (ever)
ALPG
ALPP
SCGB3A1
PIGR
CXCL17
LAMP3
MSLN
CEACAM5
HEPH
SFTPA2
SCGB3A2
MSMB
ACVRL1
APOM
TFF2
ICAM5
IVL
CRNN
RBP2
SCT
GZMH
APOD
CA14
GHR
FCRL6
SCGB1A1
NCAN
NCAM1
CEACAM16
SLITRK1
Both (n=36)
Marijuana Use (n=41)
Cigarette Smoking (n=108)
Neither (n=5229)
Lung
CNS
Cardiovascular
Gastrointestinal
Immune/hematopoietic
Kidney
Liver
Adrenal/pancreas
Skeletal muscle
Connective/bone
Reproductive
Other
$-\log_{10}(P_{adj})$
$\log_2$(fold enrichment)
Both
Marijuana
Smoking

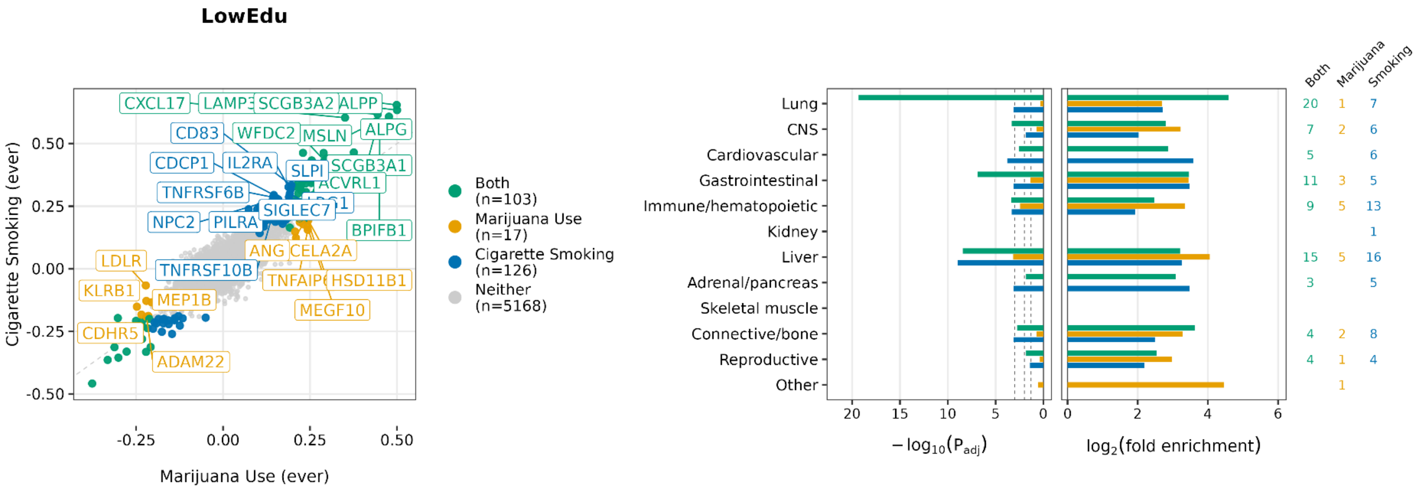
LowEdu
Cigarette Smoking (ever)
Marijuana Use (ever)
CXCL17
SCGB3A2
ALPP
CD83
WFDC2
MSLN
ALPG
CDCP1
IL2RA
SLPI
SCGB3A1
TNFRSF6B
ACVRL1
SIGLEC7
NPC2
PILRA
BPIFB1
ANG
CELA2A
LDLR
TNFRSF10B
HSD11B1
KLRB1
MEP1B
MEGF10
CDHR5
ADAM22
Both (n=103)
Marijuana Use (n=17)
Cigarette Smoking (n=126)
Neither (n=5168)
Lung
CNS
Cardiovascular
Gastrointestinal
Immune/hematopoietic
Kidney
Liver
Adrenal/pancreas
Skeletal muscle
Connective/bone
Reproductive
Other
$-\log_{10}(P_{adj})$
$\log_2$(fold enrichment)
Both
Marijuana
Smoking

**Fig. S28. Classification of the protein effects of marijuana use and cigarette smoking across education groups.**

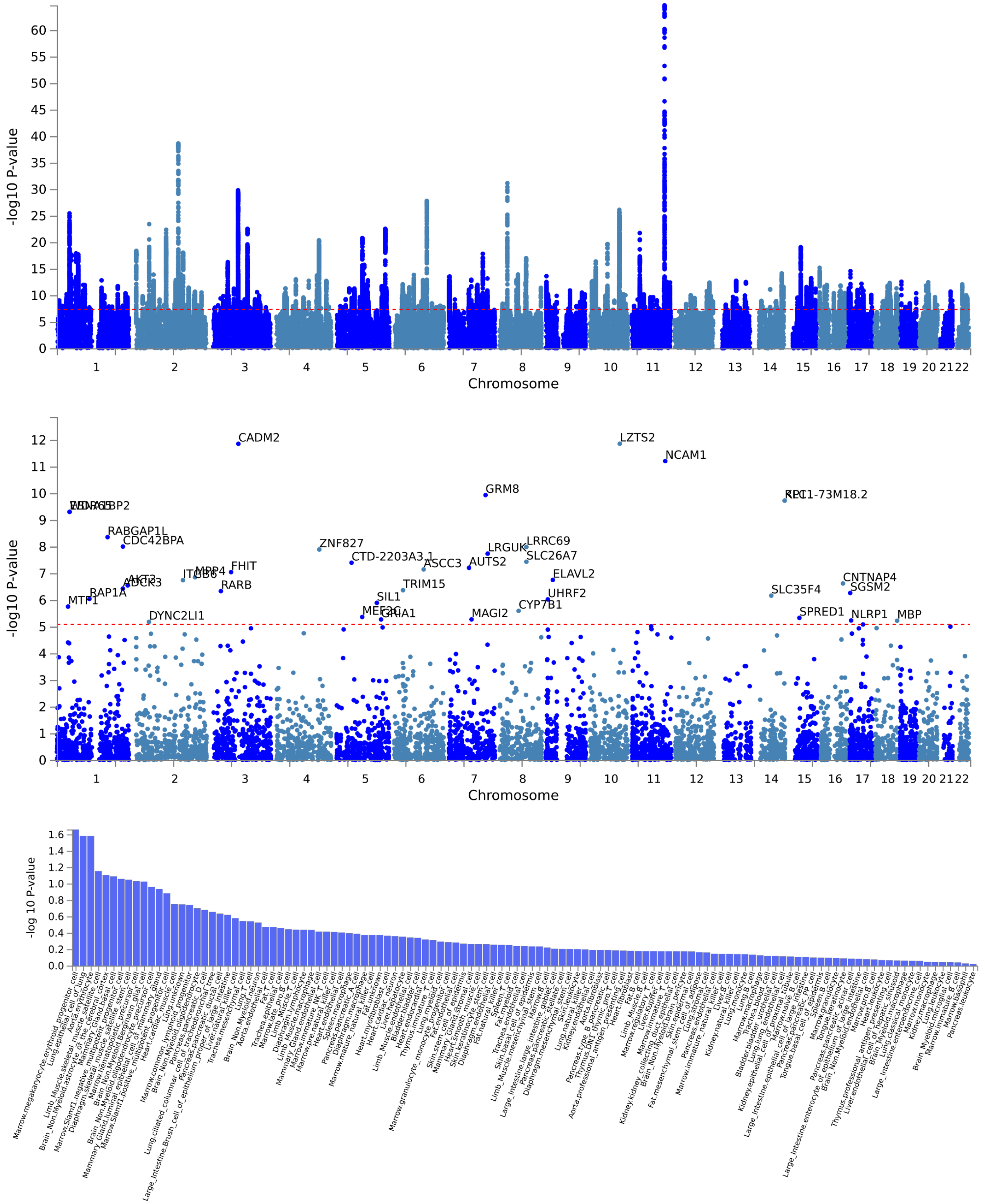


**Fig. S29. Genome-wide association study of smoking initiation from the Saunders et al. (2022).**

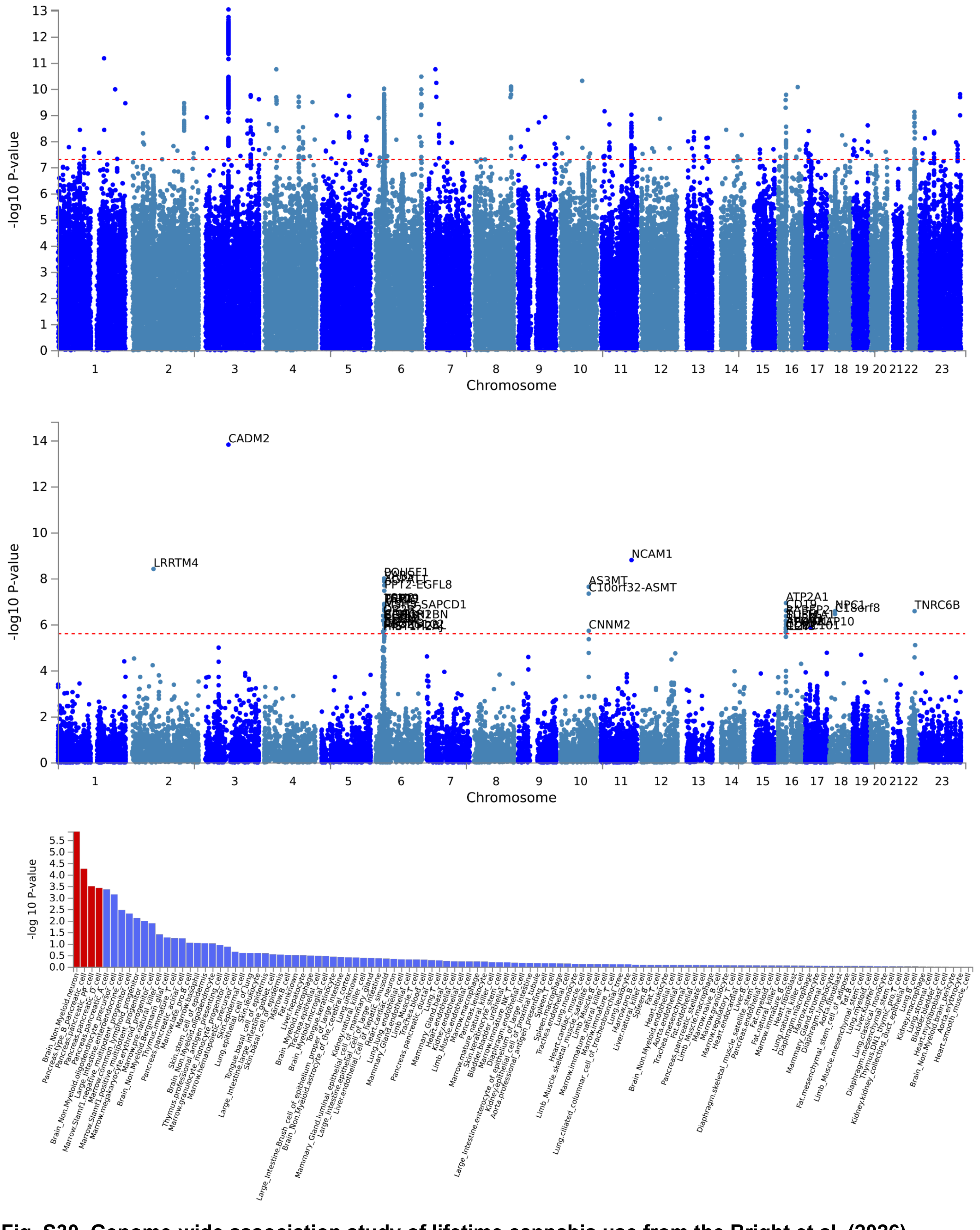


**Fig. S30. Genome-wide association study of lifetime cannabis use from the Bright et al. (2026).**
