## Supplementary text for "Proteomic Signatures Capture Environmental Exposures Across Diverse Populations"

#### Proteins associate with many exposures with varying specificity

After quality control of the All of Us Olink HT 5k proteomic data (Methods), 9,561 samples and 5,414 proteins were retained for analysis (**Fig. 1, fig. S1**, **table S1**). We tested protein associations with 108 health-related exposure variables spanning behavioral, socioeconomic, environmental, anthropometric, clinical and biomarker domains (Methods; **table S2**), identifying 62,042 significant protein-exposure pairs at a Bonferroni-corrected threshold. In the analysis pooled across all samples, the proportion of significant proteins following roughly: anthropometric (adiposity) (mean=33.2%, SD=22.3), biomarker subclasses (blood pressure: mean=29.4%, SD=1.6), ambient temperature (mean=18.9%, SD=1.4), SES (mean=7.8%, SD=3.3), air pollution (mean=7.1%, SD=6.2), and Tobacco/Substance (mean=2.7%, SD=4.0), reflecting the precision of trait measures and degree of direct impact on biology (**Fig. 2A, table S3**). About half of the measured proteins are associated with age (49.4%) and anthropometric traits such as BMI (47.7%), followed by sex (34.5%). 18.9% of proteins were associated with current cigarette smoking and 9.6% with frequent alcohol drinking (4+/wk); these percentages are smaller than reported in UKB (e.g., 50% for smoking (*1*)), likely due to power differences, as downsampling in AoU revealed a consistent decrease. Supporting this interpretation, downsampling the full AoU never/current-smoker cohort to n=1,000 (matching the never:current ratio) reduced the proportion of significant proteins to a mean of 2.5% (SD=1.0%; 5th-95th percentile across repeated subsamples: 1.5%-3.6%), a similarly small fraction as observed in UKB, suggesting sample size — rather than a fundamentally different biological effect — largely explains the discrepancy between cohorts. Up to 20% of proteins were associated with ambient temperature (365-day average temperature: 20.2%) and air pollution ($NO_2$: 23%, $PM_{2.5}$: 7.2%). SES variables like income, deprivation, and education are associated with the fewer proteins (11.1, 8.2, and 3.3%, respectively).

We first assessed whether circulating proteins capture molecular signatures of environmental variation beyond broad geographic confounding (**Fig. 1**). For most geocoded exposures, adjustment for region, defined by participants' state of residence and categorized as Northeast, Midwest, West, or South, substantially attenuated the proportion of significant protein associations (**fig. S2, table S3**), reducing the number of significant protein associations by an average of 87.8% for ambient temperature, 30.5% for SES, and 15.3% for air pollution exposures. This pattern is consistent with a major contribution from geographical structure to these signals (Methods). In contrast, $PM_{2.5}$-associated proteins increased after regional adjustment, from 390 to 491 significant proteins for 365-day average $PM_{2.5}$ and from 143 to 158 significant proteins for 30-day average $PM_{2.5}$. This pattern suggests that $PM_{2.5}$-protein associations were not explained by broad regional differences alone. Rather, adjustment for region may have reduced between-region heterogeneity that otherwise masked within-region $PM_{2.5}$-associated proteomic variation.

Several proteins remained associated with $PM_{2.5}$ (365d avg) after regional adjustment, including proteins with prior evidence linking them to respiratory disease biology. For example, *SERPINA1*–which encodes alpha-1 antitrypsin, a protein whose deficiency is a well-established genetic cause of rare forms of COPD and emphysema–was inversely associated with PM2.5 exposure ($\beta$ = -0.056, SE = 0.012, $P_{bonf}$ = 0.006) (*2*) (**table S4**). *IL1RL1*, which encodes ST2, a component of the IL-33 signaling pathway, has nearby asthma risk variants that reduce soluble ST2 and promote type 2 airway inflammation, was similarly inversely associated with $PM_{2.5}$ ($\beta$ = -0.077, SE = 0.011, $P_{bonf}$ = $3.90\times10^{-09}$) (*3*). *CHI3L1* (YKL-40), a marker of airway inflammation and remodeling and previously associated with both asthma and COPD, was also inversely associated with $PM_{2.5}$ ($\beta$ = -0.055, SE = 0.010, $P_{bonf}$ = 0.0002) (*4*). Finally, *IL6R*–encoding a key inflammatory pathway implicated in asthma, with rs2228145 influencing soluble IL-6 receptor levels and IL-6 trans-signaling–showed a similar inverse association (β = -0.077, SE = 0.011, $P_{bonf}$ = 1.00×$10^{-7}$) (*5*). These findings suggest that the $PM_{2.5}$-associated proteomic signature may capture biologically relevant inflammatory and airway-related variation beyond broad regional structure. However, without formal causal inference testing, these associations should be interpreted as molecular correlates of $PM_{2.5}$ exposure rather than evidence of mediation.

We also observed region-adjusted associations for ambient temperature exposures involving proteins with prior links to heat stress, heat acclimation, and temperature-related pathophysiology. For average warm-season temperature, positive associations after regional adjustment were observed for C1QTNF1 (β = 0.093, SE = 0.014, $P_{bonf}$ = 5.32×$10^{-8}$), HSPB6 (β = 0.064, SE = 0.012, $P_{bonf}$ = 1.50×$10^{-4}$), HSPA13 (β = 0.070, SE = 0.014, $P_{bonf}$ = 2.66×$10^{-3}$), and SMOC1 (β = 0.057, SE = 0.012, $P_{bonf}$ = 1.27×$10^{-2}$), while MOG was inversely associated after regional adjustment (β = -0.068, SE = 0.012, $P_{bonf}$ = 1.67×$10^{-4}$) (**table S4**). Notably, C1QTNF1, MOG, and SMOC1 overlap with proteins previously highlighted in human heat-stress and heat-acclimation proteomic studies, including proteins reported to change during heat stress after acclimation and to show enrichment in brain- or arterial-tissue expression contexts(*6*). Although HSPA13 and HSPB6 are heat-shock-related proteins, HSPA1A/Hsp70 and HSPB1/Hsp27 have more established roles as canonical inducible heat-shock chaperones regulated within the cellular heat-shock response(*7*–*9*). In our analyses, HSPA1A and HSPB1 showed Bonferroni-significant associations with ambient temperature variables before regional adjustment, including HSPA1A with average cold-season temperature (β = 0.0819, SE = 0.011, $P_{bonf}$ = 3.12×$10^{-09}$), 365-day average temperature (β = 0.074, SE = 0.011, $P_{bonf}$ = 1.52×$10^{-07}$), and 30-day average temperature (β = 0.094, SE = 0.017, $P_{bonf}$ = 7.48×$10^{-05}$), and HSPB1 with average warm-season temperature (β = 0.073, SE = 0.011, $P_{bonf}$ = 4.51×$10^{-07}$). However, these canonical heat-shock protein associations were not among the region-adjusted Bonferroni-significant findings. These results suggest that adjustment for region attenuated some canonical heat-shock protein associations, while a subset of warm-season temperature-associated proteins remained robust to regional adjustment and included proteins previously implicated in heat acclimation, vascular remodeling, extracellular matrix biology, and tissue-specific adaptation to heat exposure.

Second, we next estimated group-stratified exposure-protein associations and tested for heterogeneity (**Fig. 1**) to evaluate whether protein effects differed across genetically inferred ancestry groups (EUR-like, AMR-like, AFR-like, EAS-like, and SAS-like) or self-reported race and/or ethnicity groups (White, Hispanic, Black, Asian, and Mixed) (**Fig. 2B, fig. S3, table S5**). Overall, evidence of heterogeneity across ancestry and race/ethnicity strata was limited. Across more than 100 exposures, fewer than 1% of proteins per exposure, on average, were both significant in the fixed-effects meta-analysis (Bonferroni-adjusted $P < 0.05$) and heterogeneous across groups according to Cochran's Q test ($P < 0.05/3{,}011$ independent proteins), with a median proportion of 0.06% in both genetically inferred ancestry and self-reported race/ethnicity-stratified analyses. Furthermore, the proportion of heterogeneous associations did not differ significantly between the two stratification approaches (paired Wilcoxon signed-rank test, $P = 0.173$; **fig. S4**).

To evaluate cross-biobank transferability of protein scores, we next evaluated whether UK Biobank-trained protein-exposure scores generalized to the multi-ancestry All of Us cohort (**Fig. 2C, table S6, fig. S5 to S6**). After adjustment for age, sex at birth, collection year, month of sample collection, and time of day of sample collection, the UKB-trained current smoking score showed strong discrimination of current smoking status across AoU ancestry groups. Performance was highest in EUR-like participants (point-biserial $r = 0.735$, 95% CI: 0.694-0.771; AUC = 0.950, 95% CI: 0.927-0.974), followed by AFR-like ($r = 0.616$, 95% CI: 0.549-0.675; AUC = 0.899, 95% CI: 0.866-0.932), AMR-like ($r = 0.564$, 95% CI: 0.497-0.624; AUC = 0.880, 95% CI: 0.836-0.923), SAS-like ($r = 0.471$, 95% CI: 0.386-0.548; AUC = 0.899, 95% CI: 0.825-0.972), and EAS-like ($r = 0.344$, 95% CI: 0.241-0.440; AUC = 0.909, 95% CI: 0.782-1.000). These findings demonstrate broad transferability of the UKB-trained smoking score, with the strongest performance in EUR-like participants, consistent with the predominantly European ancestry of the UKB training cohort. The UKB-trained current alcohol drinking score also generalized across AoU ancestry groups, although performance varied more across populations. Based on the correlation metric, performance was highest in SAS-like participants ($r$ = 0.300, 95% CI: 0.201-0.393; AUC = 0.749, 95% CI: 0.696-0.801), followed by EAS-like ($r$ = 0.279, 95% CI: 0.171-0.380; AUC = 0.830, 95% CI: 0.764-0.896), AFR-like ($r$ = 0.239, 95% CI: 0.141-0.332; AUC = 0.696, 95% CI: 0.639-0.753), AMR-like ($r$ = 0.222, 95% CI: 0.132-0.309; AUC = 0.729, 95% CI: 0.675-0.784), and EUR-like ($r$ = 0.150, 95% CI: 0.075-0.224; AUC = 0.715, 95% CI: 0.648-0.783). Although EAS-like participants had a lower prevalence of current alcohol drinking than some other ancestry groups, they achieved the highest AUC, indicating that ancestry-specific predictive performance was not determined solely by exposure prevalence, which varied across biobanks and ancestries, but also related to genetic predisposition, for example of *ALDH2* alcohol flushing variants (e.g., rs671) that strongly influence physiological and metabolic responses to alcohol in EAS-like populations(*10*). Overall, the UKB-trained protein-exposure scores demonstrated good cross-cohort performance in AoU, supporting their portability beyond the UKB training population while highlighting the importance of evaluating performance across diverse ancestry groups.

To evaluate generalizability within AoU, we tested pooled-trained protein-exposure scores across strata defined by genetic ancestry, race and/or ethnicity, sex at birth, urbanization, income, education and area deprivation (**Fig. 2D, fig. S7 to S8, table S8**). After adjustment for age, sex at birth, collection year, month of sample collection and time of day of sample collection, score-exposure associations were directionally consistent across most groups but varied by exposure. Cardiometabolic scores showed the strongest and most stable performance: BMI had a high overall correlation of $r$ = 0.845 (0.834, 0.856), with ancestry-stratified estimates ranging from $r$ = 0.810 (0.773, 0.841) in SAS-like to $r$ = 0.860 (0.841, 0.877) in EUR-like. Across socioeconomic strata, BMI score performance was modestly higher in lower-SES groups, with correlations of $r$ = 0.850 (0.828, 0.869) in low-income participants (mean BMI = 29.1 kg/m$^2$) and $r$ = 0.848 (0.827, 0.866) in low-education participants (mean BMI = 29.1 kg/m$^2$), compared to $r$ = 0.829 (0.806, 0.850) in high-income participants (mean BMI = 26.9 kg/m$^2$) and $r$ = 0.824 (0.798, 0.847) in high-education participants (mean BMI = 26.5 kg/m$^2$). Deprivation gradients were less consistent, although the highest correlation was observed in the high-deprivation group ($r$ = 0.864 [0.846, 0.881]; mean BMI = 28.5 kg/m$^2$) compared with the low-deprivation group ($r$ = 0.847 [0.828, 0.865]; mean BMI = 27.3 kg/m$^2$).

Environmental scores were also reproducible across biobanks but to a lesser extent, with overall correlations of $r$ = 0.193 (0.153, 0.233) for 30-day $PM_{2.5}$, $r$ = 0.232 (0.195, 0.269) for 365-day $PM_{2.5}$, $r$ = 0.312 (0.277, 0.347) for 30-day temperature and $r$ = 0.321 (0.286, 0.355) for 365-day temperature. Behavioral scores showed greater heterogeneity across groups and social strata. The current-smoking score had a strong overall point-biserial correlation, $r$ = 0.676 (0.652, 0.698), but varied across ancestry groups, from $r$ = 0.328 (0.224, 0.425) in EAS-like to $r$ = 0.721 (0.678, 0.759) in EUR-like, and across socioeconomic strata, with higher performance in low-income ($r$ = 0.696 [0.652, 0.735]) and low-education participants ($r$ = 0.671 [0.627, 0.712]) than in high-income participants ($r$ = 0.383 [0.312, 0.450]) and high-education participants ($r$ = 0.341 [0.265, 0.413]). These differences paralleled exposure burden: mean cigarettes per day were 8.57 in the low-income group versus 3.14 in the high-income group, and 8.18 in the low-education group versus 3.52 in the high-education group. Smoking prevalence also differed markedly, ranging from 46.1% in the low-income group and 46.2% in the low-education group to 25.4% and 21.9% in the corresponding high-income and high-education groups. Together, these patterns suggest that the stronger score performance in lower-SES groups reflects greater exposure contrast rather than differential protein signal — a pattern mirrored in the phenotype itself.

Marijuana use showed more modest associations with their corresponding protein scores overall ($r$ = 0.312 [0.267, 0.356]), with weaker estimates in groups with fewer users. Consistent with the smoking results, marijuana score performance was higher in low-income ($r$ = 0.338 [0.249, 0.420]; marijuana use prevalence =

21.1%) and low-education participants (*r* = 0.333 [0.255, 0.407]; prevalence = 18.2%) than in high-income (*r* = 0.164 [0.068, 0.257]; prevalence = 10.4%) and high-education participants (*r* = 0.137 [0.039, 0.232]; prevalence = 7.2%). These patterns indicate that protein-exposure scores can generalize broadly while apparent differences in performance can reflect social patterning of exposure prevalence, intensity, and phenotypic variance rather than portability alone. For current smoking, the lower EAS-like *r* estimate could suggest reduced portability, but EAS-like participants also had substantially lower smoking prevalence (15.2%) and lower mean cigarettes per day (3.1) than EUR-like participants (45.2% prevalence; 6.8 cigarettes/day).

Third, we constructed multivariate protein scores to assess exposure specificity (**Fig. 1, Fig. 2E-G, table S9 to S11, fig. S9 to S10**). For each exposure-trained score, we compared prediction of the matched exposure with prediction of all other exposures. Scores were classified as "on-target" when the matched exposure had the highest $R^2$, as "off-target: within-class" when the strongest off-diagonal prediction exceeded the matched exposure but remained within the same exposure class, and as "off-target: between-class" when the strongest off-diagonal prediction was from a different exposure class (Methods). Overall, trait specificity was common across several exposure domains: all anthropometry scores were on-target (3/3, 100%), as were all blood pressure (2/2, 100%), lipid (4/4, 100%), liver (6/6, 100%), and kidney scores (2/2, 100%). Most ambient temperature and air pollution scores were also on-target (3/4, 75% for each), and 65% of hematology scores were on-target (11/17), with an additional 24% showing within-class (4/17). These patterns indicate that aggregated protein scores often preserved exposure-relevant information despite widespread cross-exposure associations at the individual protein level.

In contrast, SES and Tobacco/Substance scores showed weaker single-trait specificity. Among the four SES scores, only deprivation index was on-target (matched $R^2$=0.100 [0.079, 0.122]); income, education, and urbanization showed between-class transfer. Income and education scores most strongly cross-predicted smoking or alcohol-related traits, with the income score predicting current cigarette smoking most strongly off-target ($R^2$=0.134 [0.161, 0.109]) and the education score predicting frequent alcohol use ($R^2$=0.077 [0.043, 0.120]). The urbanization score most strongly cross-predicted BMI ($R^2$=0.133 [0.157, 0.110]), exceeding its matched-exposure $R^2$ of 0.056 (0.040, 0.074). Tobacco/Substance scores were highly heterogeneous: only 2 of 34 were on-target (5.9%), 14 of 34 showed within-class (41.2%), and 18 of 34 showed between-class transfer (52.9%). The on-target Tobacco/Substance scores were current cigarette smoking (matched $R^2$=0.490 [0.460, 0.520]) and frequent alcohol use (matched $R^2$=0.352 [0.294, 0.410]). Among within-class scores, several cross-predicted current cigarette smoking, including cigarette-per-day ($R^2$=0.431 [0.400, 0.462]), ever smoking ($R^2$=0.408 [0.377, 0.439]), marijuana ever use ($R^2$=0.200 [0.171, 0.231]), current marijuana use ($R^2$=0.195 [0.166, 0.225]), and marijuana plus cigarette smoking ($R^2$=0.257 [0.226, 0.289]). Other Tobacco/Substance scores showed between-class transfer, with strongest off-diagonal predictions involving anthropometric, kidney, glycemic, or hematologic traits, including height, waist-to-hip ratio (WHR), estimated Glomerular Filtration Rate (eGFR), creatinine, fasting glucose, insulin, mean Corpuscular Hemoglobin (MCH) White Blood Cell count (WBC). For example, ever cigar smoking most strongly predicted height ($R^2$=0.323 [0.295, 0.351]), smokeless tobacco most strongly predicted height ($R^2$=0.319 [0.291, 0.347]), hookah and electronic smoking most strongly predicted eGFR ($R^2$=0.217 [0.162, 0.276] and $R^2$=0.218 [0.163, 0.277], respectively), and cigarette smoking only most strongly predicted fasting glucose ($R^2$=0.221 [0.095, 0.368]). These results suggest a hierarchy of specificity: many anthropometric and biomarker scores behaved as precise exposure-specific signatures, whereas SES and Tobacco/Substance scores more often captured broader behavioral, demographic, or systemic proteomic patterns. Indeed, after accounting for phenotypic correlations among exposures, the cigarette-per-day score predicted its matched cigarette-per-day trait more strongly than any residual off-target association, supporting the interpretation that cross-prediction among smoking-related traits reflects shared behavioral structure rather than arbitrary nonspecificity (**table S11, fig. S11**).

#### Proteomic validation of air pollution exposure signatures

To evaluate whether the PM2.5 protein score captured measured air pollution exposure, and to characterize the broader environmental and biological correlates of this score, we first examined its spatial distribution and marginal associations with exposures. Because residential location was available only at the ZIP3 level, annual average PM2.5 estimates provided at the census-tract level were summarized to ZIP3 regions using population-density weighting and linked to individuals by ZIP3. For spatial validation, we independently classified each ZIP3 into tertiles (low, middle, high) of (i) observed ZIP3-level annual average PM2.5 and (ii) ZIP3-level mean PM2.5 protein score, then compared the two tertile classifications across ZIP3s (**Fig. 3A**). Among 247 ZIP3s with complete data, 46.6% showed concordant tertile assignment between observed PM2.5 and the protein score, comprising 18.6% low observed PM2.5/low protein score, 11.3% middle observed PM2.5/middle protein score, and 16.6% high observed PM2.5/high protein score regions. Among states represented by at least 10 ZIP3s, California showed the highest overall concordance (68.9%), driven largely by high observed PM2.5/high protein score regions. However, the bivariate map also revealed local heterogeneity and discordance: 5.7% of ZIP3s had high observed PM2.5 but a low protein score, while 4.0% had low observed PM2.5 but a high protein score. These discordant patterns may reflect residual geographic or socioeconomic confounding, ZIP3-level exposure misclassification, temporal mismatch between exposure and protein measurement, individual-level exposure heterogeneity, or heterogeneity in biological response.

The PM2.5 protein score was most strongly correlated with measured annual PM2.5 exposure (**Fig. 3B**), with $R^2 = 0.064$. The score also correlated with urbanization, although more modestly ($R^2 = 0.015$), supporting urbanization as a relevant but incomplete proxy for PM2.5-related environmental structure. Beyond air pollution and urbanization, the PM2.5 protein score was associated with several biomarker and socioeconomic correlates, including albumin, eGFR, hemoglobin, lymphocyte count, BMI, neutrophil count, white blood cell count, CRP, smoking-related variables, education, HDL, triglycerides, and average cold temperature. These associations indicate that the PM2.5 protein score captures a broader environmental and biological axis that includes air pollution, urbanicity, socioeconomic patterning, and inflammatory, metabolic, hematologic, renal, and hepatic markers.

We next evaluated whether protein scores trained on PM2.5 and related environmental or geographic traits explained measured $PM_{2.5}$ in the held-out test set (**Fig. 3C, table S12**). In models adjusted for age, sex, and blood collection timing, the $PM_{2.5}$ protein score explained the largest fraction of $PM_{2.5}$ variance (incremental partial $R^2 = 0.051$, 95% CI: 0.035, 0.069). The urbanization protein score also predicted $PM_{2.5}$ (partial $R^2 = 0.020$, 95% CI: 0.011, 0.032), whereas temperature, ozone, longitude, and latitude scores each explained comparatively little variance. A joint model including all six protein scores explained $R^2 = 0.053$ of $PM_{2.5}$ variance (95% CI: 0.040, 0.074), only modestly greater than the $PM_{2.5}$ score alone, indicating that most $PM_{2.5}$-predictive proteomic information was captured by the $PM_{2.5}$-trained score, with limited additional independent information from related environmental and geographic protein scores. Correlation analysis among protein scores showed moderate overlap between the $PM_{2.5}$ and urbanization scores ($r = 0.52$), consistent with shared proteomic variation related to urban environmental structure (**Fig. 3D**). The $PM_{2.5}$ score was also modestly inversely correlated with latitude ($r = -0.35$) and longitude scores ($r = -0.20$), suggesting some geographic patterning. In contrast, temperature, ozone, latitude, and longitude scores were strongly intercorrelated, including temperature with longitude ($r = -0.87$) and ozone with latitude ($r = -0.77$), indicating substantial shared spatial-environmental structure. These correlations may explain why the joint model including all six protein scores only modestly improved $PM_{2.5}$ prediction over the $PM_{2.5}$ protein score alone.

UKB-derived air-pollution protein-exposure scores showed limited transferability to AoU overall, with the strongest validation signal observed for $NO_2$ in AMR-like participants (**fig. S5, table S6**). In the full AoU analysis, the covariate-adjusted incremental $R^2$ for the UKB $NO_2$ score was highest in AMR-like participants ($R^2$ = 0.050, 95% CI 0.033-0.071), compared with EUR-like (0.016, 0.008-0.027), AFR-like (0.002, 0.000-0.009), EAS-like (0.003, 0.000-0.012), and SAS-like (0.006, 0.001-0.016). This stronger performance in AMR-like may partly reflect greater exposure variation, as AMR-like participants had the highest mean $NO_2$ level and large variance in the full AoU set (10.20 [4.36]) compared with EUR-like (7.64 [4.01]), AFR-like (8.85 [4.45]), EAS-like (9.77 [4.10]), and SAS-like (8.89 [4.06]). Despite being developed primarily in UKB participants of EUR-like ancestry, the $NO_2$ score therefore generalized best to the AoU population with the broadest observed $NO_2$ distribution. Overall, however, validation of UKB air-pollution scores was modest. Furthermore, additional adjustment for geographic region substantially attenuated the observed associations (**fig. S12**), with incremental $R^2$ values decreasing from 0.050 (0.033-0.071) to 1.53E-02 (7.53E-03, 0.026681755) for $NO_2$ and from 0.010 (0.003, 0.021) to 2.60E-03 (3.67E-04, 0.007) for $PM_{10}$ in AMR-like and 0.033 (0.018, 0.054) to 9.29E-03 (2.67E-03, 0.020) for $PM_{2.5}$ in AFR-like. This suggests that part of the predictive signal captured by these scores may reflect geographic environmental structure rather than individual-level exposure variation alone, consistent with what we found in our within-AoU validation above.

This limited portability is likely related to substantial differences in exposure definitions across cohorts. The UKB scores were trained using UKB-derived air-pollution phenotypes defined for specific annual exposure surfaces, including HEAP/UKB-derived mean $NO_2$ and $PM_{2.5}$ scores and UKB Field 24005 $PM_{10}$ annual average air pollution in 2010. In contrast, the AoU validation phenotypes were assigned based on U.S. residential ZIP3 and biospecimen collection timing. $PM_{2.5}$ exposure in AoU was defined as the average CDC Downscaler daily $PM_{2.5}$ concentration over the 365 days preceding biospecimen collection, whereas $NO_2$ and $PM_{10}$ were based on CACES annual average concentrations assigned according to each participant's collection year (Methods). Thus, the AoU phenotypes incorporate collection-date-aligned or collection-year-aligned exposure windows, while the UKB phenotypes reflect fixed or UKB-derived annual exposure estimates. These differences in geography, exposure models, temporal windows, and spatial aggregation likely contributed to the weaker cross-cohort validation of air-pollution protein-exposure scores compared with the smoking and alcohol scores.

#### Proteomic exposure signatures reflect socially patterned metabolic context

We stratified exposure-protein associations analyses across genetic ancestry and self-reported race and/or ethnicity groups to evaluate heterogeneity. Among all exposures examined, BMI showed the greatest degree of heterogeneity (**fig. S3**), with 111 (2.05%) and 116 (2.14%) proteins having significant meta-analysis and Cochran Q test across ancestry and race and/or ethnicity groups, respectively. Of those proteins associated with BMI, leptin was most heterogeneous (Cochran Q test p-value [$p_{het}$]=1.38e-19 for ancestry and $p_{het}$=1.97e-11 for race/ethnicity). BMI itself is highly heterogeneous across AoU ancestry and race and/or ethnicity groups: lowest mean and standard deviation in EAS-like (25.3 kg/m² [4.5]) and Asian (race and/or ethnicity) (25.6 kg/m² [4.6]), highest mean and variance in AFR-like (29.5 kg/m² [6.8]) and Black (29.5 kg/m² [7.0]) (**Fig. 4A**, **fig. S13A, fig. S16, table S13**). Consequently, the magnitude and ordering of BMI-protein associations varied substantially depending on the transformation applied to BMI, including raw values, z scores, and inverse rank normal transformation (IRNT). Using raw BMI or z scores, the association between BMI and leptin (LEP) was strongest in EAS-like populations, followed by AFR-like, EUR-like, AMR-like, and SAS-like groups. A similar pattern was observed across self-identified race and/or ethnicity categories, with the largest effect sizes in Asian individuals, followed by Black, White, Hispanic, and Mixed populations. Given the smaller variance of BMI in EAS-like populations, regression coefficients can be larger with these transformations. We therefore also applied IRNT, which standardizes the variance of BMI across groups. Under these approaches, the pattern of association shifted, with the strongest effects observed in AFR-like populations, followed by EUR-like, AMR-like, EAS-like, and SAS-like groups. Comparable trends were observed across race and/or ethnicity categories, with the largest effects in Black individuals, followed by White, Hispanic, Mixed, and Asian populations. These findings indicate that differences in phenotype scaling can substantially influence cross-population comparisons of effect sizes and imply that cautious interpretation in relative effect directions is warranted.

BMI was positively associated with leptin in all ancestry groups, but with substantial heterogeneity in effect sizes ($p_{het} = 1.38 \times 10^{-19}$), with the strongest associations observed in AFR-like and EUR-like individuals and smaller effects in EAS-like and SAS-like populations (**fig. S13B**). WHR-leptin associations also showed heterogeneity ($p_{het} = 4.16 \times 10^{-5}$), although to a lesser extent. The cross-ancestry heterogeneity is more pronounced in females than males for both BMI ($p_{het} = 8.48 \times 10^{-17}$ for female and 5.18e-03 for male) and WHR ($p_{het} = 1.54 \times 10^{-5}$ for female and $2.07 \times 10^{-3}$ for male). Comparable trends are observed across race and/or ethnicity groups for BMI ($p_{het} = 2.62 \times 10^{-7}$ for female and $8.54 \times 10^{-4}$ for male) and ($p_{het} = 9.46 \times 10^{-5}$ for female and $7.45 \times 10^{-3}$ for male) (**fig. S13B**).

Predicted leptin levels across BMI revealed clear nonlinear and ancestry-specific patterns (**fig. S16A**). At lower BMI levels (25 kg/m²), predicted leptin was highest in SAS-like among both males (-0.38) and females (0.51), and lowest in AFR-like males (-0.88) and EAS-like females (0.12). In contrast, at higher BMI levels (35 kg/m²), EAS-like individuals had the lowest predicted leptin in both males (0.14) and females (0.77), whereas SAS-like males had the highest predicted leptin (0.53) and other female ancestry groups showed higher values than EAS-like (1.07 to 1.15). These patterns suggest that ancestry-specific differences in predicted leptin varied nonlinearly across the BMI distribution, with EAS-like individuals exhibiting relatively lower predicted leptin at higher BMI in both sexes. To formally assess this nonlinearity, we compared models with shared versus ancestry-specific nonlinear BMI effects. Allowing ancestry-specific nonlinear terms significantly improved model fit ($P = 4.96 \times 10^{-18}$), and results were robust when restricted to overlapping BMI ranges across ancestries ($P = 3.94 \times 10^{-11}$). In contrast, predicted leptin levels across WHR were more consistent across ancestries, with largely parallel curves and minimal divergence.

#### Cross-ancestry differences in BMI-associated T2D risk

We next examined the relationship between BMI and T2D risk across ancestry and sex groups (**fig. S16B**). Predicted T2D risk increased monotonically with BMI in all populations, but the magnitude and trajectory of risk differed across ancestries, consistent with previous reports(*11–14*). For example, at a BMI of 30 kg/m$^2$ (a conventional threshold for obesity), the predicted T2D risk in the EUR-like ancestry group was 14.1% in male. However, the BMI required to reach an equivalent risk varied across populations: AFR-like, 27.9 kg/m$^2$; AMR-like, 23.9 kg/m$^2$; EAS-like, 27.5 kg/m$^2$; and SAS-like, 22.7 kg/m$^2$. Among females, a BMI of 30 kg/m² in the EUR-like group corresponded to a predicted T2D risk of 8.2%. The equivalent BMI was 21.4 kg/m² in AFR-like, 22.4 kg/m² in AMR-like, 25.0 kg/m² in EAS-like, and 22.2 kg/m² in SAS-like individuals. Overall, EAS-like and SAS-like females reached the same predicted T2D risk at substantially lower BMI than their EUR-like counterparts, with a greater downward shift in BMI thresholds than observed in males.

#### Leptin does not independently explain T2D risk beyond BMI

Finally, we assessed whether leptin contributes to T2D risk independently of BMI (**fig. S16C**). In models unadjusted for BMI, leptin was positively associated with T2D risk across all ancestry groups in males (AFR-like: $\beta = 0.442$, SE = 0.116; AMR-like: $\beta = 0.392$, SE = 0.115; EAS-like: $\beta = 0.520$, SE = 0.203; EUR-like: $\beta = 0.542$, SE = 0.091; SAS-like: $\beta = 0.396$, SE = 0.170; all $P < 0.05$), but was significant only in EUR-like females ($\beta = 0.524$, SE = 0.100, $P = 1.51 \times 10^{-7}$). After adjustment for BMI, these associations in males were substantially attenuated toward the null across all ancestry groups (AFR-like: β = 0.128, SE = 0.169; AMR-like: β = 0.063, SE = 0.154; EAS-like: β = 0.041, SE = 0.261; EUR-like: β = 0.157, SE = 0.122; SAS-like: β = 0.095, SE = 0.225; all P > 0.05). In contrast, in females, BMI adjustment revealed an inverse association between leptin and T2D risk in several ancestry groups (AFR-like: β = -0.297, SE = 0.147, P = 0.044; AMR-like: β = -0.766, SE = 0.161, P = $2.08 \times 10^{-6}$; EUR-like: β = -0.335, SE = 0.150, P = 0.026), while the association remained non-significant in EAS-like (β = -0.449, SE = 0.290, P = 0.122) and SAS-like (β = -0.219, SE = 0.240, P = 0.361). Although the point estimate in the AMR-like group was already directionally below the null in the unadjusted model (β = -0.174, SE = 0.123, P = 0.155) and remained the most strongly inverse association after adjustment, there was no significant heterogeneity across ancestries ($P_{het} \approx 0.40$), indicating that this observation is likely due to sampling variability rather than a true ancestry-specific effect. These findings indicate that the association between leptin and T2D is largely explained by overall adiposity captured by BMI.

#### BMI-matched leptin/T2D analysis

To assess whether this heterogeneity simply reflected differing BMI distributions across race and/or ethnicity groups, we repeated the analysis after matching each group's BMI distribution to that of Asian (race and/or ethnicity) participants (the reference group) (**fig. S14**). This matching did not produce a consistent narrowing of the leptin-BMI association: the gap relative to Asian participants narrowed for Black (+0.097 to +0.070) and Hispanic (+0.052 to +0.005) participants, but widened for White participants (+0.121 to +0.158) and reversed sign for Mixed participants (+0.013 to −0.004; n=169 after matching, a 46% reduction from the original sample). This inconsistent pattern indicates that race and/or ethnicity differences in the leptin-BMI relationship are not primarily attributable to differing BMI distributions across groups, unlike the pattern observed for T2D prevalence. Baseline T2D prevalence differed by race and/or ethnicity (range 11.7% in Asian to 18.0% in Hispanic participants); matching BMI distributions across groups substantially narrowed this gap for White, Black, and Hispanic participants (e.g., Hispanic: gap vs. Asian narrowed from +6.3 to +1.7 percentage points; White: +5.2 to +1.6; Black: +5.0 to +1.4), consistent with BMI as a substantial partial mediator of racial and/ or ethnicity disparities in T2D prevalence.

### Epidemiology of substance use in AoU

Cigarette smoking and marijuana use were highly prevalent in the All of Us proteomic cohort, with substantial heterogeneity across demographic and socioeconomic strata (**figs. S17 to S21, table S16 to S17**). Overall, 47.3% (3,979/8,418) of participants reported ever using marijuana and 34.8% (2,926/8,418) reported ever smoking cigarettes, with substantial co-use between exposures. These estimates were similar to national lifetime-use estimates from major U.S. surveillance systems, including lifetime marijuana use in the 2023 National Survey on Drug Use and Health (NSDUH) (47.1% in 12 years or older community-dwelling United States population)(*15*) and ever cigarette smoking in the 2023 National Health Interview Survey (NHIS) (33.3%, defined as having smoked ≥100 cigarettes in one's lifetime)(*16*). Among cigarette smokers in the All of Us proteomic cohort, 70.9% (2,074/2,926) also reported marijuana use, whereas 47.9% (1,905/3,979) of marijuana users reported smoking cigarettes (**Fig. 5A**, **fig. S22**).

Age- and sex-stratified analyses demonstrated distinct substance use exposure patterns (**fig. S17A**). Marijuana use was more prevalent in younger individuals and declined with age, with the highest prevalence observed among individuals aged 18-34 years for both males (58%) and females (51%). In contrast, cigarette smoking was more common in older participants, peaking among males aged 50-64 years (49%) and females aged 65 years or older (33%). Across nearly all strata, both exposures were more prevalent in males than females. Stratification by genetic ancestry showed that AFR-like and EUR-like groups had the highest overall prevalence of both marijuana use and cigarette smoking, with distinct sex-specific patterns. Among AFR-like participants, marijuana-use prevalence was 65% in males and 44% in females, while cigarette-smoking prevalence was 61% in males and 35% in females. AMR-like participants showed a similar degree of sex stratification, although overall prevalence was lower (46% in males vs. 29% in females for marijuana use). In contrast, EUR-like participants showed more even sex distributions for marijuana use, with prevalence of 59% in both males and females, but retained a male excess for cigarette smoking, with prevalence of 45% in males and 40% in females (**fig. S18**). Other ancestry groups had more limited sample sizes, making their sex-specific patterns more difficult to interpret reliably.

Socioeconomic gradients were also evident (**fig. S20**, **table S16**). Cigarette smoking was more prevalent among participants with lower income (lowest vs. highest income group - Male: 64% vs. 30%; Female: 50% vs. 22% at 50-64 years), lower educational attainment (lowest vs. highest education group - Male: 67% vs. 22%; Female: 42% vs. 18% at 50-64 years), and higher neighborhood deprivation (highest vs. lowest deprivation group - Male: 55% vs. 40%; Female: 36% vs. 28% at 50-64 years), consistent with prior epidemiologic evidence that cigarette smoking remains strongly socially patterned in the United States(*17*). Marijuana use showed more heterogeneous socioeconomic patterning. Among males, marijuana-use prevalence was higher in the high-deprivation group (53%) than in the mid- and low-deprivation groups (49% in each). Among females, the pattern was less monotonic, with prevalence of 38% in the high-deprivation group, 36% in the mid-deprivation group, and 45% in the low-deprivation group. Substance-use intensity also varied by socioeconomic strata, with higher prevalence of current smoking and higher mean cigarettes per day among participants with lower income, lower educational attainment, and higher neighborhood deprivation.

Geographic variation further highlighted heterogeneity in exposure patterns across U.S. regions (**fig. S21**, **table S17**). Among states with at least 100 participants with available substance use data, marijuana use prevalence was elevated in several Midwestern and Northeastern states, including Illinois (IL; 58.1%), Michigan (MI; 53.6%), Massachusetts (MA; 52.8%), and Pennsylvania (PA; 51.4%), with additional high estimates in Arizona (AZ; 55.7%) in the West and Louisiana (LA; 67.2%) in the South. Cigarette smoking prevalence was highest in Southern states, particularly Louisiana (LA; 60.0%) and Alabama (AL; 46.2%), and was also elevated in Arizona (AZ; 45.0%) in the West, Illinois (IL; 42.8%) in the Midwest, and Wisconsin (WI; 38.8%) in the Midwest. Other substance use behaviors, including e-cigarette, hookah, and smokeless tobacco use, exhibited distinct but partially overlapping geographic patterns, consistent with regional differences in social norms and policy environments(*18*, *19*).

#### Shared and distinct proteomic associations of marijuana use and cigarette smoking

At the individual protein level, marijuana use and cigarette smoking showed substantial overlap, with 190 proteins reaching Bonferroni significance for both exposures (**Fig. 5B, table S18**). Several of these shared proteins have prior biological and genetic support linking them to smoking-related or neurobehavioral processes. For example, NCAM1 was strongly associated with both cigarette smoking ($\beta$ = -0.225, SE = 0.022, $P_{bonf}$ = 7.29 × $10^{-21}$) and marijuana use ($\beta$ = -0.098, SE = 0.02, $P_{bonf}$ = 0.019); rs7935745 in *NCAM1* is a well-replicated locus in GWAS of smoking initiation (*20*), nicotine dependence(*21*), and smoking motives(*22*). NCAM1 knockout models have implicated it in synaptic plasticity(*23*–*25*). The shared proteins also include lung-enriched secretory proteins, including SCGB1A1, SCGB3A1/SCGB3A2, and SFTPA2 (**table S4**), which have well-characterized roles in airway epithelial function and are consistently altered in response to cigarette smoke exposure(*26*, *27*). Consist with this, the shared proteins are most enriched in the lung tissue ($-\log_{10}$FDR = 7.14, OR = 6.3) (**Fig. 5C, table S19**), an enrichment that held consistently across trait definitions (ever use, current use, cigarette-adjusted marijuana use, and marijuana-only for marijuana; and ever smoking, current smoking, cigarettes per day, and smoking-only for cigarette smoking) (**fig. S23 to S24**), supporting a dominant, shared downstream physiological response to inhalational exposures. The shared proteins also demonstrated enrichment in liver (Liver_Mixed_Cell: $-\log_{10}$FDR = 5.39, OR = 5.37), gastrointestinal (Stomach: $-\log_{10}$FDR = 3.54 , OR = 7.95), and central nervous system (CNS) regions (Brain_Frontal_Cortex_BA9: $-\log_{10}$FDR = 3.60, OR = 2.89). This component includes metabolic and neuroendocrine regulators such as GHRL, POMC, IGFBP1, and IGFBP2 (**table S4**), suggesting a neuroendocrine axis linking central appetite regulation with peripheral metabolic signaling(*28*, *29*), consistent with prior experimental evidence (*30*–*32*). Nevertheless, this signal may reflect both downstream exposure effects and upstream or shared biology linking body composition with substance-use behavior, motivating further analyses of smoking, marijuana use, BMI, and disease outcomes.

Several proteins associated with both traits, such as NOS3 (endothelial nitric oxide synthase, eNOS), showed opposite directions of effect (**fig. S25**). For example, NOS3 (endothelial nitric oxide synthase, eNOS) demonstrated opposing associations across exposures. NOS3 levels were inversely associated with marijuana use ($\beta$ = -0.126, SE = 0.022, $P_{bonf}$ = $1.10\times10^{-4}$), with the effect strengthening after adjustment for cigarette smoking ($\beta$ = -0.169, SE = 0.024, $P_{bonf}$ = $1.52\times10^{-7}$), indicating an independent marijuana-related signal. In contrast, NOS3 was positively associated with current cigarette smoking ($\beta$ = 0.164, SE = 0.030, $P_{bonf}$ = $5.20\times10^{-4}$), but not with ever smoking ($\beta$ = 0.077, SE = 0.023, $P_{bonf}$ = 1.00). These bidirectional associations are consistent with extensive experimental evidence that cigarette smoke disrupts endothelial function and nitric oxide signaling, including altered eNOS expression and activity and reduced nitric oxide bioavailability(*33*–*35*), whereas cannabinoid effects on endothelial nitric oxide pathways are more heterogeneous and context-dependent(*36*, *37*).

Proteins significant only in cigarette smoking (n=495) were enriched for pathways related to lung injury, extracellular matrix remodeling, and inflammation, including matrix metalloproteinases (e.g., MMP1, MMP8, MMP9, MMP12), vascular and endothelial markers (e.g., ICAM1, VEGFA), and surfactant-related proteins (e.g., SFTPD), all of which are strongly implicated in smoking-related pulmonary pathology(*38*–*40*). Similarly, CHI3L1 (YKL-40) (CHI3L1: $\beta$ = 0.203, SE = 0.020, $P_{bonf}$ = $5.93 \times 10^{-20}$ with cigarette smoking) is a well-established biomarker of inflammation and tissue remodeling, elevated in smokers and associated with chronic lung disease(*41*, *42*). Indeed these cigarette smoking proteins are highly enriched in the lung ($-\log_{10}$FDR = 12.65, OR = 5.20) (**Fig. 5C**) and the pattern was observed consistently across trait comparisons (ever use, current use, cigarette-adjusted marijuana use, and ever smoking, current smoking, cigarettes per day) (**fig. S23 to S24**).

Fewer proteins significantly associated with marijuana use and not smoking (n=187), and they showed weaker support from existing human genetic studies, consistent with the comparatively lower power and heterogeneity of current cannabis GWAS(*43*, *44*). These proteins are most enriched in liver ($-\log_{10}$FDR = 9.39, OR = 6.95). But when comparing Marijuana Use (Current) vs. Cigarette smoking (Ever), different enrichment patterns are found (**Fig. 5C,E, table S19**). 54 proteins are uniquely associated with marijuana use, showing significant enrichment in liver ($-\log_{10}$FDR = 9.39, OR = 6.95), Connective/bone (Adipose_Subcutaneous: $-\log_{10}$FDR = 4.01, OR = 5.63), and Immune/hematopoietic (Spleen: $-\log_{10}$FDR = 3.27, OR = 2.87). Among these, 5 proteins (CA14, DNAJC6, GPIHBP1, MEGF10, SEMA7A) showed significant enrichment in Brain_Spinal_cord_cervical ($-\log_{10}$FDR = 1.70, OR = 6.22) and 3 proteins (CA14 DNAJC6 MEGF10) showed significant enrichment in Brain_Substantia_nigra ($-\log_{10}$FDR = 1.22, OR = 8.15). Notably, CA14 has been implicated in schizophrenia previouslyin a 2025 cross-ancestry GWAS meta-analysis(*45*), where it was prioritized as a causal gene candidate via multi-omic analyses. Schizophrenia has well-known genetic overlap with cannabis use disorder (shared loci including GRM3, dopamine pathways), but CA14 itself has not been linked to substance use. Biologically, CA14 is a membrane-bound carbonic anhydrase involved in CNS pH regulation and neuronal excitability, suggesting a plausible but unproven connection to addiction neuroscience. NOS1 (neuronal nitric oxide synthase, nNOS) showed a pattern specific to marijuana use. NOS1 was positively associated with marijuana use ($\beta = 0.115$, SE = 0.019, $p_{bonf} = 1.55\times10^{-5}$), with a stronger effect after adjustment for cigarette smoking ($\beta = 0.118$, SE = 0.021, $p_{bonf} = 2.00\times10^{-4}$), and showed no association with either ever ($\beta = 0.060$, SE = 0.020, $p_{bonf} = 1.00$) or current cigarette smoking ($\beta = 0.039$, SE = 0.027, $p_{bonf} = 1.00$). Neuronal nitric oxide signaling is directly linked to cannabinoid biology. CB1 receptor activation modulates nNOS-dependent nitric oxide production and downstream processes including neurogenesis, synaptic plasticity, and reward-related signaling(*46*, *47*).

Because substance-use prevalence differed across socioeconomic strata, we performed SES-stratified analyses by income, deprivation, and education tertiles (**fig. S26 to S28**, **table S20 to S22**). The SES-stratified results showed more extensive overlap between marijuana- and cigarette-smoking-associated proteins in lower SES groups: 46.8% of dual users were in the low-income group, 37.5% were in the high-deprivation group, and 38.8% were in the low-education group. The high-deprivation group showed the largest and most significant lung-enriched shared protein set across deprivation strata (n = 71, $-\log_{10}$FDR = 14.29, fold change [FC] = 28.50), compared with mid deprivation (n = 35, $-\log_{10}$FDR = 9.81, FC = 36.64) and low deprivation (n = 19, $-\log_{10}$FDR = 4.92, FC = 37.50). Similarly, the low-education group showed the strongest lung enrichment across education strata (n = 121, $-\log_{10}$FDR = 19.33, FC = 23.95), compared with mid education (n = 42, $-\log_{10}$FDR = 9.03, FC = 30.53), with no shared protein set detected in the high-education group. The low-income group also showed a larger and more statistically significant lung-enriched shared protein set (n = 87, $-\log_{10}$FDR = 17.61, FC = 28.50) than the mid-income group (n = 11, $-\log_{10}$FDR = 4.29, FC = 51.82), while the high-income group had only one shared protein and no lung enrichment. Importantly, in lower SES strata, lung enrichment for the shared marijuana-smoking protein set was stronger than for the cigarette-smoking protein set alone, including high deprivation (shared: $-\log_{10}$FDR = 14.29, FC = 28.50; smoking: $-\log_{10}$FDR = 6.51, FC = 8.72), low education (shared: $-\log_{10}$FDR = 19.33, FC = 23.95; smoking: $-\log_{10}$FDR = 3.14, FC = 6.56), and low income (shared: $-\log_{10}$FDR = 17.61, FC = 28.50; smoking: $-\log_{10}$FDR = 3.21, FC = 7.12). These findings suggest a larger and more robustly lung-enriched shared marijuana-smoking proteomic signature in lower SES groups, although differences in co-use prevalence, exposure intensity, residual confounding, and statistical power may contribute to this pattern. These findings indicate that lower SES strata have a larger and more lung-enriched shared marijuana-smoking proteomic signature. However, this observed pattern may reflect higher co-use prevalence, differences in cumulative smoking or marijuana exposure, residual lifestyle or environmental confounding, and increased statistical power in lower SES groups.

#### Proteomic and genetic architectures reflect complementary biological layers

To place the proteomic findings in the context of inherited susceptibility, we examined published GWAS of smoking initiation(*20*) and cannabis use(*44*), together with tissue and cell-type enrichment analyses (**fig. S29 to S30**). The strongest shared genetic association was observed at *CADM2* (rs1375564; smoking initiation $P = 3.31 \times 10^{-19}$; lifetime cannabis use $P = 8.77 \times 10^{-14}$), a well-established locus associated with risk-taking behavior, impulsivity, cognitive traits, and multiple substance-use phenotypes(*48–50*) (**table S23**). *NCAM1*, a neural cell adhesion molecule involved in synaptic plasticity and dopaminergic signaling, was also strongly associated with smoking initiation ($P = 1.09 \times 10^{-18}$) and showed suggestive association with lifetime cannabis use ($P = 9.23 \times 10^{-5}$), consistent with previous GWAS(*21*, *43*). Consistent with these loci, cell-type enrichment analyses showed that cannabis-associated variants were predominantly enriched in neuronal cell types despite the substantially smaller GWAS sample size, while smoking initiation showed weaker and more diffuse enrichment (**table S24**). These findings suggest that inherited susceptibility to substance use is primarily mediated through neurobehavioral pathways. In contrast, the dominant proteomic components reflected peripheral physiological responses, including airway epithelial injury, systemic metabolic regulation, and immune signaling. These findings support a model in which genetic variation primarily influences susceptibility to initiate substance use, whereas circulating proteomic signatures capture the integrated physiological consequences of exposure across multiple organ systems.

#### Expression-matched (abundance-confound) analysis

To evaluate whether liver enrichment among marijuana- and smoking-associated proteins reflects generic detectability rather than exposure-specific biology, we tested whether liver tissue-elevated status (HPA-style tissue-specificity classification applied to GTEx v10 expression data) predicted univariate significance status independent of protein abundance (mean raw, pre-transformation NPX). Protein abundance was estimated as the mean raw (pre-IRNT, non-transformed) NPX value across all AoU participants for each protein, using pre-transformation intensity data from the same Olink panel. For each of the four liver-related tissue classifications (Liver, Liver_Hepatocyte, Liver_Mixed_Cell, Liver_Portal_Tract), we fit logistic regression models predicting hit status (Bonferroni-significant for marijuana and/or smoking vs. neither) as a function of tissue-elevated status alone, and as a function of tissue-elevated status plus mean raw NPX, and compared the tissue-elevated coefficient between the two models.

Liver-elevated status remained essentially unchanged in magnitude and significance after adjusting for abundance across all four liver-related tissue classifications (Liver, Liver_Hepatocyte, Liver_Mixed_Cell, Liver_Portal_Tract; ORs ≈ 3.7–5.1, all $P < 10^{-17}$ before and after adjustment), and liver-elevated proteins were, if anything, lower in raw abundance than the rest of the panel in every category tested. This indicates that liver enrichment among behavioral-exposure-associated proteins is not attributable to abundance-related detectability bias. Consistent with this, when we compared mean raw NPX across all 69 GTEx/HPA tissue-elevated categories represented on the panel, the four liver-related categories ranked in the lower half of the distribution (Liver_Portal_Tract: rank 11/69; Liver_Hepatocyte: rank 33/69; Liver_Mixed_Cell: rank 36/69; Liver: rank 48/69), well below broadly-expressed, non-tissue-elevated proteins (rank 1/69) and blood/immune-related tissues such as Whole_Blood (rank 2/69) and Spleen (rank 4/69) (Fig. 1).

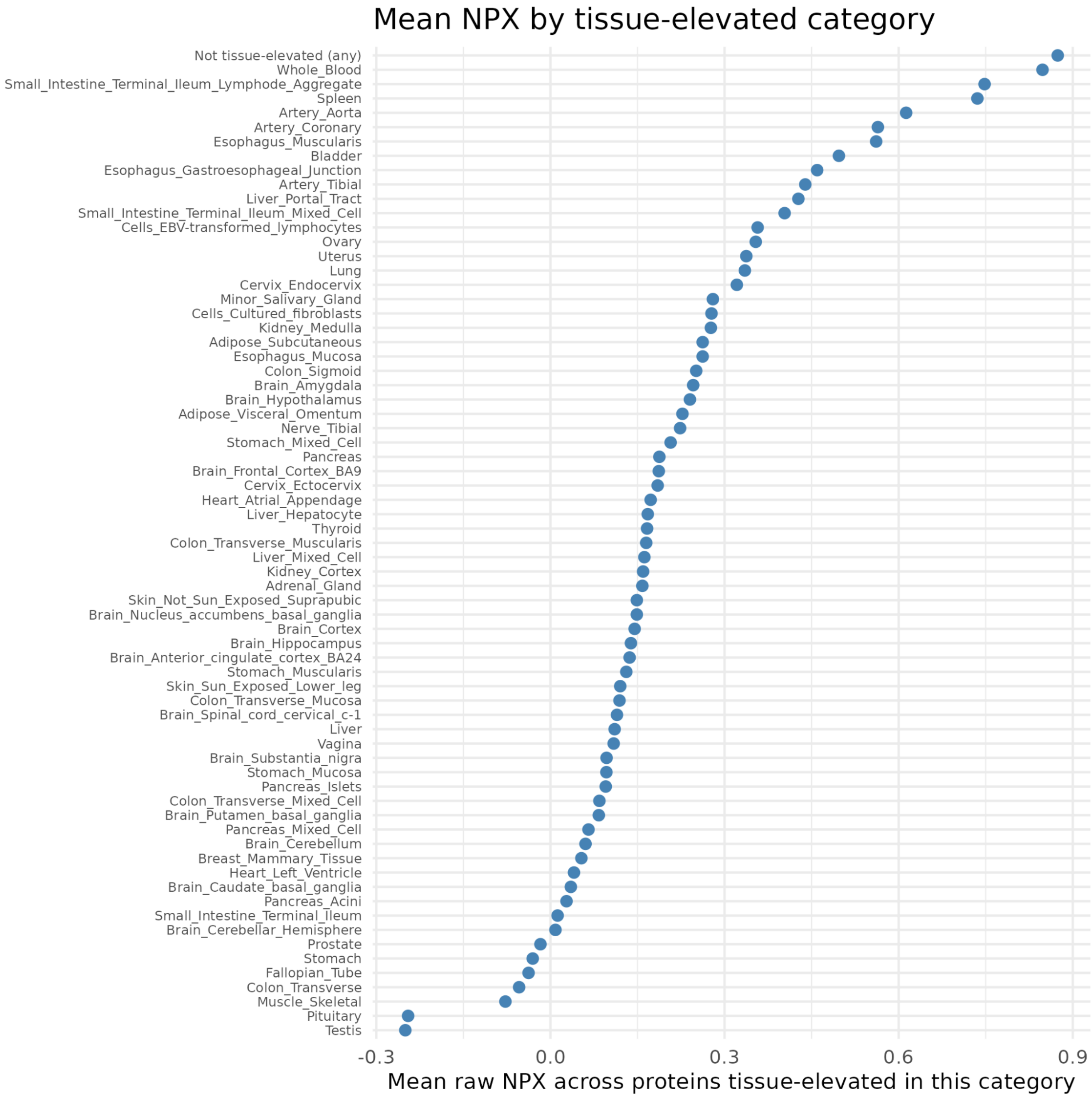


**Fig. 1.** Mean protein abundance across GTEx/HPA tissue-elevated categories in the AoU Olink panel. Each point shows the mean raw (pre-transformation) NPX value, averaged across all proteins classified as tissue-elevated for that tissue category (HPA-style classification applied to GTEx v10 expression data; categories with fewer than 5 tissue-elevated proteins in the panel excluded; 69 categories shown).

20. G. R. B. Saunders, X. Wang, F. Chen, S.-K. Jang, M. Liu, C. Wang, S. Gao, Y. Jiang, C. Khunsriraksakul, J. M. Otto, C. Addison, M. Akiyama, C. M. Albert, F. Aliev, A. Alonso, D. K. Arnett, A. E. Ashley-Koch, A. A. Ashrani, K. C. Barnes, R. G. Barr, T. M. Bartz, D. M. Becker, L. F. Bielak, E. J. Benjamin, J. C. Bis, G. Bjornsdottir, J. Blangero, E. R. Bleecker, J. D. Boardman, E. Boerwinkle, D. I. Boomsma, M. P. Boorgula, D. W. Bowden, J. A. Brody, B. E. Cade, D. I. Chasman, S. Chavan, Y.-D. I. Chen, Z. Chen, I. Cheng, M. H. Cho, H. Choquet, J. W. Cole, M. C. Cornelis, F. Cucca, J. E. Curran, M. de Andrade, D. M. Dick, A. R. Docherty, R. Duggirala, C. B. Eaton, M. A. Ehringer, T. Esko, J. D. Faul, L. Fernandes Silva, E. Fiorillo, M. Fornage, B. I. Freedman, M. E. Gabrielsen, M. E. Garrett, S. A. Gharib, C. Gieger, N. Gillespie, D. C. Glahn, S. D. Gordon, C. C. Gu, D. Gu, D. F. Gudbjartsson, X. Guo, J. Haessler, M. E. Hall, T. Haller, K. M. Harris, J. He, P. Herd, J. K. Hewitt, I. Hickie, B. Hidalgo, J. E. Hokanson, C. Hopfer, J. Hottenga, L. Hou, H. Huang, Y.-J. Hung, D. J. Hunter, K. Hveem, S.-J. Hwang, C.-M. Hwu, W. Iacono, M. R. Irvin, Y. H. Jee, E. O. Johnson, Y. Y. Joo, E. Jorgenson, A. E. Justice, Y. Kamatani, R. C. Kaplan, J. Kaprio, S. L. R. Kardia, M. C. Keller, T. N. Kelly, C. Kooperberg, T. Korhonen, P. Kraft, K. Krauter, J. Kuusisto, M. Laakso, J. Lasky-Su, W.-J. Lee, J. J. Lee, D. Levy, L. Li, K. Li, Y. Li, K. Lin, P. A. Lind, C. Liu, D. M. Lloyd-Jones, S. M. Lutz, J. Ma, R. Mägi, A. Manichaikul, N. G. Martin, R. Mathur, N. Matoba, P. F. McArdle, M. McGue, M. B. McQueen, S. E. Medland, A. Metspalu, D. A. Meyers, I. Y. Millwood, B. D. Mitchell, K. L. Mohlke, M. Moll, M. E. Montasser, A. C. Morrison, A. Mulas, J. B. Nielsen, K. E. North, E. C. Oelsner, Y. Okada, V. Orrù, N. D. Palmer, T. Palviainen, A. Pandit, S. L. Park, U. Peters, A. Peters, P. A. Peyser, T. J. C. Polderman, N. Rafaels, S. Redline, R. M. Reed, A. P. Reiner, J. P. Rice, S. S. Rich, N. E. Richmond, C. Roan, J. I. Rotter, M. N. Rueschman, V. Runarsdottir, N. L. Saccone, D. A. Schwartz, A. H. Shadyab, J. Shi, S. S. Shringarpure, K. Sicinski, A. H. Skogholt, J. A. Smith, N. L. Smith, N. Sotoodehnia, M. C. Stallings, H. Stefansson, K. Stefansson, J. A. Stitzel, X. Sun, M. Syed, R. Tal-Singer, A. E. Taylor, K. D. Taylor, M. J. Telen, K. K. Thai, H. Tiwari, C. Turman, T. Tyrfingsson, T. L. Wall, R. G. Walters, D. R. Weir, S. T. Weiss, W. B. White, J. B. Whitfield, K. L. Wiggins, G. Willemsen, C. J. Willer, B. S. Winsvold, H. Xu, L. R. Yanek, J. Yin, K. L. Young, K. A. Young, B. Yu, W. Zhao, W. Zhou, S. Zöllner, L. Zuccolo, 23andMe Research Team, Biobank Japan Project, C. Batini, A. W. Bergen, L. J. Bierut, S. P. David, S. A. Gagliano Taliun, D. B. Hancock, B. Jiang, M. R. Munafò, T. E. Thorgeirsson, D. J. Liu, S. Vrieze, Genetic diversity fuels gene discovery for tobacco and alcohol use. *Nature* **612**, 720–724 (2022).

38. M.-E. Christopoulou, E. Papakonstantinou, D. Stolz, Matrix metalloproteinases in chronic obstructive

pulmonary disease. *Int. J. Mol. Sci.* **24**, 3786 (2023).

39. H. Takizawa, M. Tanaka, K. Takami, T. Ohtoshi, K. Ito, M. Satoh, Y. Okada, F. Yamasawa, A. Umeda, Increased expression of inflammatory mediators in small-airway epithelium from tobacco smokers. *Am. J. Physiol. Lung Cell. Mol. Physiol.* **278**, L906–13 (2000).

40. C. Dalgård, F. Wang, I. L. Titlestad, K. O. Kyvik, J. Vestbo, G. L. Sorensen, Increased serum SP-D in identification of high-risk smokers at high risk of COPD. *Am. J. Physiol. Lung Cell. Mol. Physiol.* **320**, L1005–L1010 (2021).

41. Y. Sakazaki, T. Hoshino, S. Takei, M. Sawada, H. Oda, S.-I. Takenaka, H. Imaoka, K. Matsunaga, T. Ota, Y. Abe, I. Miki, K. Fujimoto, T. Kawayama, S. Kato, H. Aizawa, Overexpression of chitinase 3-like 1/YKL-40 in lung-specific IL-18-transgenic mice, smokers and COPD. *PLoS One* **6**, e24177 (2011).

42. X. Tong, D. Wang, S. Liu, Y. Ma, Z. Li, P. Tian, H. Fan, The YKL-40 protein is a potential biomarker for COPD: a meta-analysis and systematic review. *Int. J. Chron. Obstruct. Pulmon. Dis.* **13**, 409–418 (2018).

43. J. A. Pasman, K. J. H. Verweij, Z. Gerring, S. Stringer, S. Sanchez-Roige, J. L. Treur, A. Abdellaoui, M. G. Nivard, B. M. L. Baselmans, J.-S. Ong, H. F. Ip, M. D. van der Zee, M. Bartels, F. R. Day, P. Fontanillas, S. L. Elson, 23andMe Research Team, H. de Wit, L. K. Davis, J. MacKillop, Substance Use Disorders Working Group of the Psychiatric Genomics Consortium, International Cannabis Consortium, J. L. Derringer, S. J. T. Branje, C. A. Hartman, A. C. Heath, P. A. C. van Lier, P. A. F. Madden, R. Mägi, W. Meeus, G. W. Montgomery, A. J. Oldehinkel, Z. Pausova, J. A. Ramos-Quiroga, T. Paus, M. Ribases, J. Kaprio, M. P. M. Boks, J. T. Bell, T. D. Spector, J. Gelernter, D. I. Boomsma, N. G. Martin, S. MacGregor, J. R. B. Perry, A. A. Palmer, D. Posthuma, M. R. Munafò, N. A. Gillespie, E. M. Derks, J. M. Vink, GWAS of lifetime cannabis use reveals new risk loci, genetic overlap with psychiatric traits, and a causal influence of schizophrenia. *Nat. Neurosci.* **21**, 1161–1170 (2018).
