## Supplementary material for "Proteomic Signatures Capture Environmental Exposures Across Diverse Populations": Materials and Methods

#### Preprocessing exposure data

##### PM2.5 and ozone

Daily census tract-level concentrations of fine particulate matter ($PM_{2.5}$) and ozone ($O_3$) were obtained from the Centers for Disease Control and Prevention (CDC) Environmental Public Health Tracking Network Downscaler products (https://data.cdc.gov/),. These datasets provide modeled daily pollutant concentrations for all U.S. census tracts by combining measurements from the U.S. Environmental Protection Agency (EPA) Air Quality System monitoring network with outputs from the Community Multiscale Air Quality (CMAQ) model, thereby generating spatially continuous daily estimates.

Daily pollutant concentrations were aggregated from census tracts to ZIP3 regions using population-weighted averages. Let $X_{t,d}$ denote the tract-level concentration for tract *t* on day *d*, and let $w_{t|j}$ represent the proportion of the ZIP3 population residing in tract *t*. The ZIP3-level concentration for ZIP3 region *j* on day *d* was calculated as

$$X_{j,d} = \frac{\sum_{t \in A_j} w_{t|j} X_{t,d}}{\sum_{t \in A_j} w_{t|j}},$$

where $A_j$ denotes the set of census tracts contributing population to ZIP3 *j*.

For each participant, 30-day and 365-day average exposures were calculated by averaging ZIP3-level daily concentrations over the 30-day and 365-day periods preceding the biospecimen collection date.

CDC Downscaler data were available through 2020. To obtain exposure estimates for participants with biospecimen collection dates after 2020, ZIP3-specific daily concentrations for 2021-2023 were extrapolated using generalized additive models fit to historical ZIP3-level pollutant concentrations from 2016-2020. Models included cyclic seasonal effects (day of year), calendar year trends, and ZIP3-specific random effects to preserve both seasonal patterns and spatial variation. Historical observations were retained unchanged, and only post-2020 values were predicted using the fitted models.

##### PM10, CO, NO2, and SO2

Annual average concentrations of particulate matter less than 10 μm in aerodynamic diameter ($PM_{10}$), carbon monoxide (CO), nitrogen dioxide ($NO_2$), and sulfur dioxide ($SO_2$) were obtained from the Center for Air, Climate and Energy Solutions (CACES) national empirical air pollution exposure database. This article includes concentration estimates developed by CACES using v1 empirical models as described in Kim et al. (2018)(S.-Y. Kim et al., 2020) and Lu et al. (2025)(Lu et al., 2025). The CACES database provides annual criteria air pollutant concentration estimates for the contiguous United States based on empirical geospatial prediction models that integrate EPA monitoring data with land-use, emissions, meteorological, satellite, traffic, and other geospatial predictors. We used the CACES LUR-derived population-weighted concentration variable, pred_wght, which represents population-weighted concentrations based on block-level centroid predictions.

Units vary by pollutant. CACES also provides population-weighted latitude and longitude based on block-level centroids.

Annual tract-level CACES predictions were aggregated to ZIP3 regions using the same population-weighting framework described above. For each ZIP3 region $j$ and calendar year $y$, annual pollutant concentrations were calculated as

$$X_{j,y} = \frac{\sum_{t \in A_j} w_{t|j} X_{t,y}}{\sum_{t \in A_j} w_{t|j}},$$

where $X_{j,y}$ denotes the annual average pollutant concentration in tract *t* during year $y$.

CACES annual estimates were available for 2016-2020. For participants whose biospecimen collection occurred during 2021-2022, annual ZIP3-level concentrations were extrapolated using pollutant-specific generalized additive models fit to 2016-2020 ZIP3-level annual estimates. Models included calendar year as a linear predictor and ZIP3-specific random intercepts to preserve spatial heterogeneity while extending temporal trends beyond the observed period. Extrapolated values were used only for collection years beyond the available CACES data period.

Annual pollutant concentrations corresponding to each participant's collection year were assigned based on ZIP3 of residence and collection year. Participants residing in ZIP3 regions outside the geographic coverage of the CACES models (e.g., Alaska, Hawaii, and U.S. territories) were not assigned CACES-based pollutant exposures.

#### Ambient temperature

Daily ambient temperature estimates were obtained from the PRISM Climate Group gridded climate dataset (Oregon State University), which provides daily mean temperature surfaces across the contiguous United States at approximately 4-km spatial resolution. PRISM temperature rasters were accessed through the R package prism and processed for the study period (2016-2024).

Daily raster values were spatially linked to 2010 U.S. Census tracts. For each census tract and day, tract-level mean temperature was estimated by extracting the PRISM temperature value at the tract centroid. Temperatures were converted from degrees Celsius to degrees Fahrenheit.

To evaluate agreement between PRISM-derived estimates and observed temperatures, daily temperature measurements from the EPA Air Quality System (AQS) meteorological monitoring network were assigned to census tracts using a point-in-polygon spatial join. Tract-level observed temperatures were calculated as the arithmetic mean across all monitors within a tract on a given day. Correlation between PRISM-derived and monitor-based tract temperatures was assessed among tract-days with both measurements available.

Daily ZIP3-level temperature estimates were derived by aggregating tract-level PRISM temperatures using population-density weights. Let $w_{t|j}$ denote the proportion of the population of ZIP3 area *j* residing in census tract *t*. For each ZIP3 *j* and day *d*, temperature was calculated as

$$T_{j,d} = \frac{\sum_{t \in A_j} w_{t|j} T_{t,d}}{\sum_{t \in A_j} w_{t|j}}$$

where $T_{t,d}$ is the PRISM-derived daily mean temperature for tract *t* on day *d*, and $A_j$ denotes the set of census tracts contributing population to ZIP3 j. The denominator rescales the weights when tract-level temperature estimates were unavailable for a subset of tracts. The resulting dataset consisted of daily ZIP3-level mean ambient temperature estimates for all ZIP3 regions represented in the study population.

#### Rural-urban classification

Tract-level rural-urban status was defined using the 2010 Rural-Urban Commuting Area (RUCA) codes from the U.S. Department of Agriculture Economic Research Service. RUCA codes classify census tracts into ten categories based on population density, urbanization, and commuting patterns.

For each ZIP3, tract RUCA codes were summarized using population weights to estimate the proportion of residents living in each RUCA category:

$$p_{c,j} = \sum_{t \in j} w_{t|j} 1(C_t = c)$$

where $C_t$ denotes the RUCA category for tract t.

ZIP3s were assigned a dominant RUCA category corresponding to the category with the largest population share. In addition, the urban population proportion (which we use as an “urbanization” variable) for each ZIP3 was calculated as the share of the ZIP3 population residing in tracts classified as metropolitan (RUCA codes ≤ 3).

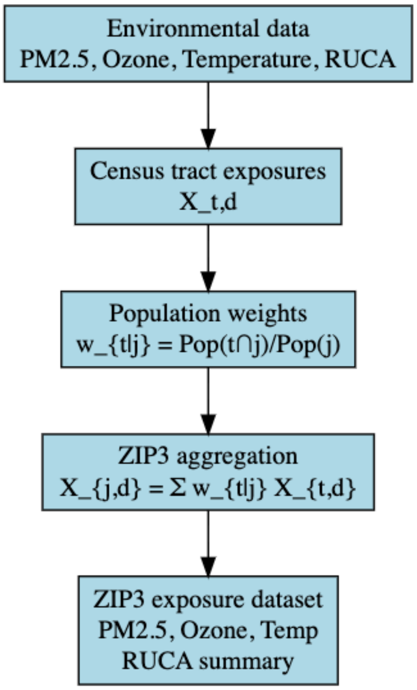


#### Tobacco use

Cigarette smoking status was defined according to established epidemiologic conventions. Participants were first classified based on lifetime cigarette exposure using the question “Have you smoked at least 100 cigarettes in your entire life?". Individuals responding "No" were classified as Never smokers. Among those responding "Yes", current smoking status was determined using the question "Do you now smoke cigarettes every day, some days, or not at all?". Participants reporting "Every day" or "Some days" were classified as Current smokers, whereas those reporting "Not at all" were classified as Previous smokers. Participants with missing or ambiguous responses such as "Don't know" were coded as missing.

For cigars, electronic nicotine products, hookah, and smokeless tobacco, participants were first classified using product-specific "ever use" questions (e.g., "Have you ever used [product], even one or two times?"). Individuals responding "No" were classified as Never users. Among those responding "Yes", current use status was determined using corresponding frequency questions (e.g., "Do you now use [product] every day, some days, or not at all?"). Participants reporting "Every day" or "Some days" were classified as Current users, whereas those reporting "Not at all" were classified as Previous users. Responses such as "Don't know" or missing frequency data were treated as missing.

#### Substance use

Lifetime use of recreational substances was ascertained using the survey question: "In your lifetime, which of the following substances have you ever used?", which allowed multiple selections from a predefined list. Substances evaluated included cocaine, hallucinogens, inhalants, marijuana, methamphetamine, prescription opioids, prescription stimulants, sedatives, street opioids, and other specified drugs.

For each substance, a binary "ever use" indicator was defined. Participants whose response included the substance of interest were classified as ever users, whereas participants who completed the survey and did not select that substance were classified as never users. Participants with missing responses or those who selected "Prefer Not To Answer" without endorsing any substances were coded as missing.

To isolate substance-specific effects independent of co-exposures, mutually exclusive "only-use" phenotypes were constructed. For each substance, participants were classified as only users if they endorsed that substance and did not report use of any other substances or tobacco products, including cigarettes, cigars, electronic nicotine products, hookah, or smokeless tobacco. These participants were coded as 1 for the corresponding "only-use" variable. A shared reference group was defined to enable consistent comparisons across all "only-use" phenotypes. This hyperclean control group comprised participants who reported "None of these drugs" and had no history of use of any tobacco products. Individuals meeting these criteria were coded as controls (0) across all "only-use" variables.

### Smoking and alcohol prevalence-matched analyses

To evaluate whether differences in current-smoking score performance across ancestry and race and/or ethnicity groups reflected differences in smoking prevalence rather than reduced signal portability, we performed a prevalence-matching (downsampling) analysis in both the cross-biobank (UKB-trained score applied to AoU) and within-biobank (AoU pooled-trained score) validation settings. For the UKB-trained current smoking and current drinking, we tested whether ancestry differences in protein score performance reflected between-ancestry differences in exposure prevalence and intensity rather than differences in score validity. For a chosen reference ancestry (EAS-like for smoking, EUR-like for drinking – in each case the ancestry with the lowest covariate-adjusted point-biserial correlation between score and the exposure), we computed quantile bin edges for the continuous intensity variable (cigarettes/day for smoking; a 0-4 ordinal past-year drinking-frequency scale for drinking) using only that reference ancestry's own ever-exposed participants (3 bins for smoking, 4 for drinking; outer edges extended to +/-infinity). Combined with a separate "never-exposed" stratum, this defines a joint categorical distribution over exposure status and intensity. Every other ancestry was then downsampled without replacement, via quota sampling within each stratum, so that its post-matching stratum proportions equal the reference ancestry's; the reference ancestry itself was retained in full. This matches ever/never prevalence and the shape of the intensity distribution among exposed participants simultaneously, in one procedure, rather than matching either dimension alone.

For the AoU-trained protein scores, matching was applied separately to the training (70%) and test (30%) splits, and the LASSO model was retrained from scratch on the matched training set for each analysis; because the reference ancestry's exposed-participant pool in the held-out test set was too small to independently support stable quantile cut points for smoking, bin edges were always computed from the training split and reused, unchanged, to classify test-set participants. Protein score-exposure associations within each ancestry were assessed as partial correlations adjusted for age at biospecimen collection, sex, collection year, and cyclical terms for collection month and time of day.

#### BMI distribution-matched analysis

To evaluate whether cross-race/ethnicity differences in BMI protein score performance and BMI-associated leptin heterogeneity reflected differing BMI distributions across race and/or ethnicity groups rather than differences in the underlying score-phenotype or protein-phenotype relationship, we performed a BMI-distribution-matching analysis using Asian participants — who had the lowest mean BMI among race and/or ethnicity groups — as the reference distribution. BMI values among Asian participants were divided into quintile bins (20th, 40th, 60th, and 80th percentiles), with the lowest and highest bins extended to $-\infty/+\infty$ so that all participants, in every group, were assigned to one of five bins. Participants in each non-Asian race and/or ethnicity group were classified into these same Asian-derived bins, and a common target sample size was identified such that each bin could supply an equal ~20% share of the total matched sample; this target was constrained by the smallest available bin within each group. Within each bin, participants were then randomly subsampled to this target count, yielding a matched subsample per race/ethnicity group with a BMI distribution approximating that of the Asian reference group. The Asian group itself was retained in full (minus participants with missing BMI) and not subsampled. BMI protein score performance, BMI-leptin associations, and T2D prevalence were recomputed within each group's matched subsample and compared to the corresponding unmatched ("original") estimates.
